# A systematic review of psychological factors influencing attitudes and intentions toward, and uptake of, Covid-19 vaccines in adolescents

**DOI:** 10.1101/2024.09.17.24313392

**Authors:** Angie Pitt, Richard Amlôt, Catherine Heffernan, G. James Rubin, Louise E. Smith

## Abstract

Vaccination was a key measure to tackle the Covid-19 pandemic, however adolescents were less likely than adults to accept the vaccine. Low vaccine uptake reduces the effectiveness of vaccination campaigns and threatens global public health. Understanding why adolescents are hesitant to accept new vaccines is therefore crucial to support the development of novel vaccine uptake interventions. Prior reviews have included far fewer citations, excluded qualitative data, studies after 2022 and have not mapped adolescent Covid-19 vaccine behaviour onto psychological models. This systematic review investigated psychological factors influencing attitudes and intentions toward and uptake of Covid-19 vaccines in adolescents aged 10 to 19 years globally. It mapped results onto the COM-B framework to inform future interventions. Our search identified 25,354 citations, and included 77 in this review. The quality of studies was mixed, predominantly cross-sectional in design. According to our review, key influences on adolescent Covid-19 vaccine behaviour were: i) Reflective motivation (safety concerns, perceived susceptibility to/severity of Covid-19, perceived vaccine effectiveness, ii) Social opportunity (social norms, autonomy and prosocial attitudes), iii) Psychological capability (attitude and knowledge about vaccines). Our review provides new insights into psychological factors influencing adolescent Covid-19 vaccine behaviour, and maps factors to the COM-B model of behaviour change. To improve vaccine uptake, future vaccine interventions should support adolescents to think critically about the pros and cons of vaccines and consider external influences on their decisions.

## 1. Introduction

The Covid-19 pandemic killed over 7 million people worldwide as of spring 2024 ^1^ and led to substantial psychological, social and economic disruption. For children and adolescents, school closures and physical distancing measures led to a decline in physical and mental health and broadened existing disparities between the richest and poorest in society ^2–4^.

Vaccination was identified by the World Health Organization (WHO) as key to ending the crisis ^5^. Cross-population uptake of vaccinations was required to protect the public and mitigate the spread of the virus preventing further social and economic disruption (Zimet et al., 2020). The first Covid-19 vaccines were approved for use in adults in 2020, and in adolescents in 2021, e.g., May 2021 in the USA ^6^. However, the effectiveness of these programmes was threatened by vaccine hesitancy ^7^, defined as a delay in acceptance or refusal of vaccination despite the availability of vaccinations ^8^.

Covid-19 vaccine uptake in adolescents lags behind that in adults. In the USA, for example, 17.4% of adults are unvaccinated, compared to 26.4% of 12-15 year-olds and 19% of 16-17 year-olds ^9^. Adolescence is defined by the World Health Organization (WHO) ^10^ as ages 10 to 19 years. During this life stage, adolescents define their social identity ^11, 12^, gain autonomy ^13^ and experience intense neurodevelopmental changes. While the adolescent neurological reward system becomes hyperresponsive, leading to increased reward-seeking behaviour, the prefrontal cortex is not fully developed, limiting adolescents’ ability to regulate this behaviour ^14^. As a result, adolescents often prioritize short-term rewards over long-term benefits, influenced by emotions and peer interactions.

Studies investigating Covid-19 vaccine behaviour in adolescents have implicated multiple psychological factors. Two prior systematic reviews ^15, 16^ found concerns over vaccine safety, efficacy and side effects, low perceived necessity and needle phobia were all barriers to adolescent Covid-19 vaccination acceptance. However, these reviews’ searches were limited, and so included only 15 and seven studies respectively, and did not include qualitative data, grey literature, or studies post-2022. Further, no studies to date have examined how behaviour theory maps onto adolescent Covid-19 vaccine behaviour: a necessary step in the development of behaviour-change interventions. Previous adolescent-facing vaccine interventions have aimed to increase knowledge, with mixed results ^17^. We will map our findings onto COM-B ^18^ given this framework allows the consideration of both internal and external factors. COM-B classifies these factors as capability, opportunity, and motivation.

Capability is defined as an individual’s physical (e.g., ability or strength) and psychological (e.g., knowledge, memory) capacity to engage in the behaviour. Opportunity is defined as external factors that enable or prompt the behaviour, i.e., an individual’s physical and social environment. Motivation covers reflective and automatic brain processes that direct behaviour. COM-B has previously been used to explain health behaviours including vaccine behaviour in adults ^19^, health behaviours in adolescents ^20, 21^ and medication adherence in the general population ^22^. In this review we will synthesise psychological factors influencing Covid-19 vaccine-related attitudes, intentions, and behaviours in adolescents. We will then examine how these factors map to components of COM-B, drawing implications for future adolescent vaccination interventions.

## 2. Methods

### 2.1 Identification of studies

The protocol was registered with Prospero (CRD42023406768). We followed the Preferred Reporting Items for Systematic Reviews and Meta-Analyses (PRISMA) guidelines ^23^.

Searches of PsycINFO, Embase, MEDLINE and Scopus were carried out on 31 March 2023. We conducted reference and forward citation screening of included studies, and a grey literature search including screening the first 100 results of a Google search on 15 September 2023. The search was repeated on 9 April 2024, and we screened reference and forward citations of newly included studies on 16 April 2024. Search terms combined subject headings and free text searches for: Covid-19 terms (SARS-CoV-2 or coronavirus); vaccines (immunisation, inoculation); adolescents (e.g., teen, youth); and attitudes (e.g., beliefs, views) with publication from 1 January 2020 (Appendix A).

### 2.2 Selection criteria

Studies were eligible for inclusion if participants were aged 10 to 19 years, (including 10- to 19-year-olds included in a study as a subgroup); gave self-reported reasons for vaccine intention or uptake and/or provided statistical analyses of self-reported psychological factors relating to vaccine attitude, intention, or uptake; and if a full text English version was available. Any study design was eligible for inclusion. We excluded studies where the sample were higher education students, pregnant or parents.

### 2.3 Data extraction

We extracted information about study design and methods, location, date of data collection, outcome measures, and results of studies relating to psychological factors influencing attitudes and intention toward, and uptake of the Covid-19 vaccine.

### 2.4 Quality assessment

We used the National Heart Lung and Blood Institute (NIH) Quality Assessment Tool for observational cohort and cross-sectional studies ^24^ to assess cross-sectional studies, and the NIH Before-After Studies with No Control Group tool for interventions ^24^. We used the Critical Appraisal Skills Programme (CASP) ^25^ tool to assess qualitative studies. Where studies employed mixed methods, quantitative and qualitative sections were assessed separately using the above tools. AP and LES independently quality assessed 10% of studies to check consistency, resolving disagreements through discussion. AP assessed the remaining studies with LES supporting.

### 2.5 Procedure

We investigated:

- Self-reported reasons for vaccination acceptance or hesitancy (intended or completed uptake; quantitative data)
- Psychological factors associated with vaccine attitudes, intention, and uptake (quantitative data).
- Barriers and facilitators to vaccine intention and uptake (qualitative data).

AP conducted all searches, with guidance from LES. LES independently screened the first 100 citations with consistent results. Where participant age was unclear, we contacted study authors for clarification. Where authors did not reply, citations were excluded. Remaining screening and data extraction were carried out by AP with LES supporting. Where multiple papers and conference abstracts reported the same study, all reported data were considered together. We synthesised quantitative results according to Synthesis Without Meta-analysis (SWiM) guidelines ^26^. Results have been included in the synthesis if the factor was investigated in at least two studies. Quantitative results were synthesised narratively. Qualitative results were synthesised following Noblit and Hare’s ^27^ seven stages of meta-ethnography, with themes coded in NVivo and new overarching themes generated. We then conducted a secondary analysis, mapping factors relating to intention or uptake of the vaccine onto components of COM-B ^18^ and combining quantitative and qualitative results. Where factors did not map on to the COM-B model, for example because they could impact capability, motivation, or opportunity, we synthesised these separately. Willingness to accept a vaccine was defined as intention to accept a vaccine ^28^. In our review, we used ***vaccine hesitancy*** to encompass reluctance, doubts, and refusal to accept or intend to accept a vaccine. We defined ***vaccine acceptance*** as the willingness to accept or intend to accept a vaccine ^29^. If studies differentiated between vaccine hesitancy and vaccine reluctance, we clarified in the results. Definitions of vaccine attitude were based on individual study definitions, however where attitude was measured as **willingness to be vaccinated**, these studies were categorized with the outcome as **intention**.

## 3. Results

Searches of Embase, PsycINFO, Medline and Scopus yielded 25,354 citations. Following screening, 77 citations were included in the review, describing 73 studies (Figure 1).

**Figure 1.**
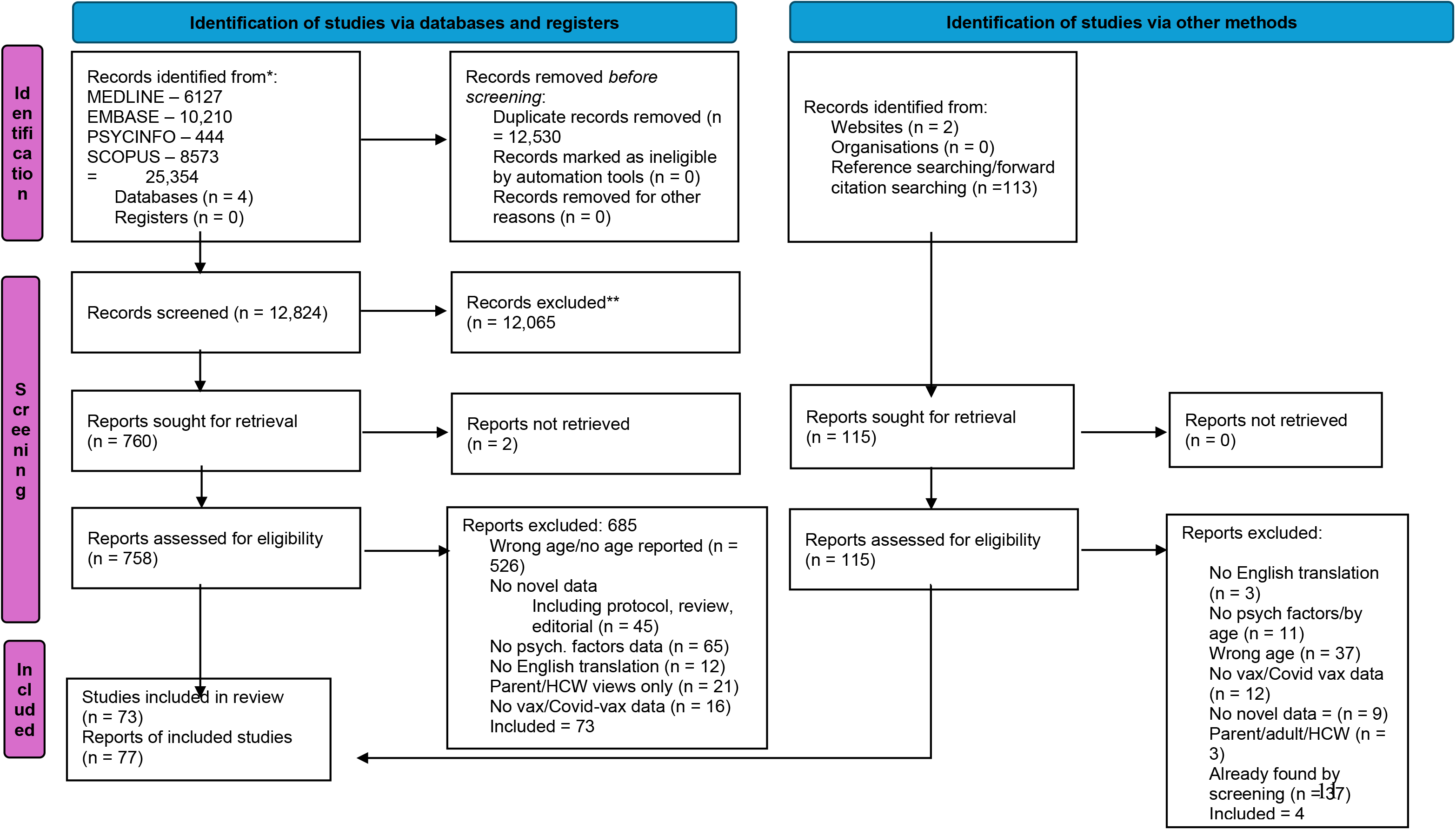
PRISMA Flow Chart.

### 3.1 Study characteristics

Table 1 provides an overview of key characteristics of all included studies. Included studies were from Africa (n = 7), Asia (n = 26), Europe (n = 10), the Middle East (n = 3), North America (n = 24) and South America (n = 2). Three studies compared data from different countries ^30–32^. The number of relevant participants ranged from 6 to 272 914. Study designs were cross-sectional (n = 59) qualitative (n = 10), mixed methods (n = 5), retrospective cohort (n = 2) before-after design with no control group (n = 1).

**Table 1.**
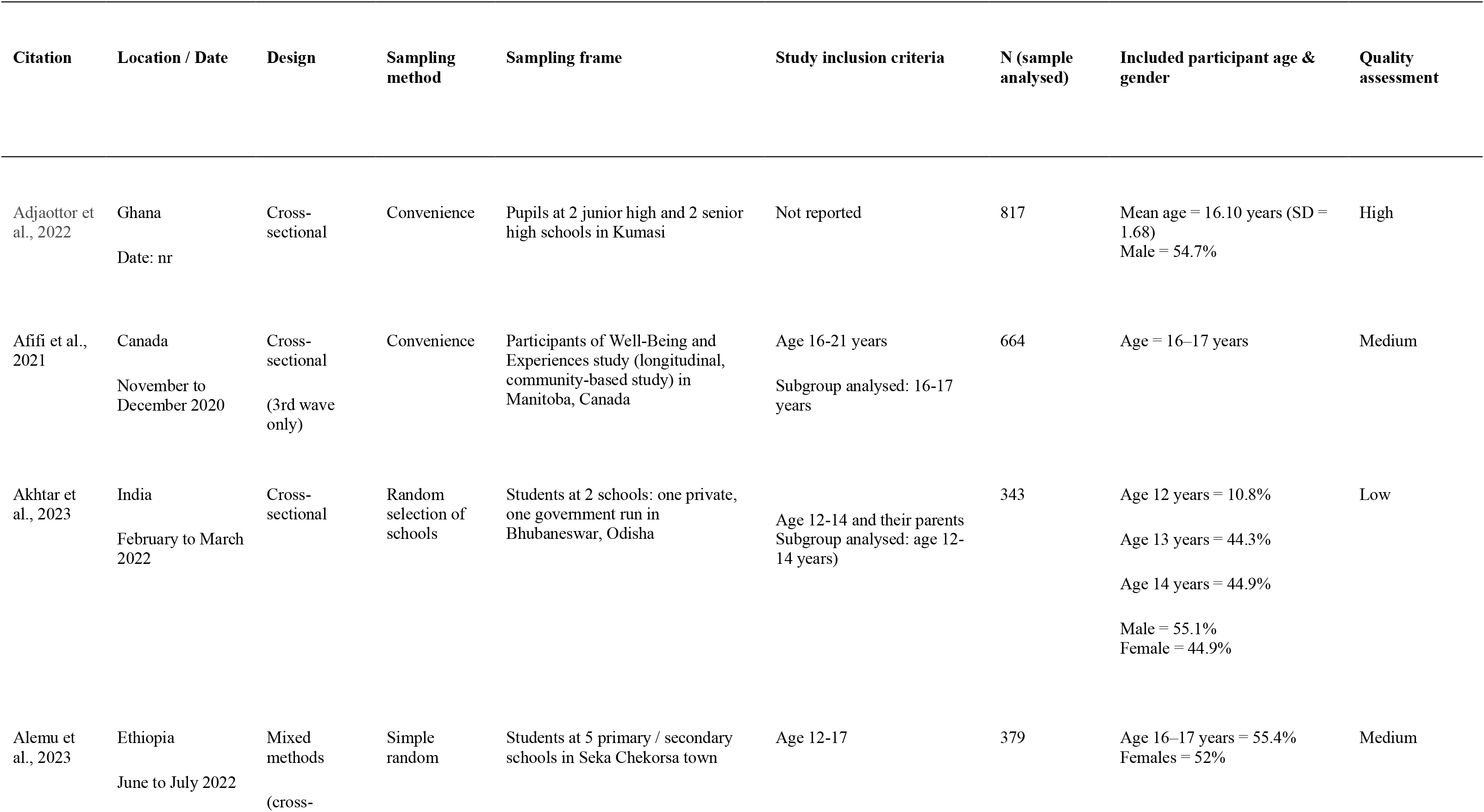
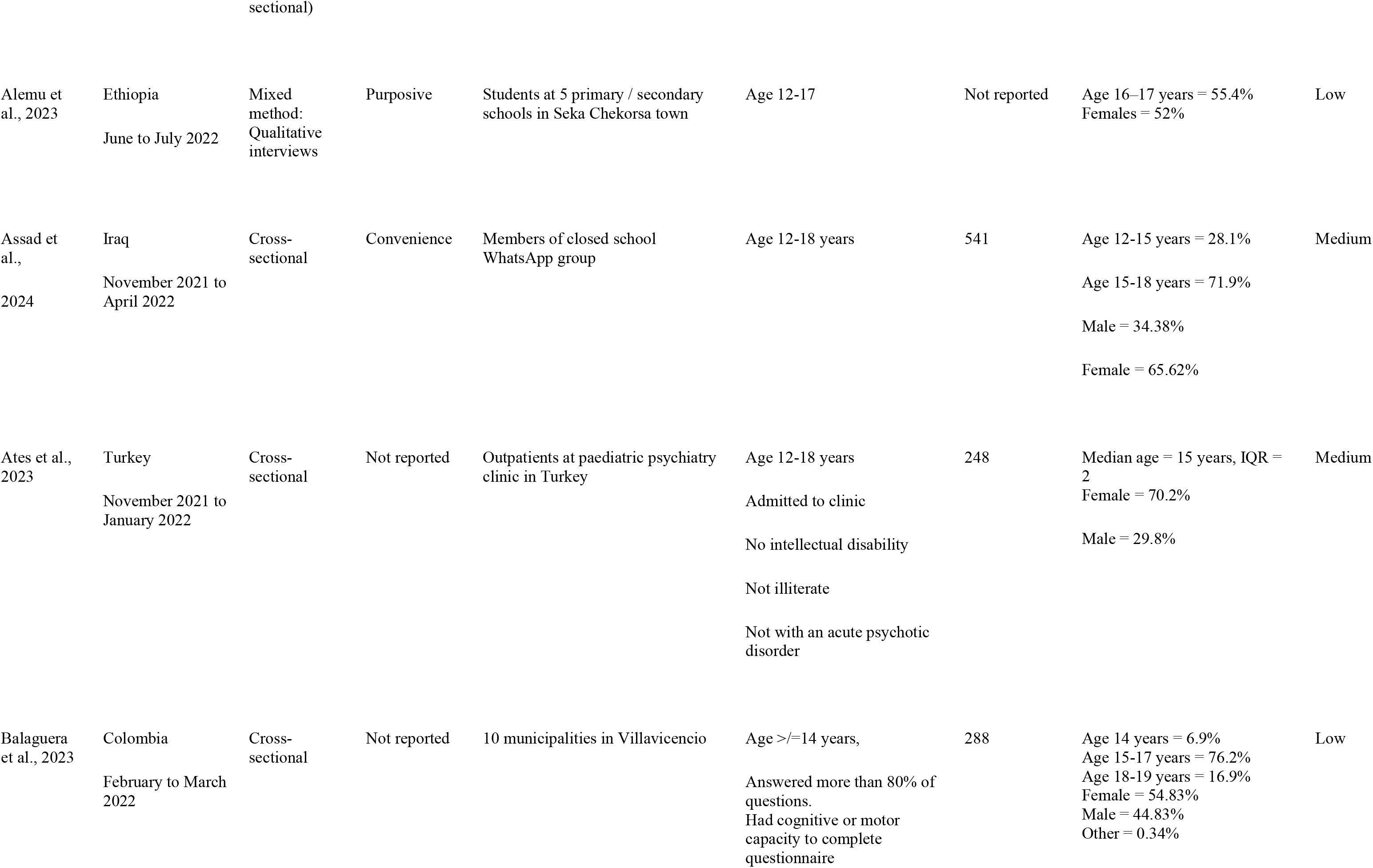
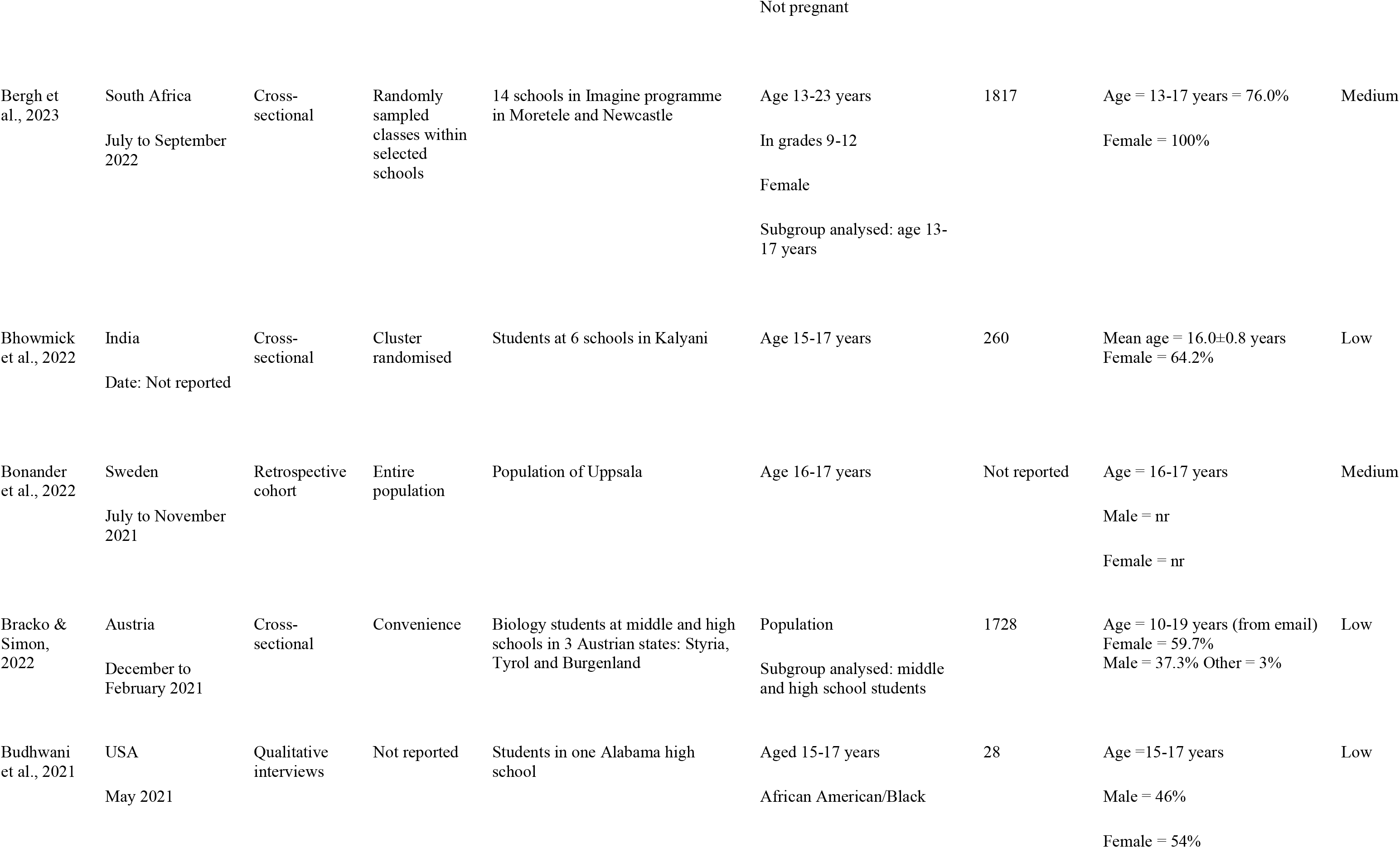
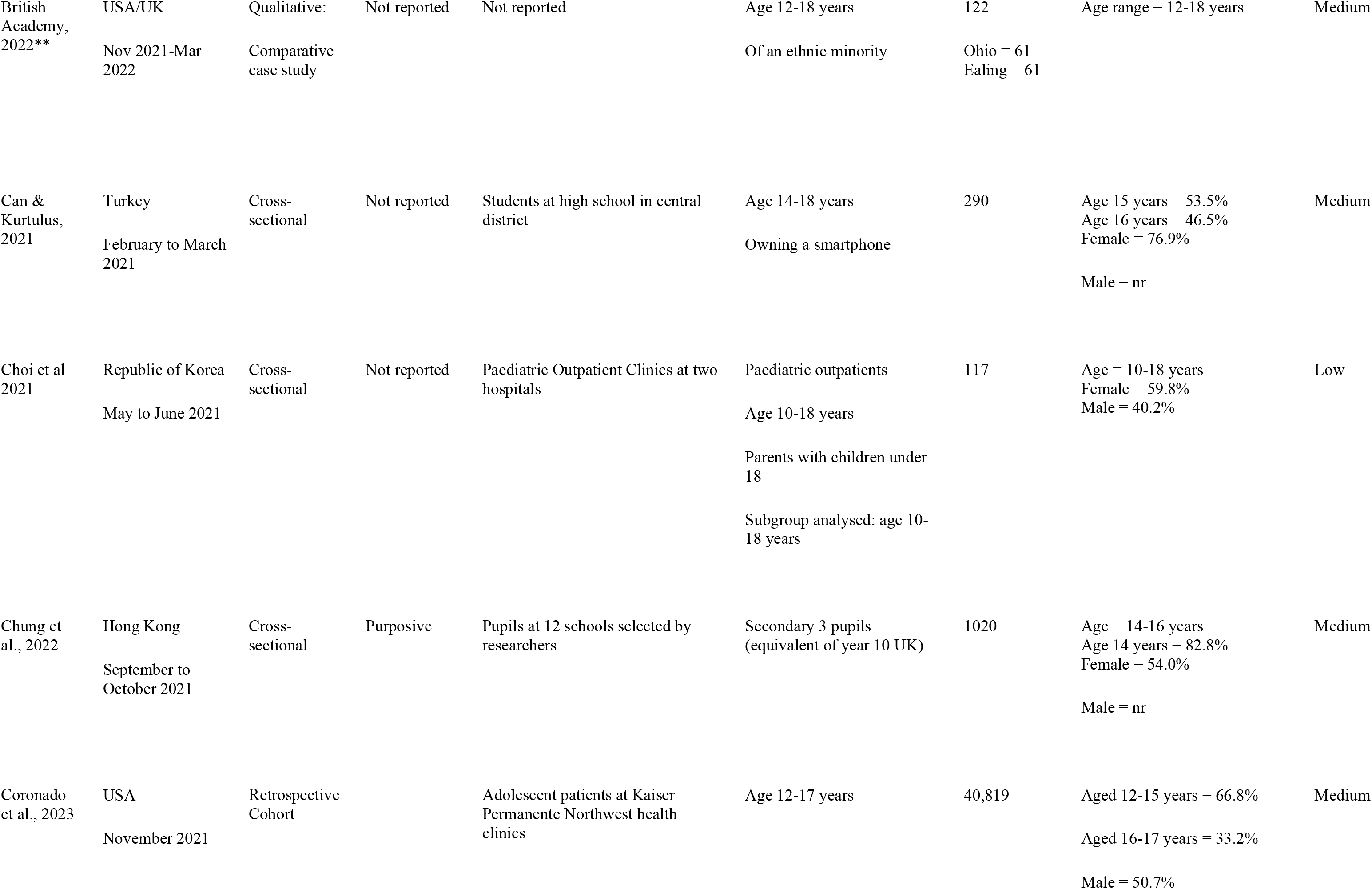
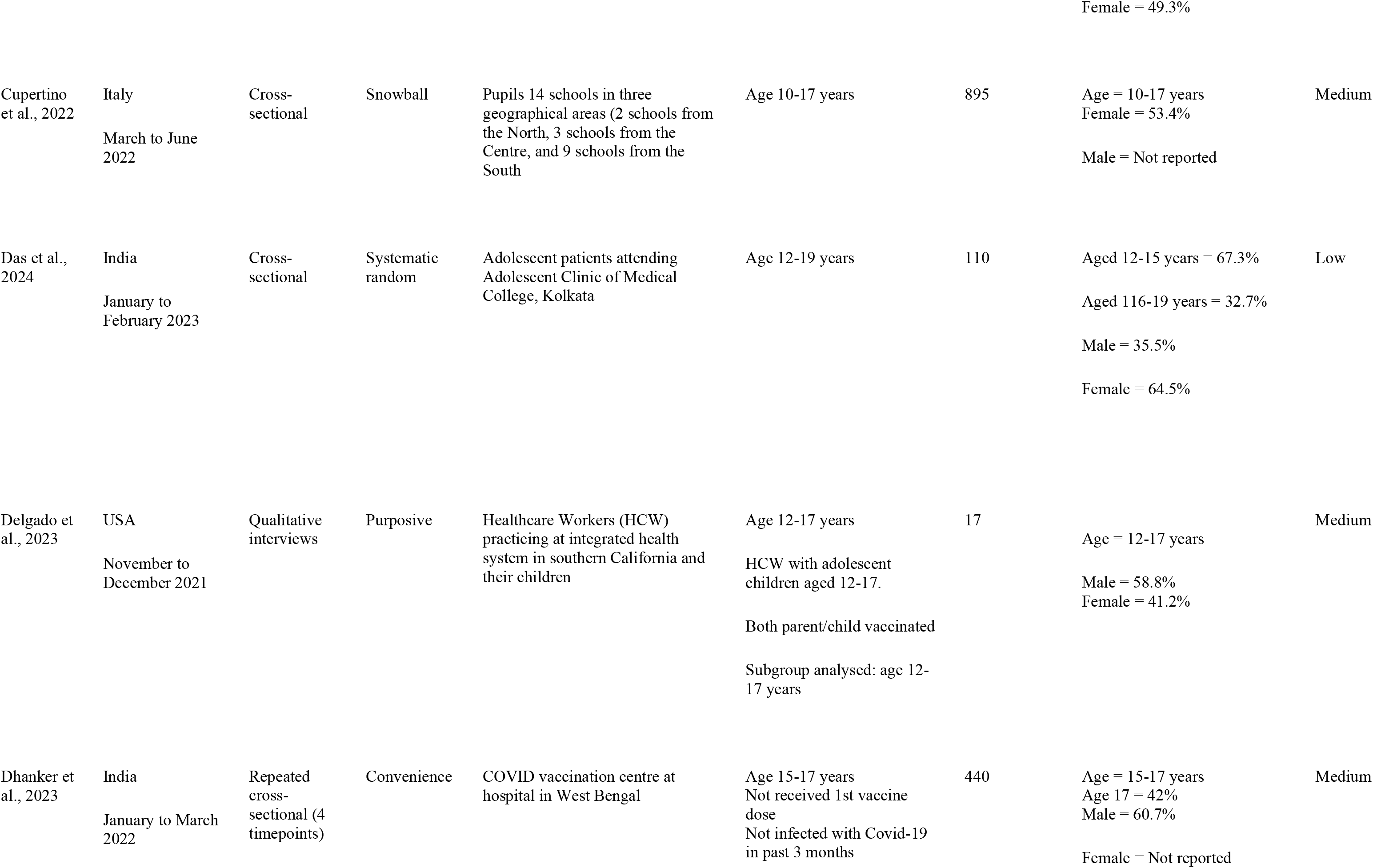
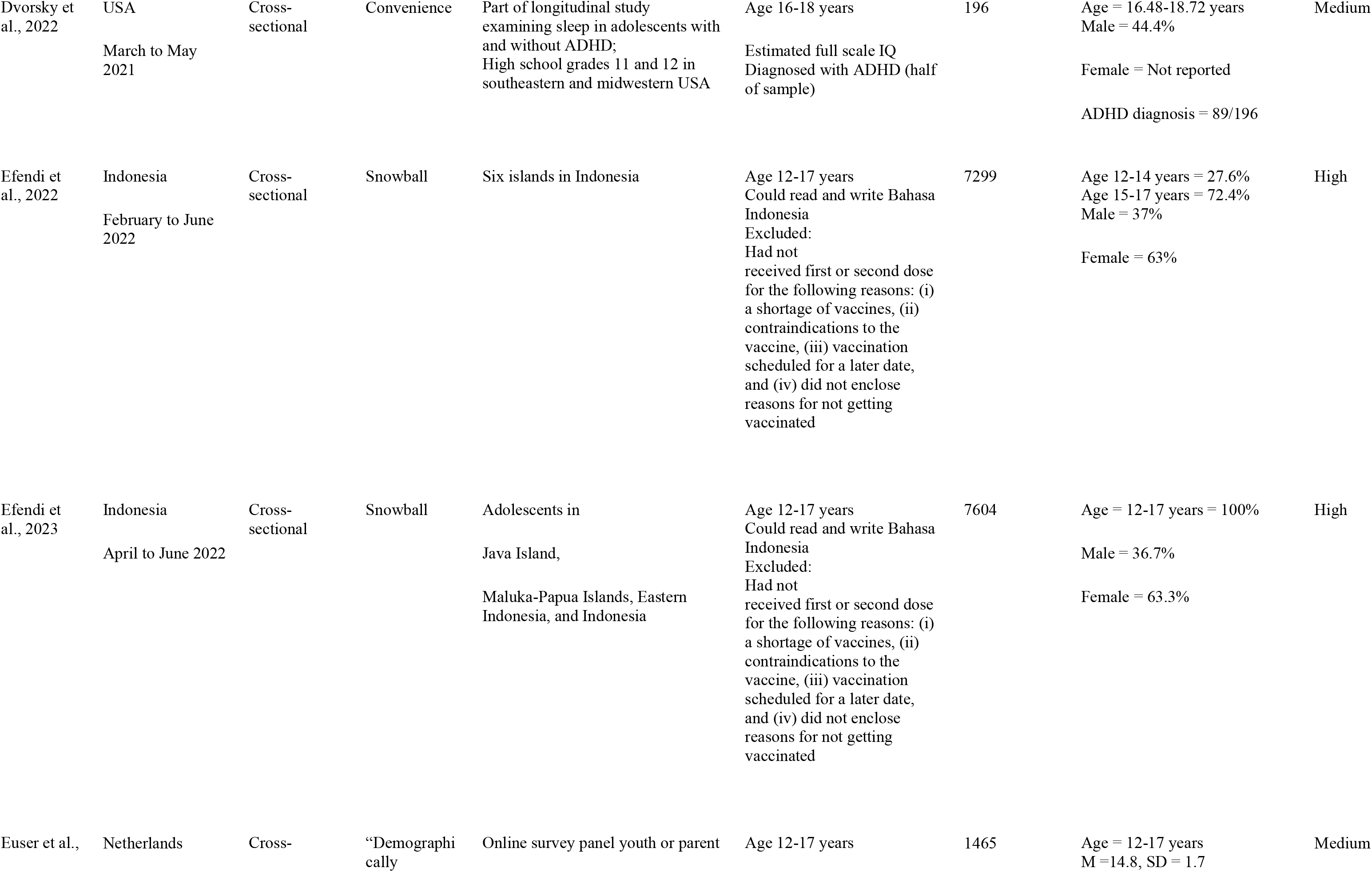
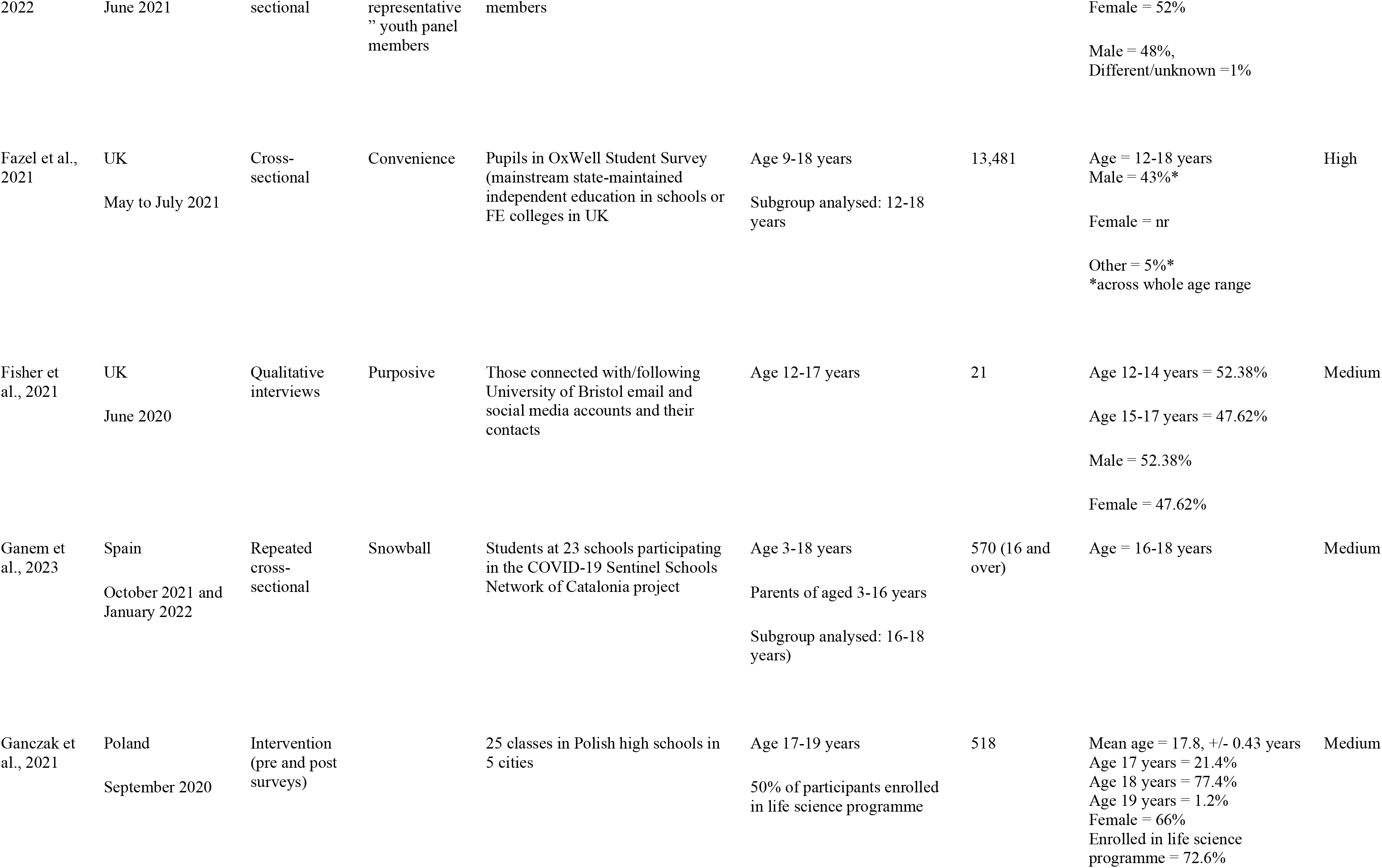
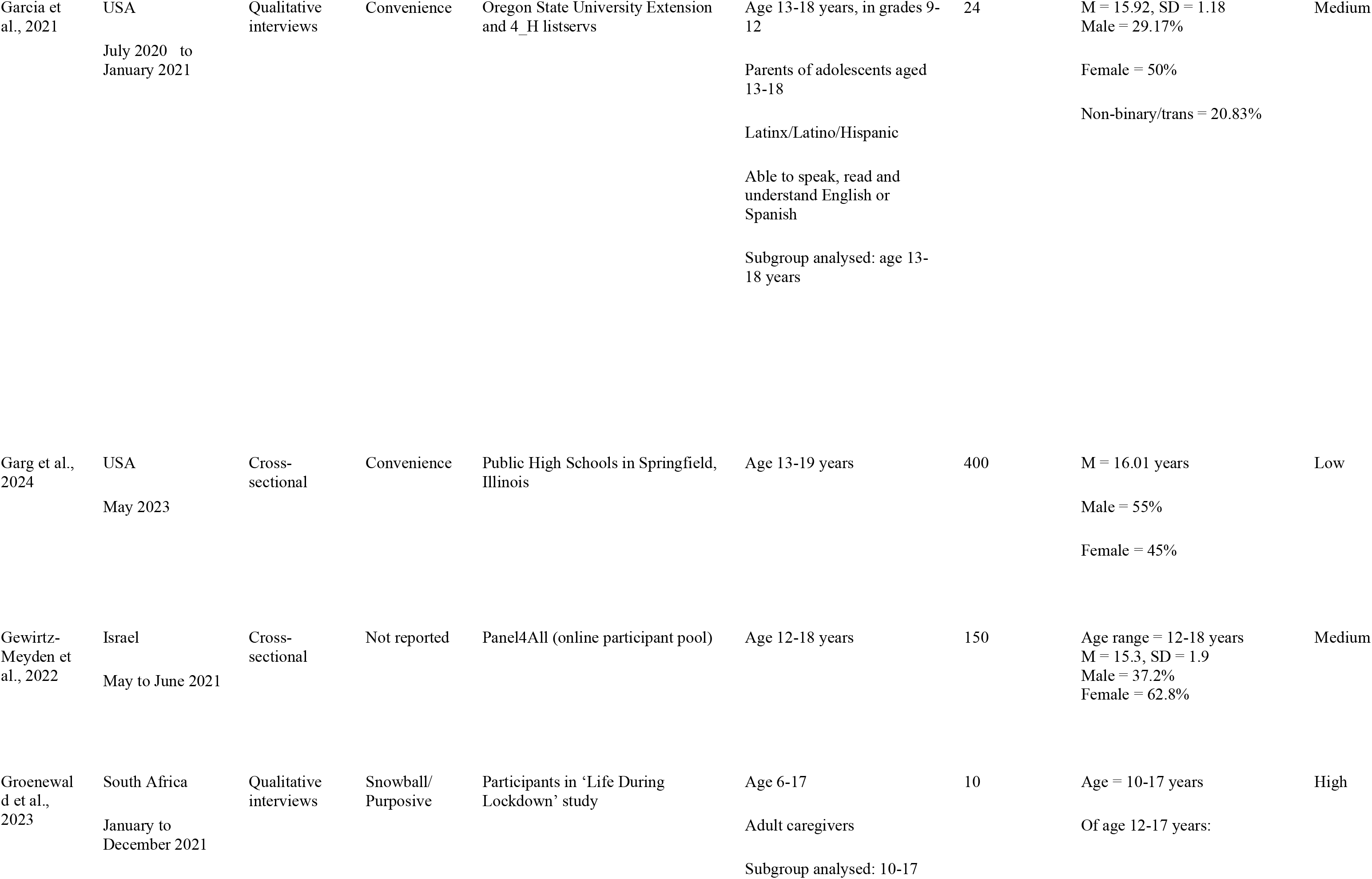
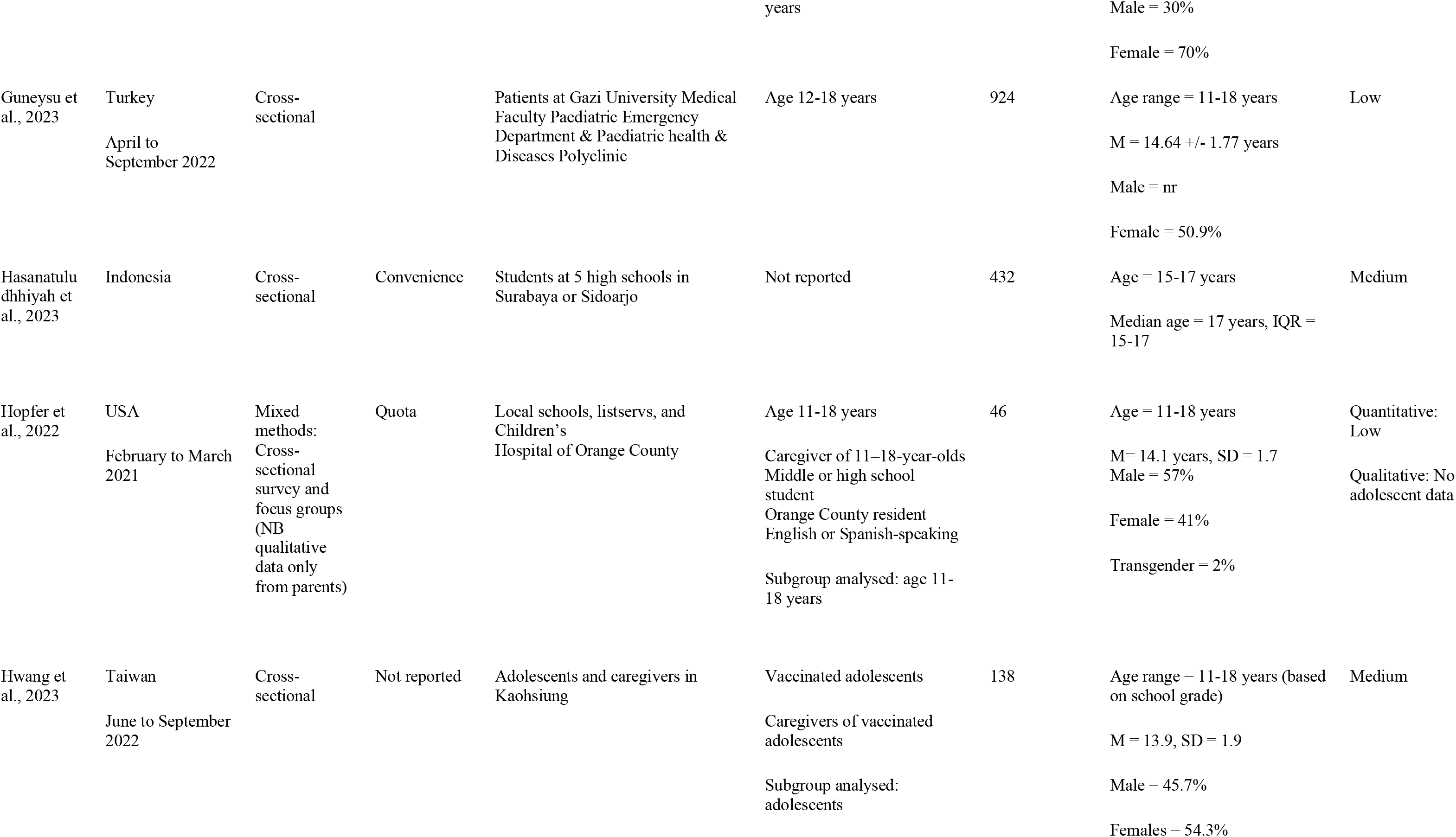
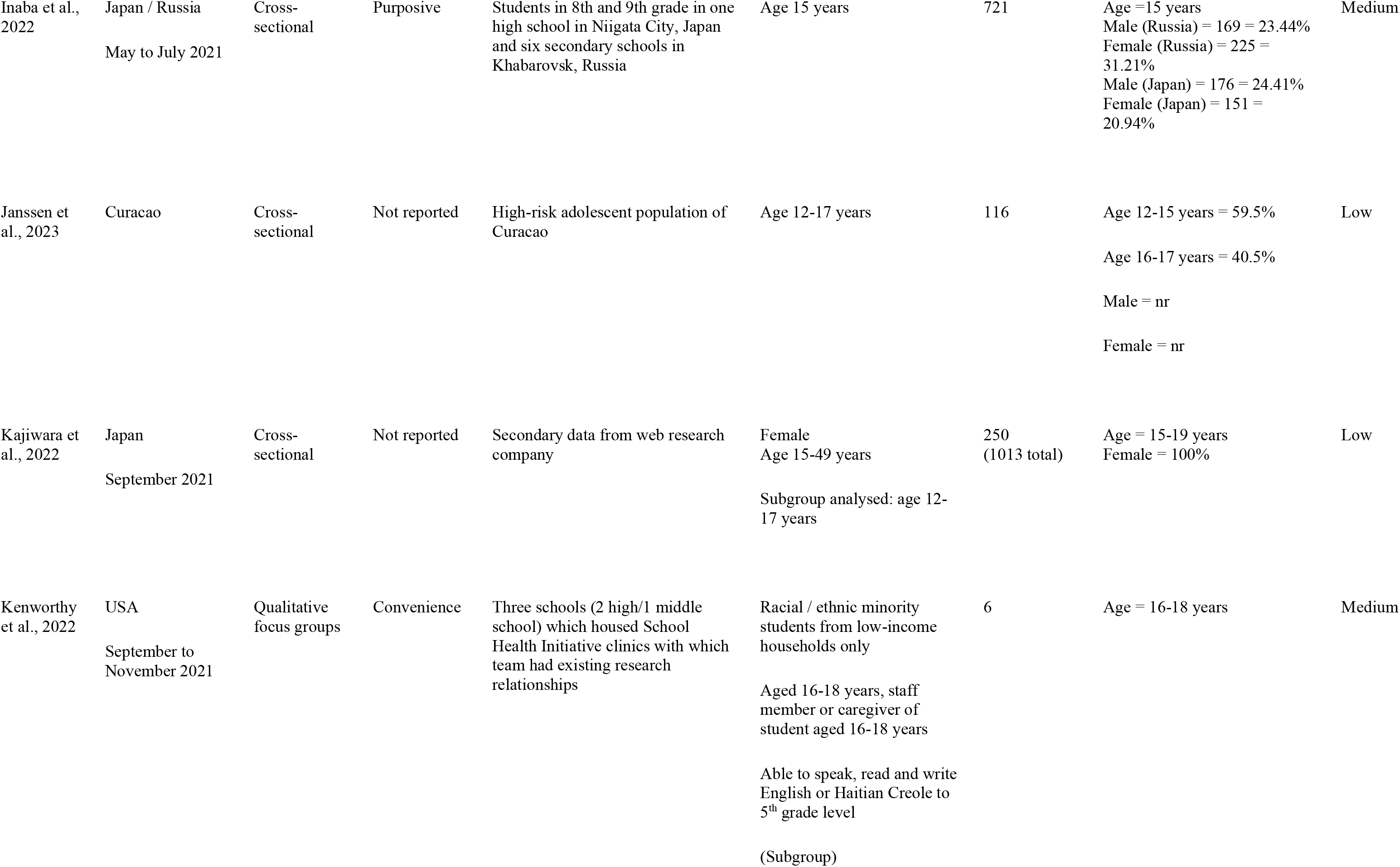
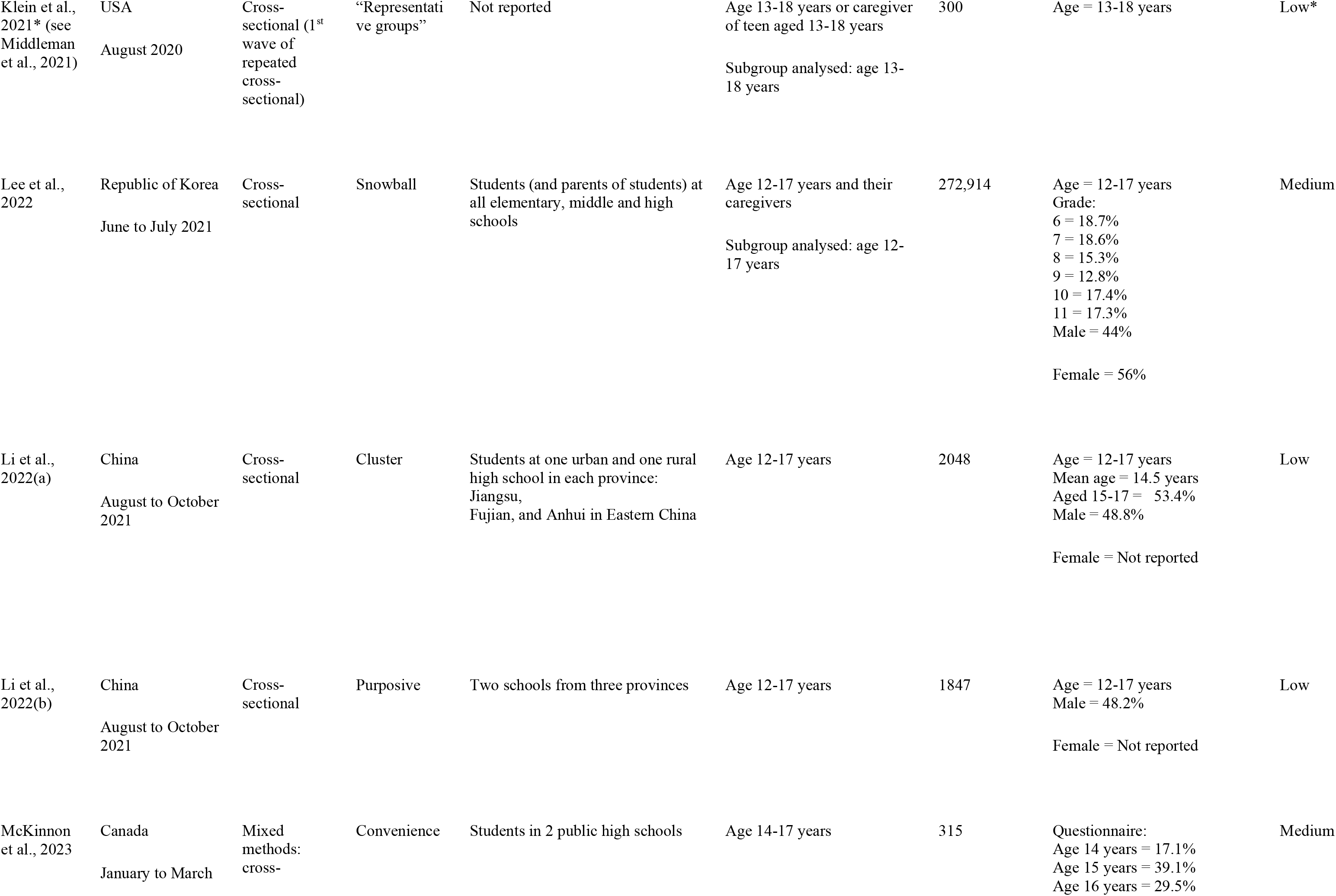
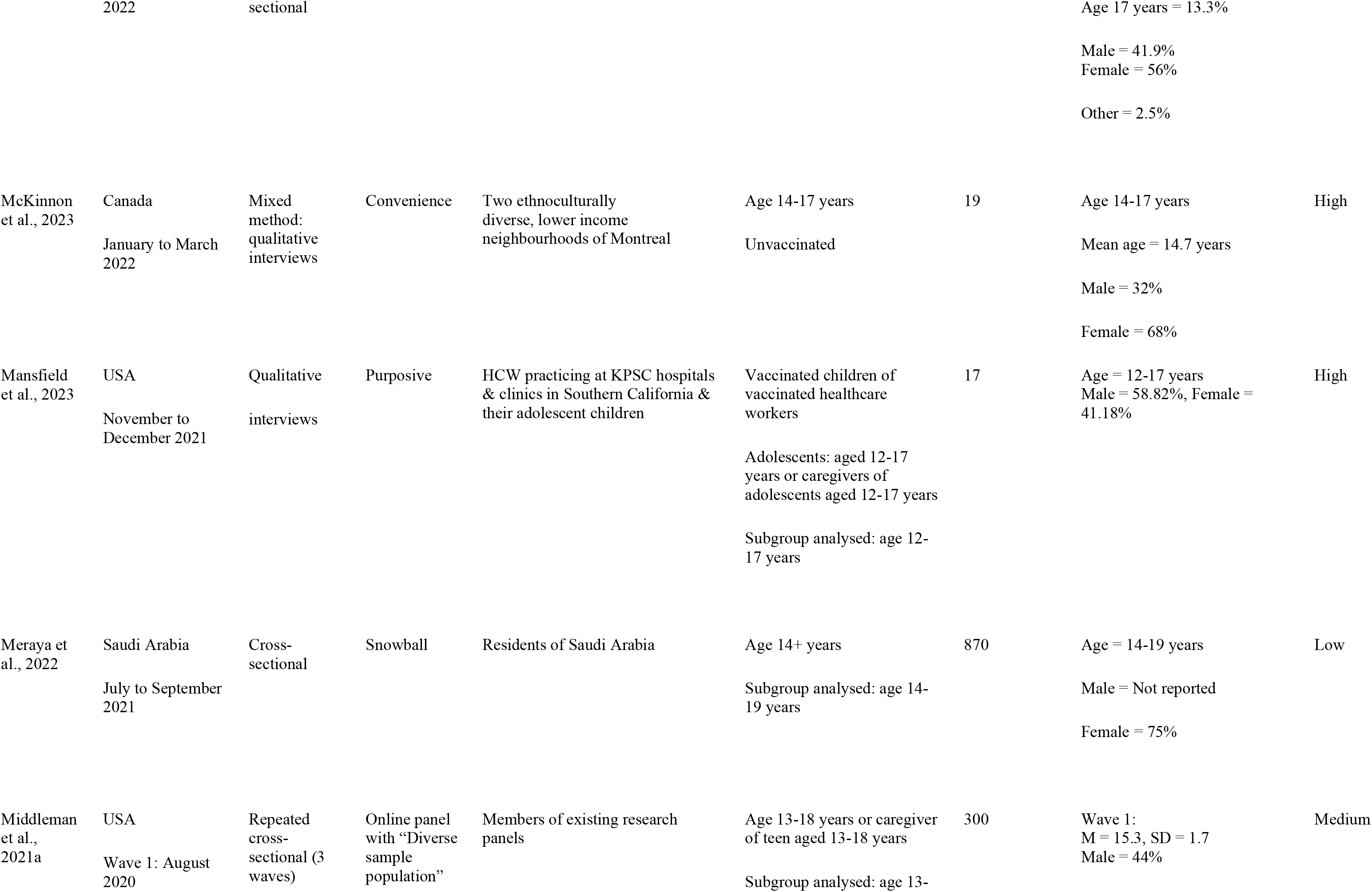
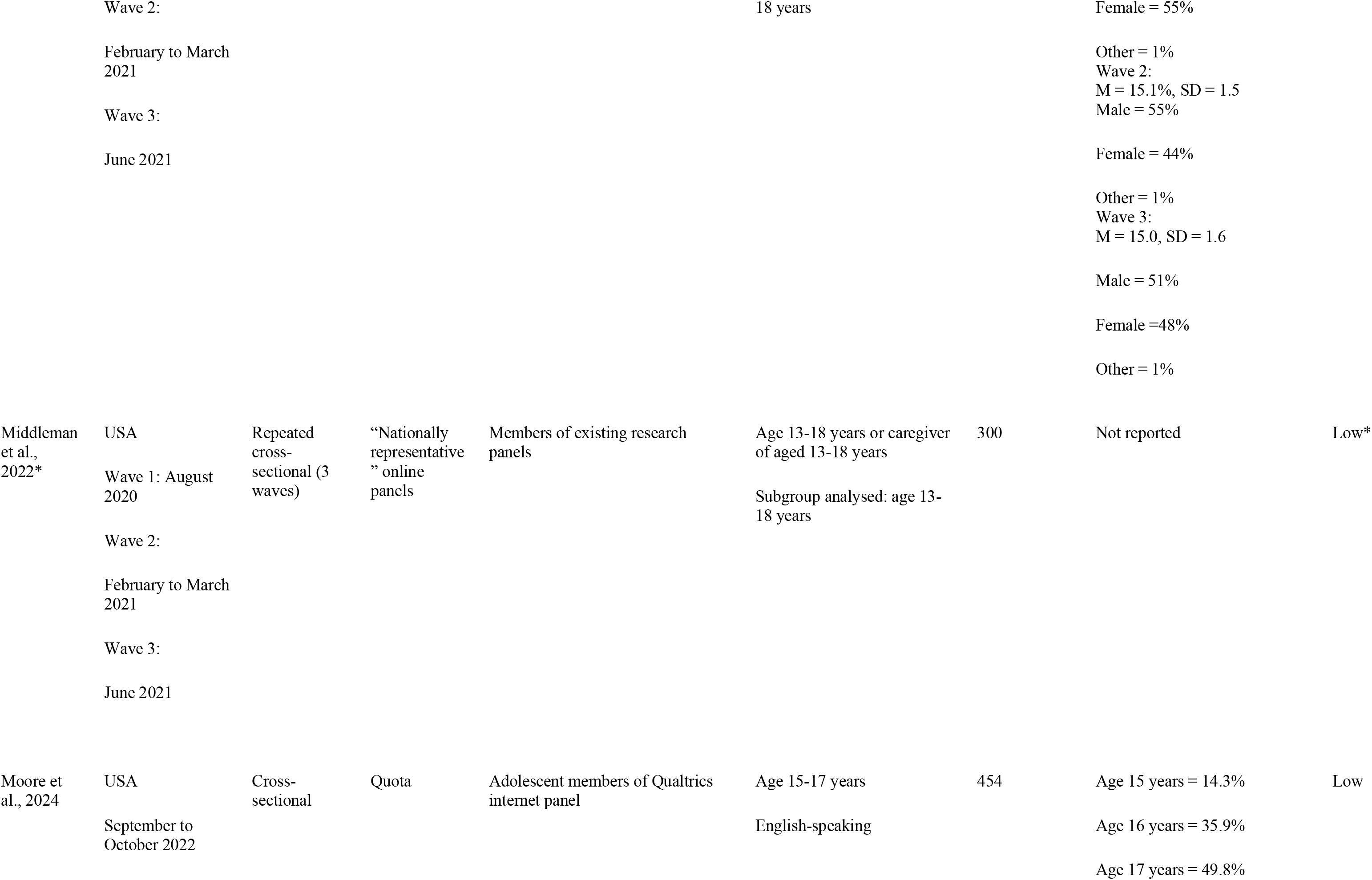
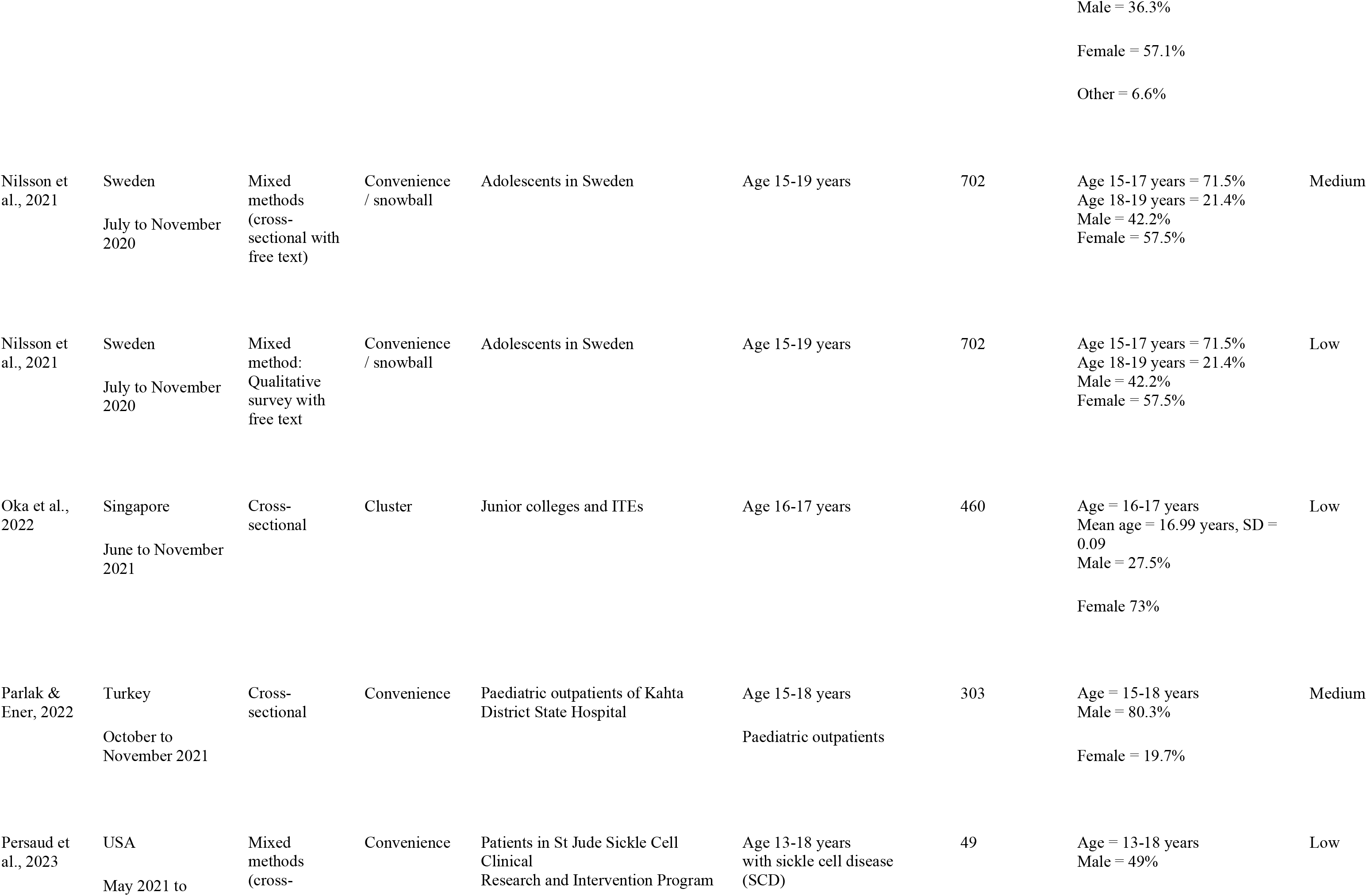
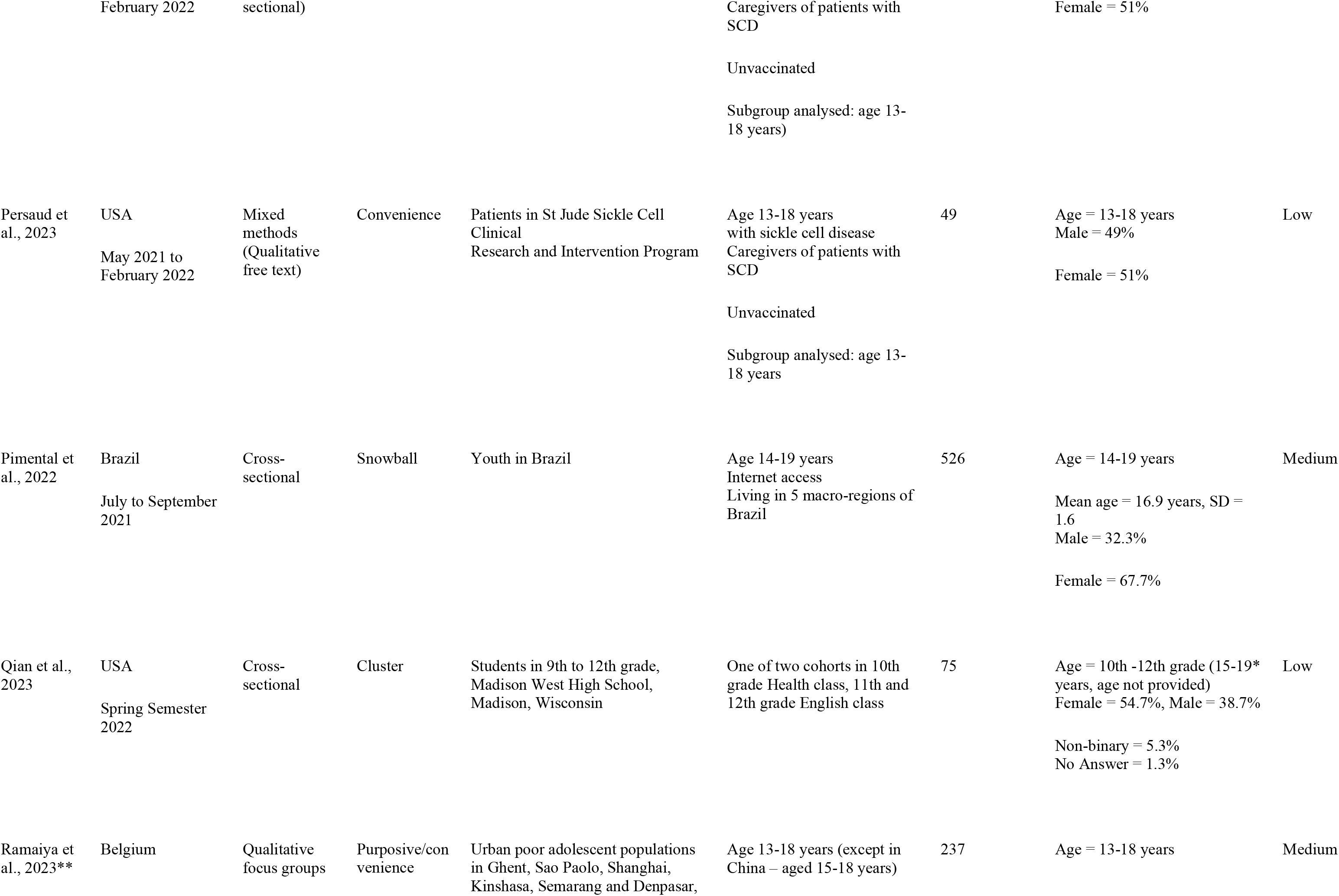
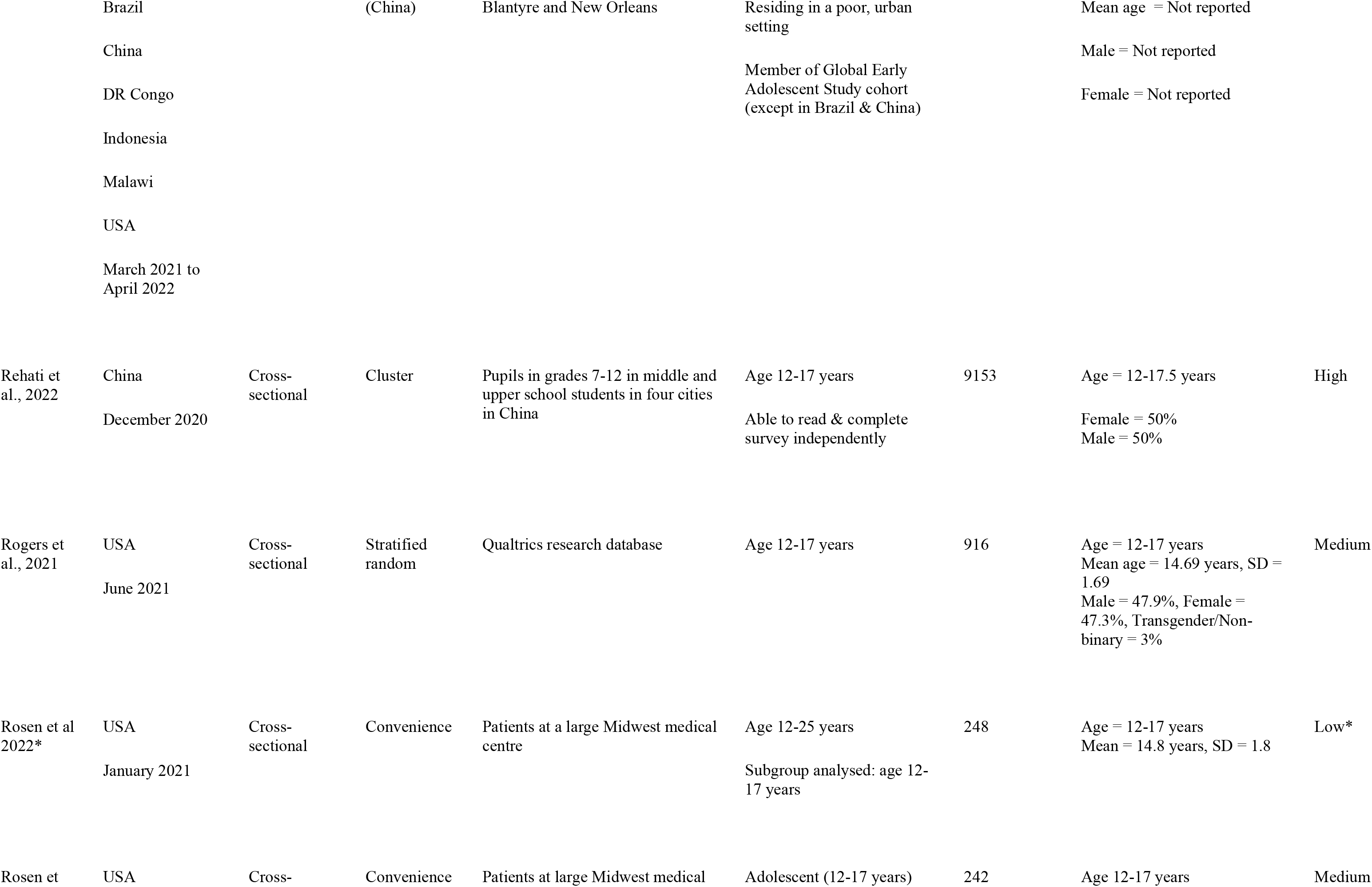
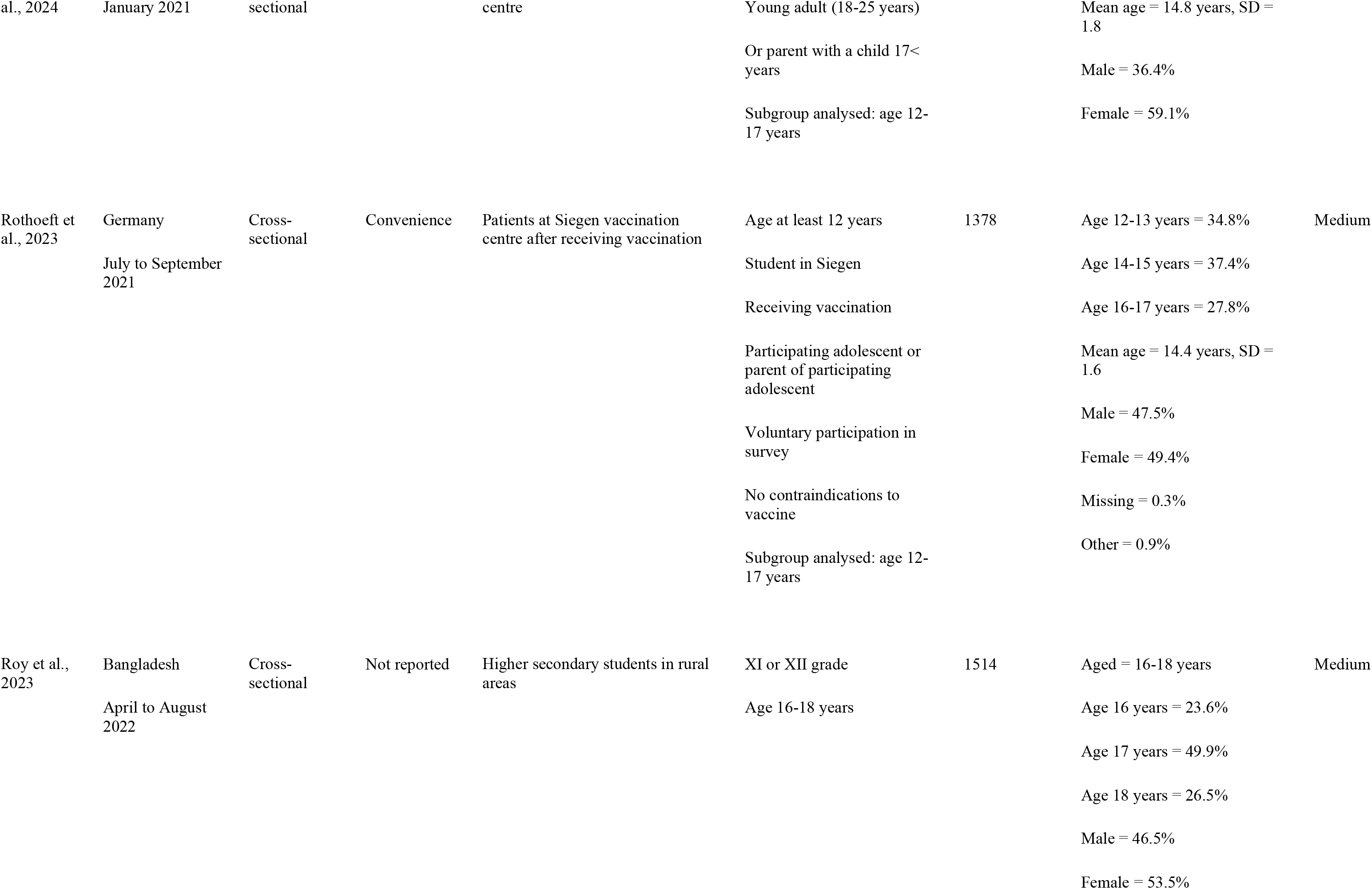
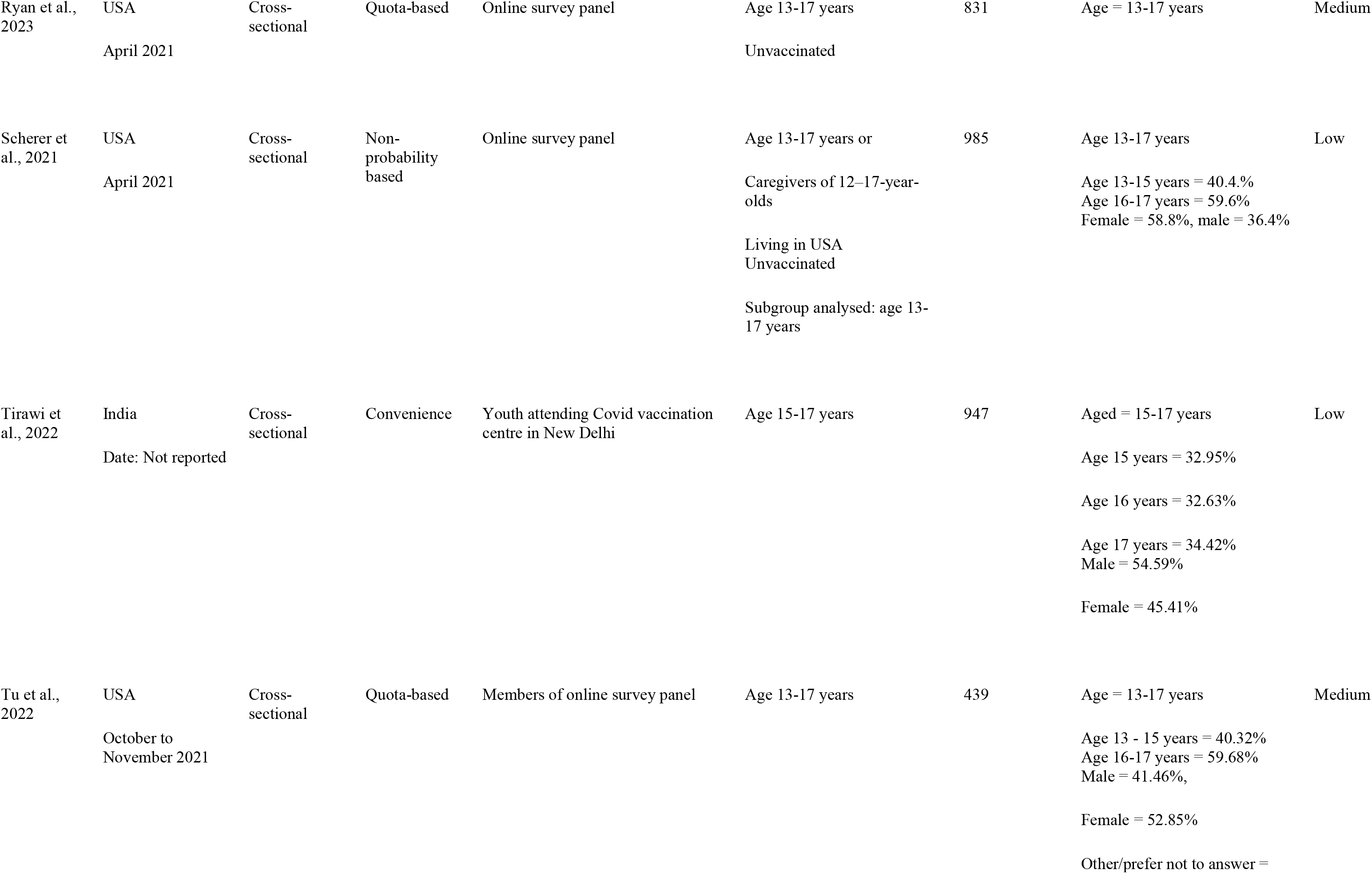
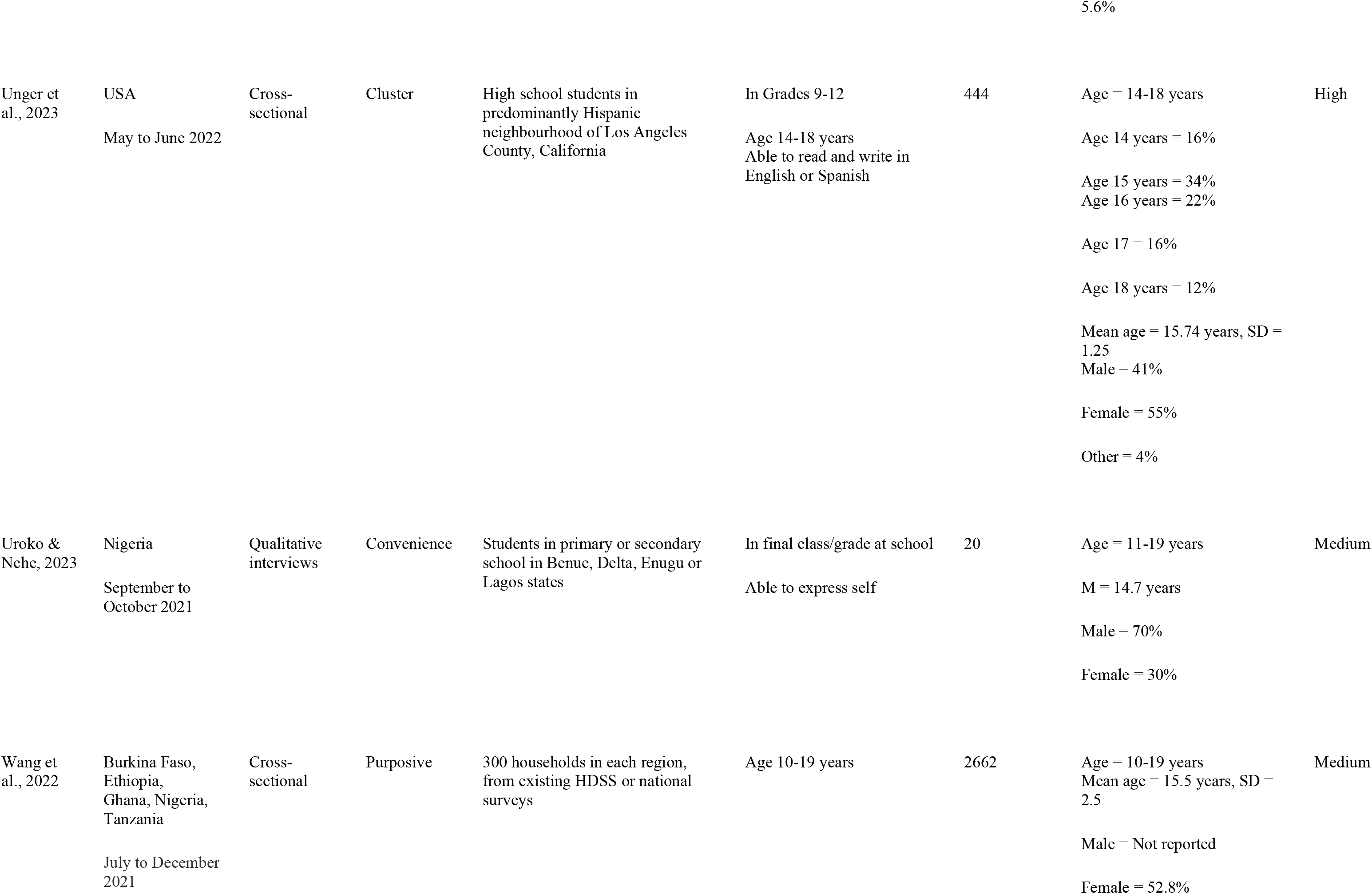
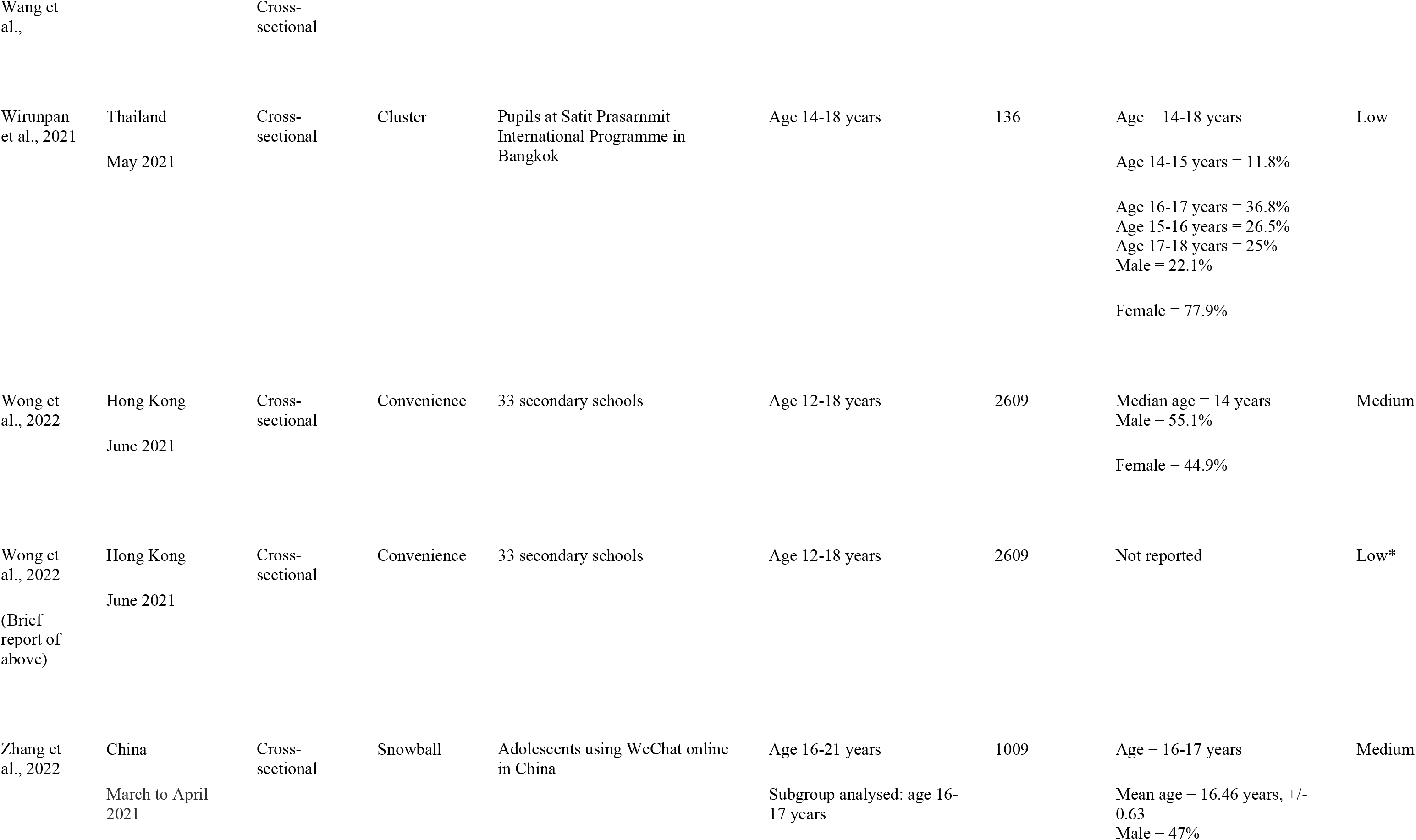
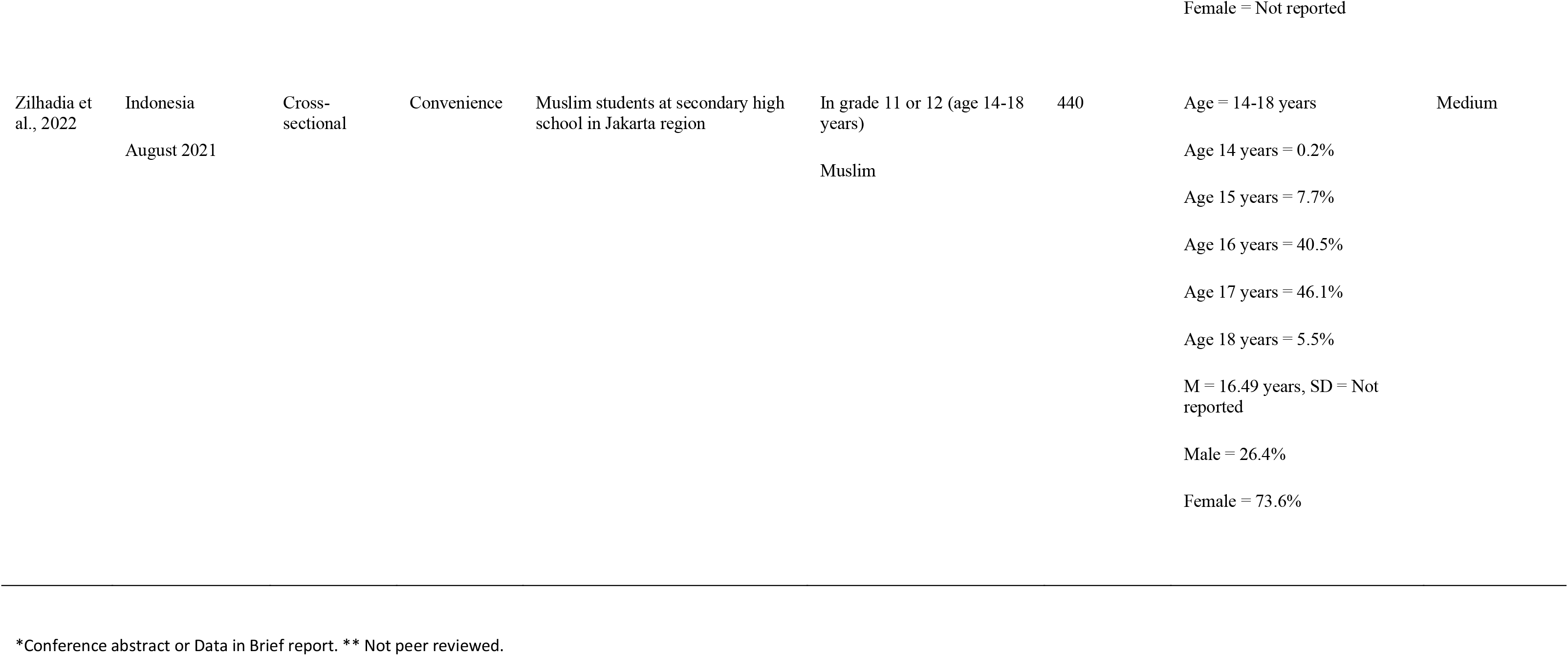
Characteristics of included studies.

| Citation | Location / Date | Design | Sampling method | Sampling frame | Study inclusion criteria | N (sample analysed) | Included participant age & gender | Quality assessment |
| --- | --- | --- | --- | --- | --- | --- | --- | --- |
| Adjaottor et al., 2022 | Ghana<br>Date: nr | Cross-sectional | Convenience | Pupils at 2 junior high and 2 senior high schools in Kumasi | Not reported | 817 | Mean age = 16.10 years (SD = 1.68)<br>Male = 54.7% | High |
| Afifi et al., 2021 | Canada<br>November to December 2020 | Cross-sectional<br>(3rd wave only) | Convenience | Participants of Well-Being and Experiences study (longitudinal, community-based study) in Manitoba, Canada | Age 16-21 years<br>Subgroup analysed: 16-17 years | 664 | Age = 16–17 years | Medium |
| Akhtar et al., 2023 | India<br>February to March 2022 | Cross-sectional | Random selection of schools | Students at 2 schools: one private, one government run in Bhubaneswar, Odisha | Age 12-14 and their parents<br>Subgroup analysed: age 12-14 years) | 343 | Age 12 years = 10.8%<br>Age 13 years = 44.3%<br>Age 14 years = 44.9%<br>Male = 55.1%<br>Female = 44.9% | Low |
| Alemu et al., 2023 | Ethiopia<br>June to July 2022 | Mixed methods<br>(cross- | Simple random | Students at 5 primary / secondary schools in Seka Chekorsa town | Age 12-17 | 379 | Age 16–17 years = 55.4%<br>Females = 52% | Medium |
| Alemu et al., 2023 | Ethiopia<br>June to July 2022 | Mixed method:<br>Qualitative interviews | Purposive | Students at 5 primary / secondary schools in Seka Chekorsa town | Age 12-17 | Not reported | Age 16–17 years = 55.4%<br>Females = 52% | Low |
| Assad et al., 2024 | Iraq<br>November 2021 to April 2022 | Cross-sectional | Convenience | Members of closed school WhatsApp group | Age 12-18 years | 541 | Age 12-15 years = 28.1%<br>Age 15-18 years = 71.9%<br>Male = 34.38%<br>Female = 65.62% | Medium |
| Ates et al., 2023 | Turkey<br>November 2021 to January 2022 | Cross-sectional | Not reported | Outpatients at paediatric psychiatry clinic in Turkey | Age 12-18 years<br>Admitted to clinic<br>No intellectual disability<br>Not illiterate<br>Not with an acute psychotic disorder | 248 | Median age = 15 years, IQR = 2<br>Female = 70.2%<br>Male = 29.8% | Medium |
| Balaguera et al., 2023 | Colombia<br>February to March 2022 | Cross-sectional | Not reported | 10 municipalities in Villavicencio | Age >=14 years,<br>Answered more than 80% of questions.<br>Had cognitive or motor capacity to complete questionnaire | 288 | Age 14 years = 6.9%<br>Age 15-17 years = 76.2%<br>Age 18-19 years = 16.9%<br>Female = 54.83%<br>Male = 44.83%<br>Other = 0.34% | Low |
|  |  |  |  |  | Not pregnant |  |  |  |
| Bergh et al., 2023 | South Africa<br>July to September 2022 | Cross-sectional | Randomly sampled classes within selected schools | 14 schools in Imagine programme in Moretele and Newcastle | Age 13-23 years<br>In grades 9-12<br>Female<br>Subgroup analysed: age 13-17 years | 1817 | Age = 13-17 years = 76.0%<br>Female = 100% | Medium |
| Bhowmick et al., 2022 | India<br>Date: Not reported | Cross-sectional | Cluster randomised | Students at 6 schools in Kalyani | Age 15-17 years | 260 | Mean age = 16.0±0.8 years<br>Female = 64.2% | Low |
| Bonander et al., 2022 | Sweden<br>July to November 2021 | Retrospective cohort | Entire population | Population of Uppsala | Age 16-17 years | Not reported | Age = 16-17 years<br>Male = nr<br>Female = nr | Medium |
| Bracko & Simon, 2022 | Austria<br>December to February 2021 | Cross-sectional | Convenience | Biology students at middle and high schools in 3 Austrian states: Styria, Tyrol and Burgenland | Population<br>Subgroup analysed: middle and high school students | 1728 | Age = 10-19 years (from email)<br>Female = 59.7%<br>Male = 37.3% Other = 3% | Low |
| Budhwani et al., 2021 | USA<br>May 2021 | Qualitative interviews | Not reported | Students in one Alabama high school | Aged 15-17 years<br>African American/Black | 28 | Age =15-17 years<br>Male = 46%<br>Female = 54% | Low |
| British Academy, 2022** | USA/UK | Qualitative: | Not reported | Not reported | Age 12-18 years | 122 | Age range = 12-18 years | Medium |
|  | Nov 2021-Mar 2022 | Comparative case study |  |  | Of an ethnic minority | Ohio = 61<br>Ealing = 61 |  |  |
| Can & Kurtulus, 2021 | Turkey | Cross-sectional | Not reported | Students at high school in central district | Age 14-18 years | 290 | Age 15 years = 53.5%<br>Age 16 years = 46.5%<br>Female = 76.9% | Medium |
|  | February to March 2021 |  |  |  | Owning a smartphone |  | Male = nr |  |
| Choi et al 2021 | Republic of Korea | Cross-sectional | Not reported | Paediatric Outpatient Clinics at two hospitals | Paediatric outpatients | 117 | Age = 10-18 years<br>Female = 59.8%<br>Male = 40.2% | Low |
|  | May to June 2021 |  |  |  | Age 10-18 years<br>Parents with children under 18<br>Subgroup analysed: age 10-18 years |  |  |  |
| Chung et al., 2022 | Hong Kong | Cross-sectional | Purposive | Pupils at 12 schools selected by researchers | Secondary 3 pupils (equivalent of year 10 UK) | 1020 | Age = 14-16 years<br>Age 14 years = 82.8%<br>Female = 54.0% | Medium |
|  | September to October 2021 |  |  |  |  |  | Male = nr |  |
| Coronado et al., 2023 | USA | Retrospective Cohort |  | Adolescent patients at Kaiser Permanente Northwest health clinics | Age 12-17 years | 40,819 | Aged 12-15 years = 66.8% | Medium |
|  | November 2021 |  |  |  |  |  | Aged 16-17 years = 33.2%<br>Male = 50.7% |  |
|  |  |  |  |  |  |  | Female = 49.3% |  |
| Cupertino et al., 2022 | Italy<br>March to June 2022 | Cross-sectional | Snowball | Pupils 14 schools in three geographical areas (2 schools from the North, 3 schools from the Centre, and 9 schools from the South) | Age 10-17 years | 895 | Age = 10-17 years<br>Female = 53.4%<br>Male = Not reported | Medium |
| Das et al., 2024 | India<br>January to February 2023 | Cross-sectional | Systematic random | Adolescent patients attending Adolescent Clinic of Medical College, Kolkata | Age 12-19 years | 110 | Aged 12-15 years = 67.3%<br>Aged 116-19 years = 32.7%<br>Male = 35.5%<br>Female = 64.5% | Low |
| Delgado et al., 2023 | USA<br>November to December 2021 | Qualitative interviews | Purposive | Healthcare Workers (HCW) practicing at integrated health system in southern California and their children | Age 12-17 years<br>HCW with adolescent children aged 12-17.<br>Both parent/child vaccinated<br>Subgroup analysed: age 12-17 years | 17 | Age = 12-17 years<br>Male = 58.8%<br>Female = 41.2% | Medium |
| Dhanker et al., 2023 | India<br>January to March 2022 | Repeated cross-sectional (4 timepoints) | Convenience | COVID vaccination centre at hospital in West Bengal | Age 15-17 years<br>Not received 1st vaccine dose<br>Not infected with Covid-19 in past 3 months | 440 | Age = 15-17 years<br>Age 17 = 42%<br>Male = 60.7%<br>Female = Not reported | Medium |
| Dvorsky et al., 2022 | USA<br>March to May 2021 | Cross-sectional | Convenience | Part of longitudinal study examining sleep in adolescents with and without ADHD; High school grades 11 and 12 in southeastern and midwestern USA | Age 16-18 years<br>Estimated full scale IQ<br>Diagnosed with ADHD (half of sample) | 196 | Age = 16.48-18.72 years<br>Male = 44.4%<br>Female = Not reported<br>ADHD diagnosis = 89/196 | Medium |
| Efendi et al., 2022 | Indonesia<br>February to June 2022 | Cross-sectional | Snowball | Six islands in Indonesia | Age 12-17 years<br>Could read and write Bahasa Indonesia<br>Excluded:<br>Had not received first or second dose for the following reasons: (i) a shortage of vaccines, (ii) contraindications to the vaccine, (iii) vaccination scheduled for a later date, and (iv) did not enclose reasons for not getting vaccinated | 7299 | Age 12-14 years = 27.6%<br>Age 15-17 years = 72.4%<br>Male = 37%<br>Female = 63% | High |
| Efendi et al., 2023 | Indonesia<br>April to June 2022 | Cross-sectional | Snowball | Adolescents in<br>Java Island,<br>Maluka-Papua Islands, Eastern Indonesia, and Indonesia | Age 12-17 years<br>Could read and write Bahasa Indonesia<br>Excluded:<br>Had not received first or second dose for the following reasons: (i) a shortage of vaccines, (ii) contraindications to the vaccine, (iii) vaccination scheduled for a later date, and (iv) did not enclose reasons for not getting vaccinated | 7604 | Age = 12-17 years = 100%<br>Male = 36.7%<br>Female = 63.3% | High |
| Euser et al., | Netherlands | Cross- | “Demographically | Online survey panel youth or parent | Age 12-17 years | 1465 | Age = 12-17 years<br>M = 14.8, SD = 1.7 | Medium |
| 2022 | June 2021 | sectional | representative<br>" youth panel<br>members | members |  |  | Female = 52% |  |
|  |  |  |  |  |  |  | Male = 48%,<br>Different/unknown =1% |  |
| Fazel et al.,<br>2021 | UK<br>May to July 2021 | Cross-sectional | Convenience | Pupils in OxWell Student Survey<br>(mainstream state-maintained<br>independent education in schools or<br>FE colleges in UK | Age 9-18 years<br>Subgroup analysed: 12-18<br>years | 13,481 | Age = 12-18 years<br>Male = 43%*<br>Female = nr<br>Other = 5%*<br>*across whole age range | High |
| Fisher et<br>al., 2021 | UK<br>June 2020 | Qualitative<br>interviews | Purposive | Those connected with/following<br>University of Bristol email and<br>social media accounts and their<br>contacts | Age 12-17 years | 21 | Age 12-14 years = 52.38%<br>Age 15-17 years = 47.62%<br>Male = 52.38%<br>Female = 47.62% | Medium |
| Ganem et<br>al., 2023 | Spain<br>October 2021 and<br>January 2022 | Repeated<br>cross-sectional | Snowball | Students at 23 schools participating<br>in the COVID-19 Sentinel Schools<br>Network of Catalonia project | Age 3-18 years<br>Parents of aged 3-16 years<br>Subgroup analysed: 16-18<br>years) | 570 (16 and<br>over) | Age = 16-18 years | Medium |
| Ganczak et<br>al., 2021 | Poland<br>September 2020 | Intervention<br>(pre and post<br>surveys) |  | 25 classes in Polish high schools in<br>5 cities | Age 17-19 years<br>50% of participants enrolled<br>in life science programme | 518 | Mean age = 17.8, +/- 0.43 years<br>Age 17 years = 21.4%<br>Age 18 years = 77.4%<br>Age 19 years = 1.2%<br>Female = 66%<br>Enrolled in life science<br>programme = 72.6% | Medium |
| Garcia et al., 2021 | USA<br>July 2020 to January 2021 | Qualitative interviews | Convenience | Oregon State University Extension and 4_H listservs | Age 13-18 years, in grades 9-12<br>Parents of adolescents aged 13-18<br>Latinx/Latino/Hispanic<br>Able to speak, read and understand English or Spanish<br>Subgroup analysed: age 13-18 years | 24 | M = 15.92, SD = 1.18<br>Male = 29.17%<br>Female = 50%<br>Non-binary/trans = 20.83% | Medium |
| Garg et al., 2024 | USA<br>May 2023 | Cross-sectional | Convenience | Public High Schools in Springfield, Illinois | Age 13-19 years | 400 | M = 16.01 years<br>Male = 55%<br>Female = 45% | Low |
| Gewirtz-Meyden et al., 2022 | Israel<br>May to June 2021 | Cross-sectional | Not reported | Panel4All (online participant pool) | Age 12-18 years | 150 | Age range = 12-18 years<br>M = 15.3, SD = 1.9<br>Male = 37.2%<br>Female = 62.8% | Medium |
| Groenewald et al., 2023 | South Africa<br>January to December 2021 | Qualitative interviews | Snowball/Purposive | Participants in 'Life During Lockdown' study | Age 6-17<br>Adult caregivers<br>Subgroup analysed: 10-17 | 10 | Age = 10-17 years<br>Of age 12-17 years: | High |
|  |  |  |  |  | years |  | Male = 30% |  |
|  |  |  |  |  |  |  | Female = 70% |  |
| Guneyesu et al., 2023 | Turkey<br>April to September 2022 | Cross-sectional |  | Patients at Gazi University Medical Faculty Paediatric Emergency Department & Paediatric health & Diseases Polyclinic | Age 12-18 years | 924 | Age range = 11-18 years<br>M = 14.64 +/- 1.77 years<br>Male = nr<br>Female = 50.9% | Low |
| Hasanatulu dhhiyah et al., 2023 | Indonesia | Cross-sectional | Convenience | Students at 5 high schools in Surabaya or Sidoarjo | Not reported | 432 | Age = 15-17 years<br>Median age = 17 years, IQR = 15-17 | Medium |
| Hopfer et al., 2022 | USA<br>February to March 2021 | Mixed methods: Cross-sectional survey and focus groups (NB qualitative data only from parents) | Quota | Local schools, listservs, and Children's Hospital of Orange County | Age 11-18 years<br>Caregiver of 11–18-year-olds<br>Middle or high school student<br>Orange County resident<br>English or Spanish-speaking<br>Subgroup analysed: age 11-18 years | 46 | Age = 11-18 years<br>M= 14.1 years, SD = 1.7<br>Male = 57%<br>Female = 41%<br>Transgender = 2% | Quantitative: Low<br>Qualitative: No adolescent data |
| Hwang et al., 2023 | Taiwan<br>June to September 2022 | Cross-sectional | Not reported | Adolescents and caregivers in Kaohsiung | Vaccinated adolescents<br>Caregivers of vaccinated adolescents<br>Subgroup analysed: adolescents | 138 | Age range = 11-18 years (based on school grade)<br>M = 13.9, SD = 1.9<br>Male = 45.7%<br>Females = 54.3% | Medium |
| Inaba et al., 2022 | Japan / Russia<br>May to July 2021 | Cross-sectional | Purposive | Students in 8th and 9th grade in one high school in Niigata City, Japan and six secondary schools in Khabarovsk, Russia | Age 15 years | 721 | Age = 15 years<br>Male (Russia) = 169 = 23.44%<br>Female (Russia) = 225 = 31.21%<br>Male (Japan) = 176 = 24.41%<br>Female (Japan) = 151 = 20.94% | Medium |
| Janssen et al., 2023 | Curacao | Cross-sectional | Not reported | High-risk adolescent population of Curacao | Age 12-17 years | 116 | Age 12-15 years = 59.5%<br>Age 16-17 years = 40.5%<br>Male = nr<br>Female = nr | Low |
| Kajiwaru et al., 2022 | Japan<br>September 2021 | Cross-sectional | Not reported | Secondary data from web research company | Female<br>Age 15-49 years<br>Subgroup analysed: age 12-17 years | 250<br>(1013 total) | Age = 15-19 years<br>Female = 100% | Low |
| Kenworthy et al., 2022 | USA<br>September to November 2021 | Qualitative focus groups | Convenience | Three schools (2 high/1 middle school) which housed School Health Initiative clinics with which team had existing research relationships | Racial / ethnic minority students from low-income households only<br>Aged 16-18 years, staff member or caregiver of student aged 16-18 years<br>Able to speak, read and write English or Haitian Creole to 5<sup>th</sup> grade level<br>(Subgroup) | 6 | Age = 16-18 years | Medium |
| Klein et al., 2021* (see Middleman et al., 2021) | USA<br>August 2020 | Cross-sectional (1<sup>st</sup> wave of repeated cross-sectional) | “Representative groups” | Not reported | Age 13-18 years or caregiver of teen aged 13-18 years<br>Subgroup analysed: age 13-18 years | 300 | Age = 13-18 years | Low* |
| Lee et al., 2022 | Republic of Korea<br>June to July 2021 | Cross-sectional | Snowball | Students (and parents of students) at all elementary, middle and high schools | Age 12-17 years and their caregivers<br>Subgroup analysed: age 12-17 years | 272,914 | Age = 12-17 years<br>Grade:<br>6 = 18.7%<br>7 = 18.6%<br>8 = 15.3%<br>9 = 12.8%<br>10 = 17.4%<br>11 = 17.3%<br>Male = 44%<br>Female = 56% | Medium |
| Li et al., 2022(a) | China<br>August to October 2021 | Cross-sectional | Cluster | Students at one urban and one rural high school in each province: Jiangsu, Fujian, and Anhui in Eastern China | Age 12-17 years | 2048 | Age = 12-17 years<br>Mean age = 14.5 years<br>Aged 15-17 = 53.4%<br>Male = 48.8%<br>Female = Not reported | Low |
| Li et al., 2022(b) | China<br>August to October 2021 | Cross-sectional | Purposive | Two schools from three provinces | Age 12-17 years | 1847 | Age = 12-17 years<br>Male = 48.2%<br>Female = Not reported | Low |
| McKinnon et al., 2023 | Canada<br>January to March | Mixed methods: cross- | Convenience | Students in 2 public high schools | Age 14-17 years | 315 | Questionnaire:<br>Age 14 years = 17.1%<br>Age 15 years = 39.1%<br>Age 16 years = 29.5% | Medium |
|  | 2022 | sectional |  |  |  |  | Age 17 years = 13.3% |  |
|  |  |  |  |  |  |  | Male = 41.9% |  |
|  |  |  |  |  |  |  | Female = 56% |  |
|  |  |  |  |  |  |  | Other = 2.5% |  |
| McKinnon et al., 2023 | Canada<br>January to March 2022 | Mixed method: qualitative interviews | Convenience | Two ethnoculturally diverse, lower income neighbourhoods of Montreal | Age 14-17 years<br>Unvaccinated | 19 | Age 14-17 years<br>Mean age = 14.7 years<br>Male = 32%<br>Female = 68% | High |
| Mansfield et al., 2023 | USA<br>November to December 2021 | Qualitative interviews | Purposive | HCW practicing at KPSC hospitals & clinics in Southern California & their adolescent children | Vaccinated children of vaccinated healthcare workers<br>Adolescents: aged 12-17 years or caregivers of adolescents aged 12-17 years<br>Subgroup analysed: age 12-17 years | 17 | Age = 12-17 years<br>Male = 58.82%, Female = 41.18% | High |
| Meraya et al., 2022 | Saudi Arabia<br>July to September 2021 | Cross-sectional | Snowball | Residents of Saudi Arabia | Age 14+ years<br>Subgroup analysed: age 14-19 years | 870 | Age = 14-19 years<br>Male = Not reported<br>Female = 75% | Low |
| Middleman et al., 2021a | USA<br>Wave 1: August 2020 | Repeated cross-sectional (3 waves) | Online panel with "Diverse sample population" | Members of existing research panels | Age 13-18 years or caregiver of teen aged 13-18 years<br>Subgroup analysed: age 13- | 300 | Wave 1:<br>M = 15.3, SD = 1.7<br>Male = 44% | Medium |
|  | Wave 2: |  |  |  | 18 years |  | Female = 55% |  |
|  | February to March 2021 |  |  |  |  |  | Other = 1% |  |
|  | Wave 3: |  |  |  |  |  | Wave 2:<br>M = 15.1%, SD = 1.5 |  |
|  | June 2021 |  |  |  |  |  | Male = 55% |  |
|  |  |  |  |  |  |  | Female = 44% |  |
|  |  |  |  |  |  |  | Other = 1% |  |
|  |  |  |  |  |  |  | Wave 3:<br>M = 15.0, SD = 1.6 |  |
|  |  |  |  |  |  |  | Male = 51% |  |
|  |  |  |  |  |  |  | Female =48% |  |
|  |  |  |  |  |  |  | Other = 1% |  |
| Middleman et al., 2022* | USA | Repeated cross-sectional (3 waves) | “Nationally representative ” online panels | Members of existing research panels | Age 13-18 years or caregiver of aged 13-18 years | 300 | Not reported | Low* |
|  | Wave 1: August 2020 |  |  |  | Subgroup analysed: age 13-18 years |  |  |  |
|  | Wave 2: |  |  |  |  |  |  |  |
|  | February to March 2021 |  |  |  |  |  |  |  |
|  | Wave 3: |  |  |  |  |  |  |  |
|  | June 2021 |  |  |  |  |  |  |  |
| Moore et al., 2024 | USA | Cross-sectional | Quota | Adolescent members of Qualtrics internet panel | Age 15-17 years | 454 | Age 15 years = 14.3% | Low |
|  | September to October 2022 |  |  |  | English-speaking |  | Age 16 years = 35.9% |  |
|  |  |  |  |  |  |  | Age 17 years = 49.8% |  |
|  |  |  |  |  |  |  | Male = 36.3% |  |
|  |  |  |  |  |  |  | Female = 57.1% |  |
|  |  |  |  |  |  |  | Other = 6.6% |  |
| Nilsson et al., 2021 | Sweden<br>July to November 2020 | Mixed methods (cross-sectional with free text) | Convenience / snowball | Adolescents in Sweden | Age 15-19 years | 702 | Age 15-17 years = 71.5%<br>Age 18-19 years = 21.4%<br>Male = 42.2%<br>Female = 57.5% | Medium |
| Nilsson et al., 2021 | Sweden<br>July to November 2020 | Mixed method: Qualitative survey with free text | Convenience / snowball | Adolescents in Sweden | Age 15-19 years | 702 | Age 15-17 years = 71.5%<br>Age 18-19 years = 21.4%<br>Male = 42.2%<br>Female = 57.5% | Low |
| Oka et al., 2022 | Singapore<br>June to November 2021 | Cross-sectional | Cluster | Junior colleges and ITEs | Age 16-17 years | 460 | Age = 16-17 years<br>Mean age = 16.99 years, SD = 0.09<br>Male = 27.5%<br>Female 73% | Low |
| Parlak & Ener, 2022 | Turkey<br>October to November 2021 | Cross-sectional | Convenience | Paediatric outpatients of Kahta District State Hospital | Age 15-18 years<br>Paediatric outpatients | 303 | Age = 15-18 years<br>Male = 80.3%<br>Female = 19.7% | Medium |
| Persaud et al., 2023 | USA<br>May 2021 to | Mixed methods (cross- | Convenience | Patients in St Jude Sickle Cell Clinical Research and Intervention Program | Age 13-18 years with sickle cell disease (SCD) | 49 | Age = 13-18 years<br>Male = 49% | Low |
|  | February 2022 | sectional) |  |  | Caregivers of patients with SCD |  | Female = 51% |  |
|  |  |  |  |  | Unvaccinated |  |  |  |
|  |  |  |  |  | Subgroup analysed: age 13-18 years) |  |  |  |
| Persaud et al., 2023 | USA<br>May 2021 to February 2022 | Mixed methods (Qualitative free text) | Convenience | Patients in St Jude Sickle Cell Clinical Research and Intervention Program | Age 13-18 years with sickle cell disease<br>Caregivers of patients with SCD<br>Unvaccinated<br>Subgroup analysed: age 13-18 years | 49 | Age = 13-18 years<br>Male = 49%<br>Female = 51% | Low |
| Pimental et al., 2022 | Brazil<br>July to September 2021 | Cross-sectional | Snowball | Youth in Brazil | Age 14-19 years<br>Internet access<br>Living in 5 macro-regions of Brazil | 526 | Age = 14-19 years<br>Mean age = 16.9 years, SD = 1.6<br>Male = 32.3%<br>Female = 67.7% | Medium |
| Qian et al., 2023 | USA<br>Spring Semester 2022 | Cross-sectional | Cluster | Students in 9th to 12th grade, Madison West High School, Madison, Wisconsin | One of two cohorts in 10th grade Health class, 11th and 12th grade English class | 75 | Age = 10th -12th grade (15-19* years, age not provided)<br>Female = 54.7%, Male = 38.7%<br>Non-binary = 5.3%<br>No Answer = 1.3% | Low |
| Ramaiya et al., 2023** | Belgium | Qualitative focus groups | Purposive/convenience | Urban poor adolescent populations in Ghent, Sao Paolo, Shanghai, Kinshasa, Semarang and Denpasar, | Age 13-18 years (except in China – aged 15-18 years) | 237 | Age = 13-18 years | Medium |
|  | Brazil |  | (China) | Blantyre and New Orleans | Residing in a poor, urban setting |  | Mean age = Not reported |  |
|  | China |  |  |  |  |  | Male = Not reported |  |
|  | DR Congo |  |  |  | Member of Global Early Adolescent Study cohort (except in Brazil & China) |  | Female = Not reported |  |
|  | Indonesia |  |  |  |  |  |  |  |
|  | Malawi |  |  |  |  |  |  |  |
|  | USA |  |  |  |  |  |  |  |
|  | March 2021 to April 2022 |  |  |  |  |  |  |  |
| Rehati et al., 2022 | China | Cross-sectional | Cluster | Pupils in grades 7-12 in middle and upper school students in four cities in China | Age 12-17 years | 9153 | Age = 12-17.5 years | High |
|  | December 2020 |  |  |  | Able to read & complete survey independently |  | Female = 50%<br>Male = 50% |  |
| Rogers et al., 2021 | USA | Cross-sectional | Stratified random | Qualtrics research database | Age 12-17 years | 916 | Age = 12-17 years<br>Mean age = 14.69 years, SD = 1.69<br>Male = 47.9%, Female = 47.3%, Transgender/Non-binary = 3% | Medium |
|  | June 2021 |  |  |  |  |  |  |  |
| Rosen et al 2022* | USA | Cross-sectional | Convenience | Patients at a large Midwest medical centre | Age 12-25 years | 248 | Age = 12-17 years<br>Mean = 14.8 years, SD = 1.8 | Low* |
|  | January 2021 |  |  |  | Subgroup analysed: age 12-17 years |  |  |  |
| Rosen et | USA | Cross- | Convenience | Patients at large Midwest medical | Adolescent (12-17 years) | 242 | Age 12-17 years | Medium |
| al., 2024 | January 2021 | sectional |  | centre | Young adult (18-25 years) |  | Mean age = 14.8 years, SD = 1.8 |  |
|  |  |  |  |  | Or parent with a child 17< years |  | Male = 36.4% |  |
|  |  |  |  |  | Subgroup analysed: age 12-17 years |  | Female = 59.1% |  |
| Rothoeft et al., 2023 | Germany<br>July to September 2021 | Cross-sectional | Convenience | Patients at Siegen vaccination centre after receiving vaccination | Age at least 12 years | 1378 | Age 12-13 years = 34.8% | Medium |
|  |  |  |  |  | Student in Siegen |  | Age 14-15 years = 37.4% |  |
|  |  |  |  |  | Receiving vaccination |  | Age 16-17 years = 27.8% |  |
|  |  |  |  |  | Participating adolescent or parent of participating adolescent |  | Mean age = 14.4 years, SD = 1.6 |  |
|  |  |  |  |  | Voluntary participation in survey |  | Male = 47.5% |  |
|  |  |  |  |  | No contraindications to vaccine |  | Female = 49.4% |  |
|  |  |  |  |  | Subgroup analysed: age 12-17 years |  | Missing = 0.3% |  |
|  |  |  |  |  |  |  | Other = 0.9% |  |
| Roy et al., 2023 | Bangladesh<br>April to August 2022 | Cross-sectional | Not reported | Higher secondary students in rural areas | XI or XII grade | 1514 | Aged = 16-18 years | Medium |
|  |  |  |  |  | Age 16-18 years |  | Age 16 years = 23.6% |  |
|  |  |  |  |  |  |  | Age 17 years = 49.9% |  |
|  |  |  |  |  |  |  | Age 18 years = 26.5% |  |
|  |  |  |  |  |  |  | Male = 46.5% |  |
|  |  |  |  |  |  |  | Female = 53.5% |  |
| Ryan et al., 2023 | USA<br>April 2021 | Cross-sectional | Quota-based | Online survey panel | Age 13-17 years<br>Unvaccinated | 831 | Age = 13-17 years | Medium |
| Scherer et al., 2021 | USA<br>April 2021 | Cross-sectional | Non-probability based | Online survey panel | Age 13-17 years or<br>Caregivers of 12–17-year-olds<br>Living in USA<br>Unvaccinated<br>Subgroup analysed: age 13-17 years | 985 | Age 13-17 years<br>Age 13-15 years = 40.4%<br>Age 16-17 years = 59.6%<br>Female = 58.8%, male = 36.4% | Low |
| Tirawi et al., 2022 | India<br>Date: Not reported | Cross-sectional | Convenience | Youth attending Covid vaccination centre in New Delhi | Age 15-17 years | 947 | Aged = 15-17 years<br>Age 15 years = 32.95%<br>Age 16 years = 32.63%<br>Age 17 years = 34.42%<br>Male = 54.59%<br>Female = 45.41% | Low |
| Tu et al., 2022 | USA<br>October to November 2021 | Cross-sectional | Quota-based | Members of online survey panel | Age 13-17 years | 439 | Age = 13-17 years<br>Age 13 - 15 years = 40.32%<br>Age 16-17 years = 59.68%<br>Male = 41.46%,<br>Female = 52.85%<br>Other/prefer not to answer = | Medium |
|  |  |  |  |  |  |  | 5.6% |  |
| Unger et al., 2023 | USA<br>May to June 2022 | Cross-sectional | Cluster | High school students in predominantly Hispanic neighbourhood of Los Angeles County, California | In Grades 9-12<br>Age 14-18 years<br>Able to read and write in English or Spanish | 444 | Age = 14-18 years<br>Age 14 years = 16%<br>Age 15 years = 34%<br>Age 16 years = 22%<br>Age 17 = 16%<br>Age 18 years = 12%<br>Mean age = 15.74 years, SD = 1.25<br>Male = 41%<br>Female = 55%<br>Other = 4% | High |
| Uroko & Nche, 2023 | Nigeria<br>September to October 2021 | Qualitative interviews | Convenience | Students in primary or secondary school in Benue, Delta, Enugu or Lagos states | In final class/grade at school<br>Able to express self | 20 | Age = 11-19 years<br>M = 14.7 years<br>Male = 70%<br>Female = 30% | Medium |
| Wang et al., 2022 | Burkina Faso, Ethiopia, Ghana, Nigeria, Tanzania<br>July to December 2021 | Cross-sectional | Purposive | 300 households in each region, from existing HDSS or national surveys | Age 10-19 years | 2662 | Age = 10-19 years<br>Mean age = 15.5 years, SD = 2.5<br>Male = Not reported<br>Female = 52.8% | Medium |
| Wang et al., |  | Cross-sectional |  |  |  |  |  |  |
| Wirunpan et al., 2021 | Thailand<br>May 2021 | Cross-sectional | Cluster | Pupils at Satit Prasarnmit International Programme in Bangkok | Age 14-18 years | 136 | Age = 14-18 years<br>Age 14-15 years = 11.8%<br>Age 16-17 years = 36.8%<br>Age 15-16 years = 26.5%<br>Age 17-18 years = 25%<br>Male = 22.1%<br>Female = 77.9% | Low |
| Wong et al., 2022 | Hong Kong<br>June 2021 | Cross-sectional | Convenience | 33 secondary schools | Age 12-18 years | 2609 | Median age = 14 years<br>Male = 55.1%<br>Female = 44.9% | Medium |
| Wong et al., 2022<br>(Brief report of above) | Hong Kong<br>June 2021 | Cross-sectional | Convenience | 33 secondary schools | Age 12-18 years | 2609 | Not reported | Low* |
| Zhang et al., 2022 | China<br>March to April 2021 | Cross-sectional | Snowball | Adolescents using WeChat online in China | Age 16-21 years<br>Subgroup analysed: age 16-17 years | 1009 | Age = 16-17 years<br>Mean age = 16.46 years, +/- 0.63<br>Male = 47% | Medium |
| Female = Not reported |  |  |  |  |  |  |  |  |
| Zilhadia et al., 2022 | Indonesia<br>August 2021 | Cross-sectional | Convenience | Muslim students at secondary high school in Jakarta region | In grade 11 or 12 (age 14-18 years)<br>Muslim | 440 | Age = 14-18 years<br>Age 14 years = 0.2%<br>Age 15 years = 7.7%<br>Age 16 years = 40.5%<br>Age 17 years = 46.1%<br>Age 18 years = 5.5%<br>M = 16.49 years, SD = Not reported<br>Male = 26.4%<br>Female = 73.6% | Medium |

### 3.2 Quality assessment

Overall, the quality of the included studies was mixed (Appendix B). Only five quantitative ^33–37^ and three qualitative studies ^38–40^ were assessed as high quality. Twenty-three quantitative ^41–63^ and four qualitative studies ^54, 64–66^ were assessed as low quality, although three of those assessed as low quality were conference abstracts and therefore lacked detailed data. The remainder of studies were assessed as medium quality. Two studies were not peer-reviewed ^31, 32^. Scores on the amended NIH cohort and cross-sectional tool ^24^ ranged between two and 12. Notably, only nine studies used validated, reliable measures for all predictors, and seven for outcomes. The assessed quality of each study is included in Table 1, and the full quality assessment is included at Appendix B. Qualitative study scores using CASP ^25^ ranged from four to 12. All studies were included in this review regardless of quality, but we have incorporated quality assessments when considering the strength of evidence for each predictor.

### 3.3 Psychological factors related to attitudes, intention or uptake of Covid-19 vaccines

#### 3.3.1 Vaccine attitudes

Data on the predictors of vaccine attitudes was limited: of the included studies, only five reported on vaccine attitudes as an outcome, investigating different attitudes. One low-quality study found that vaccine cost and accessibility was significantly positively associated with a positive attitude toward the vaccine ^43^, defining a positive attitude as the likelihood of recommending the Covid-19 vaccine to others.

#### 3.3.2 Vaccine uptake and intention

Table 2 provides a summary of psychological factors associated with intention or uptake, and corresponding qualitative data, categorized against COM-B components ^18^. Table 3 provides a summary of top three reasons for vaccine acceptance and vaccine hesitancy reported by cross-sectional studies. A summary of results is narratively reported. For full results, see appendix C.

**Table 2.**
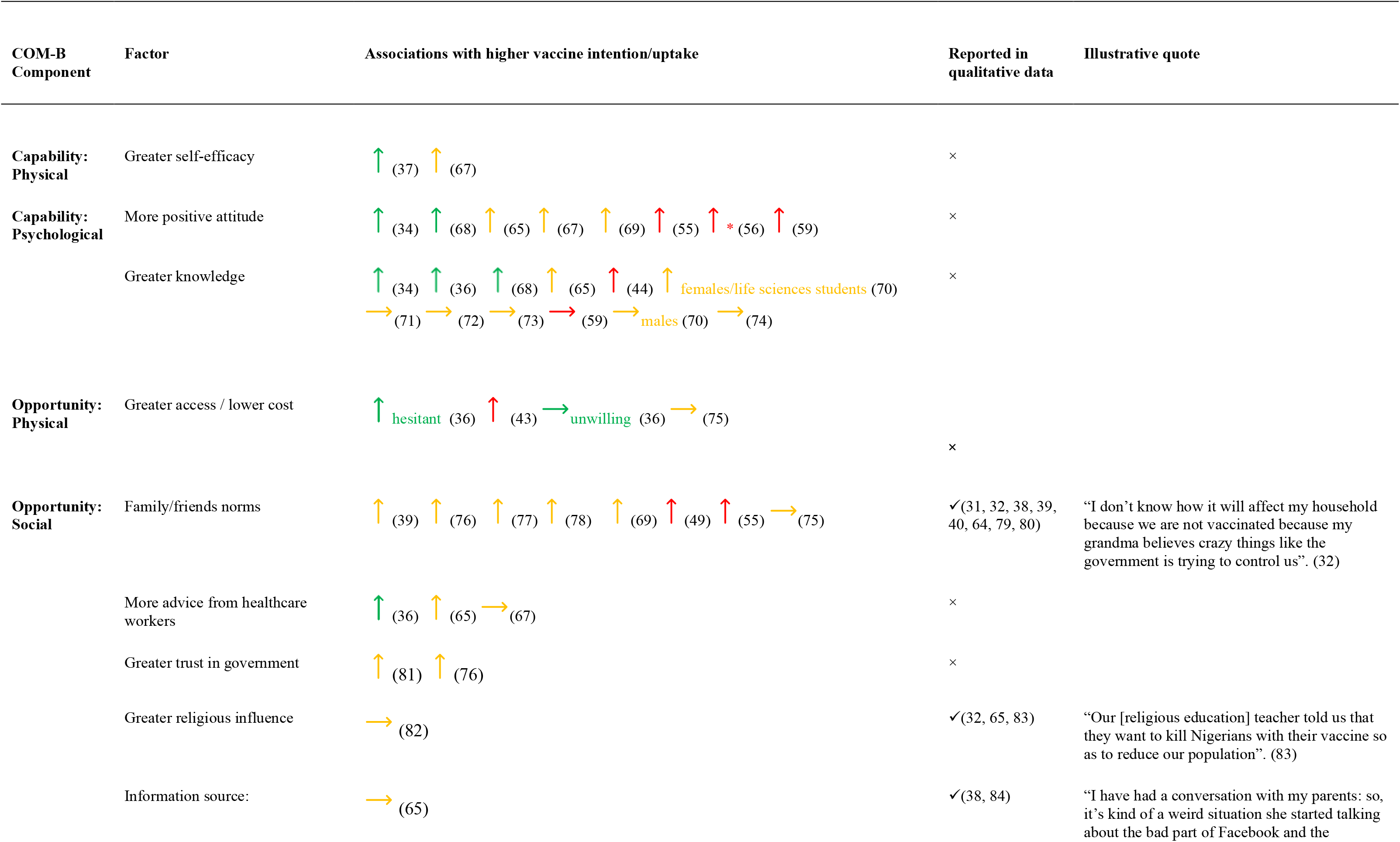
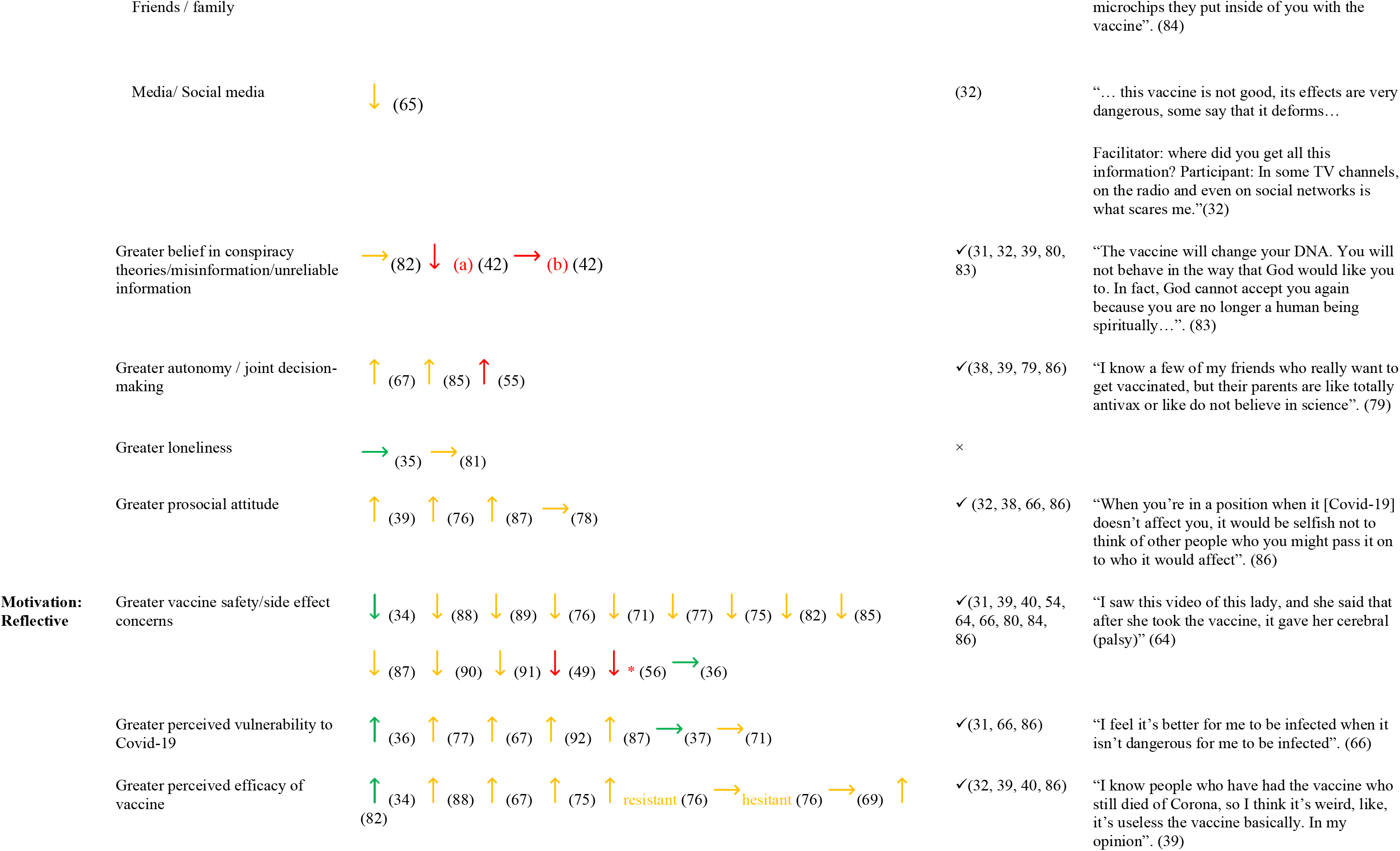
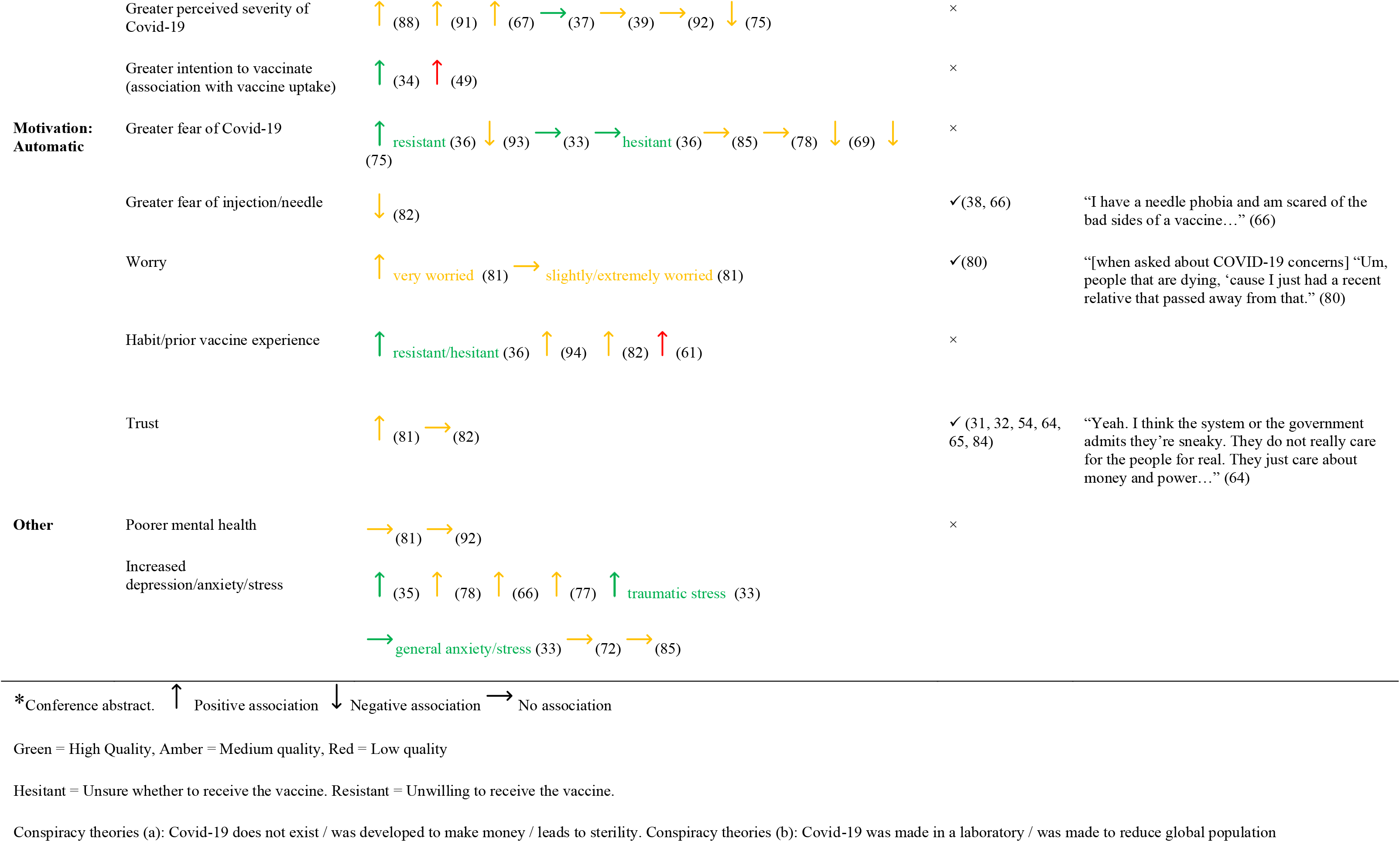
Factors associated with vaccine intention or uptake and qualitative results classified by COM-B model components. [To be printed in colour]

**Table 3.** Summary of studies reporting quantitative data on reasons for/against Covid-19 vaccination, top three reasons.

| Citation | Reasons to be vaccine acceptant (descriptive data)<br>*based on graph estimates | Reasons to be vaccine hesitant (descriptive data)<br>*based on graph estimates |
| --- | --- | --- |
| **Afifi et al., 2021** | n/a | Worried that vaccine is not safe (65.9%)<br>Don't know enough about vaccine (60.9%)<br>Do not think it will work (23.9%) |
| **Akhtar et al., 2023** | Persuaded by parents (36.1%)<br>Friends getting vaccinated (27.7%)<br>Doctors' advice (16.6%) | Fear of side effects of vaccination (65.2%)<br>Allergies (22.2%)<br>No benefit from vaccination (16.6%) |
| **Assad et al., 2023** | n/a | Insufficient information (31%)<br>Misinformation & rumours (26%)<br>Vaccine not safe (18%) |
| **Ates et al., 2023** | n/a | Fear of side effects (36.3%)<br>Uncertainty about effect of vaccine/insufficient studies (21.7%)<br>No valid reason to not get vaccinated (40.1%) |
| **Bergh et al., 2023** | Covid-19 would be a serious illness for people in my community (68.5%)<br>Most of my family think getting the vaccine is a good thing (58.3%)<br>Vaccine will protect me from becoming very sick (56.7%) | Fear of vaccine needle (58.0%)<br>Vaccine site too far away (26.4%)<br>Vaccine centre waiting times too long (22.3%) |
| **Bhowmick et al., 2022** | Recommended by government (22.7%)<br>Seeing many people get infected with Covid (21.5%)<br>Scared about getting Covid-19 (19%) | n/a |
| **Bracko & Simon, 2022** | Self-protection and protection of others (12%)<br>Contribution to herd immunity (7.6%)<br>General trust in vaccine(s) (2.4%) | Vaccine safety uncertainties (50.2%)<br>No interest in getting vaccinated (2.8%)<br>No need to do so because of young age, strong |
|  |  | immune system or having already had the disease (2.4%) |
| **Can & Kurtulus, 2021** | Fear of transmitting virus to family and loved ones (46.2%)<br>Threat of pandemic (8.6%)<br>Fear of disease and death (7.9%) | Side effects (30%)<br>Not believing effectiveness of vaccine (5.9%)<br>Feeling like a guinea pig (3.4%) |
| **Cupertino et al., 2022** | To protect the general population (46.9%)<br>To return to a normal life (31.0%) | Influenced by parents and family's decisions (65.8%)<br>Fear of side effects (40.4%)<br>Parent decision to not be immunized (23.7%) |
| **Das et al., 2024** | Vaccine will protect me (65.45%)<br>Vaccine is generally safe (44.54%)<br>Vaccines will help control the pandemic (55.44%) | Vaccines are unsafe (12.73%) – Due to side effects (100%)<br>Unvaccinated/unwilling to be vaccinated in future:<br>Fear of side effects (50%)<br>Considered vaccine a hoax (50%) |
| **Dhanker et al., 2023** | Fear of getting COVID-19 infection (50.5%)<br>family recommendation (34.8%)<br>Government recommendation (33.2%) | Vaccine might produce serious side effects (35.7%)<br>COVID-19 vaccine is not safe (2.8%) |
| **Efendi et al., 2022** | n/a | Vaccines 1/2:<br>Parents forbade them (49.6%) / (57.5%)<br>Fear of side effects (22.9%) / (19.5%)<br>Efficacy doubts (15.1%) / (10.8%) |
| **Euser et al., 2022** | Willing/Non-willing:<br>I want to protect myself (81%)/(41%)<br>I want to protect others (79%/40%)<br>I want to help ending the corona crisis (67%/42%) | Not willing / hesitant:<br>I am afraid of unknown long-term effects of the vaccine 73% / 56% I do not know enough about the vaccine yet 38% / 45%<br>I am afraid of side effects 38% / 41% |
| **Ganem et al., 2023** | n/a | **October 2021:**<br>Concern with the time to develop the vaccine (63.6%)<br>Concern about side effects (50.9%)<br>Necessity for more information before deciding to vaccinate (45.5%).<br>**January 2022:**<br>Concern about side effects (68.4%)<br>Time to develop the vaccine (63.2%)<br>Previous COVID-19 disease (42.1%) |
| **Garg et al., 2024** | To protect self (78%)<br>To protect family/friends (74%)<br>To protect community (25%) | Do not trust vaccine (57%)<br>Parent/guardian decision (39%)<br>Question efficacy (21%) |
| **Gewirtz-Meyden et al., 2022** | n/a | Do not trust drug companies to make sure vaccine will be safe (56.1%)<br>Virus not dangerous (53.7%)<br>Do not believe in the safety of the vaccine in the short term (51.2%) |
| **Guneyasu et al., 2023** | If I get sick, I will have a lighter disease course (46.5%)<br>I fulfil my social responsibility (25.8%)<br>I will not get infected again (12.2%) | Due to vaccine side effects (5.8%)<br>Distrust of vaccine content (2.6%)<br>Don't think the vaccine will work (2.5%) |
| **Hopfer et al., 2022** | Confidence in vaccine safety - 99<br>Social benefit -89<br>Confidence in vaccine efficacy -82<br>*NB Ranking scores not %ages* | Concerns over potential long term side effects - 128<br>Concerns over potential short term side effects or reactions - 99<br>Concerns over how well the vaccine works - 59<br>*NB Ranking scores not %ages* |
| **Janssen et al., 2023** | n/a | Aged 12-15 years:<br>Do not trust vaccine (28%)<br>Admitted to hospital (4%)<br>Forgot (4%)<br>Aged 16-17 years:<br>Fear of needles (4%)<br>Do not trust vaccine (13%)<br>Taking post-op medication (4%) |
| **Kajiwara et al., 2022** | n/a | Would not get vaccinated / would not accept 3rd dose:<br>I would be concerned about side effects from the vaccine (29.5%) / (33.3%)<br>I am afraid/anxious (25%) / (0%)<br>I do not like needles/injections (20.5%) / 0% |
| **Lee et al., 2022** | Prevent infection for oneself (62%*)<br>Prevent transmission family and friends (58%*)<br>Return to normal lifestyle (30%*) | Concerns for safety issues (90%*)<br>Do not want interruptions in school and studies (30%*)<br>Low risk of infection (20%*)<br>Public health measures sufficient for prevention (20%*) |
| **Li et al., 2022a** | Confidence in vaccine effectiveness (in vaccinated) 95% (in not vaccinated) 93.6%<br>Totally own decision, no specific reason (70.6%) | Worry about vaccine safety (in vaccinated) 15.5%, in non-vaccinated (25.8%)<br>Willing to take, but declined by vaccination staff 6 (20.0) |
|  | Life is affected by Covid-19 - seriously affected/affected but not seriously (in vaccinated) 64.1% (in non-vaccinated 58.1%) | Worry about the safety because of own chronic diseases 5 (16.7) |
| **McKinnon et al., 2023** | Vaccinated agree with following statements:<br>By being vaccinated against COVID-19, I am helping to protect the health of others in my community (77.6%)<br>Most of my friends and family members are vaccinated against COVID-19 (82.7%)<br>Vaccines are an effective way to reduce the risk of contracting COVID-19 (60.6%) | Unvaccinated - agree with following statements:<br>Physical distancing, frequent handwashing and wearing a mask are enough to protect me from COVID-19 (48.1%)<br>By being vaccinated against COVID-19, I am helping to protect the health of others in my community (37.2%)<br>I distrust COVID-19 vaccines because they were developed too quickly (44.3%)<br>Vaccines against COVID-19 are safe (10%) |
| **Middleman et al., 2022a** | Agree / strongly agree: Wave 1/2/3:<br>I want to protect myself (53% / 49% / 51%)<br>I want to protect everyone in my family (47% / 52% / 44%)<br>Vaccination is the best way for me to avoid a potentially serious disease (37%/38% /38%) | %age choosing agree / strongly agree:<br>I have some concerns about the safety of vaccines - 56% / 64% / 62%<br>I have some concerns about the effectiveness of some vaccines - 55% / 64% / 63%<br>What I have read on social media has concerned me about the safety of some vaccines - 50% 56% / 57%<br>Wave 1 / 2 / 3:<br>Concerned about possible side effects - 40% / 47% / 58%<br>Concerned I could get Covid from the vaccine - 23% / 20% / 15%<br>I do not think the Covid vaccine will work well - 17% / 20% / 15% |
| **Moore et al., 2024** | *To protect self (80.8%)<br>*To protect others (81.3%)<br>*I want to return to normal life (11.5%)<br>*Ranked importance 1, 2 3 respectively | *We do not know about long-term effects of vaccine (57.7%)<br>*Concern about side effects (56.6%)<br>*I do not like needles/shots (45.2%)<br>*Ranked importance 1, 2 3 respectively<br>Biggest barriers<br>Parent preference (43.3%)<br>I had little or no free time to get vaccinated (38.73%)<br>Few or not vaccination sites available (13.3%) |
| **Oka et al., 2022** | n/a | Long term risk to health (38.7%)<br>Vaccine efficacy (37.4%)<br>Fear turning into a carrier (21.3%)<br>Feel it is not necessary (17.0%) |
| **Parlak & Ener, 2022** | n/a | Concerns about adverse effects of vaccines (25.7%)<br>Negative comments of others on vaccines (18.8%)<br>I think the COVID-19 vaccine is ineffective (18.2%) |
| **Persaud et al., 2023** | n/a | Lack of personal utility/benefit (53%)<br>Mistrust / misinformation / rushed (40%)<br>Unanswered medical questions (7%) |
| **Rehati et al., 2022** | n/a | Major concerns that affect my Covid-19 vaccination decision:<br>Resistant / Hesitant / Willing:<br>Safety (8% / 31.3% / 60.7%)<br>Effectiveness (7.5% / 30.7% / 61.8%)<br>Price (6.1% / 31.3% / 62.7%) |
| **Rogers et al., 2021** | n/a | Frequency counts for vaccine concern items on 5-point scale:<br>Vaccine not tested enough (M = 3.12, SD = 1.38)<br>Worried about side effects (M= 3.32, SD = 1.34)<br>Will wait for others to get the vaccine (M = 2.95, SD = 1.47) |
| **Rosen et al., 2024** |  |  |
| **Rothoeft et al., 2023** | Concern about contracting Covid-19 (82.3%)<br>Access to leisure facilities (76.9%)<br>Protecting family members (70.9%) |  |
| **Roy et al., 2023** | Trust (81.37%)<br>Culture (usual habit) (79.4%)<br>Belief in vaccine efficacy (75.63%) | Negative information (79.99%)<br>Injection anxiety (70.28%)<br>Fear of side effects (60.97%) |
| **Ryan et al., 2023** | Protecting the health of family and friends (87.2%)<br>Personal Covid-19 prevention (77.3%)<br>Protecting the health of communities (75.6%) | Concernn about side effects (45%)<br>Waiting to see if it is safe (43.9%)<br>My parents/caregivers will decide for me (36.7%) |
| **Scherer et al., 2021** | Factors which would increase intention (among unvaccinated):<br>If vaccination was a school requirement (23.9%)<br>More information about safety (21.7%)<br>More information about efficacy (17.6%) | n/a |
| **Tirawi et al., 2022** | Motivation to have vaccine:<br>Parents (69.8%)<br>Teachers (24.08%)<br>Friends (4.86%) | n/a |
| **Tu et al., 2022** | Reasons for being vaccinated:<br>To protect family and friends (44.3%)<br>To protect self (43.1%)<br>To socialise / go out freely (25.6%)<br>Factors which would encourage hesitant to be vaccinated:<br>Evidence showing efficacy (17.1%)<br>School mandates (10.7%)<br>More information about safety (6.1%) | n/a |
| **Wang et al., 2022a** | To keep self and family safe (94.3%)<br>Parents'/family's will (71.7%)<br>Doctor suggested (63.1%) | Perceived low necessity (46.7%)<br>Concerns about effectiveness (11.2%)<br>Concerns about safety (45%) |
| **Wirunpan et al., 2021** | n/a | Concern over the unknown side effects (24.3%)<br>Lack of confidence in government (16.9%)<br>Personal reasons (11.8%) |
| **Wong et al., 2022a** | Worried to be infected (59%)<br>To protect family (53%)<br>To return to the normality before COVID-19 (42%) | Concerned about vaccine safety (79%)<br>Concerned about vaccine efficacy (52%)<br>Facemasks and social distancing are sufficient (26%) |
| **Zhang et al., 2022** | Reduce the risk of infection (66.3%)<br>Vaccine has been proved to be safe (56.6%)<br>Low risk of side effects from the vaccine (51.2%) | Concerns over side effects (70.5%)<br>No risk of infection (34.9%)<br>Unpleasant vaccination experience (19.3%) |

##### Capability

###### i) Physical

One high- ^37^ and one medium-quality study ^67^ reported that higher self-efficacy was significantly associated with higher vaccine acceptance. No qualitative studies reported themes pertaining to physical capability.

###### **ii)** Psychological

We found strong evidence that a positive attitude toward vaccines was a reason to be vaccinated, reported in 12 quantitative studies ^39, 43, 45, 46, 50, 58, 68–73^. Furthermore, eight studies found those with a positive vaccine attitude were significantly more likely to accept it ^34, 55, 56, 59, 65, 67, 74, 75^. However, vaccine attitude measures were heterogenous. Evidence relating to vaccine knowledge was mixed. Three high-quality ^34, 36, 75^, one medium- ^65^ and one low-quality quantitative study ^44^, and one medium-quality intervention study reported higher vaccine knowledge was associated with higher vaccine acceptance. However, five studies ^59, 76–79^ of medium or low quality reported no significant association.

##### Opportunity

###### i) Physical

There was moderate evidence that physical opportunity impacted vaccine behaviour. Low vaccine cost was a reason to accept the vaccine in four studies and higher vaccine or travel cost a reason not to accept the vaccine in two studies (Table 2). Increased time, accessibility, and convenience were reasons to accept a vaccine in eight studies and inconvenience, or lack of time were reasons not to accept a vaccine in two studies ^63, 80^ . One high- ^36^ and one low-quality ^43^ study found that lower cost and increased accessibility increased acceptance in those who were vaccine hesitant (undecided), but not for those who were vaccine resistant (unwilling). On the other hand, one medium-quality study found no association between increased convenience and vaccine acceptance ^80^.

###### **ii)** Social

There was strong evidence that family and/or friend norms influenced participants’ vaccine behaviour, reported in quantitative and mixed methods studies (n=22, Table 2). Five qualitative studies supported this finding, with adolescents tending to have the same vaccination status as their in-group (family or friends) and reassuring peers on their vaccine experience ^31, 38, 64, 81, 82^. Seven studies of medium ^39, 73, 74, 83, 84^ or low-quality ^49, 55^ found friend or family vaccine acceptance were significantly associated with adolescent vaccine acceptance. However, one medium-quality study reported no significant association between friend and family norms and vaccine acceptance ^80^. No high-quality studies investigated associations between family or friend norms and vaccine behaviour.

Advice from healthcare workers was a further influence, reported in eight quantitative or mixed methods studies as a reason to vaccinate (Table 2). In one medium and one high-quality study, receiving advice and information from healthcare workers was associated with increased vaccine acceptance ^36, 65^ but a medium-quality study found no significant association ^67^. The state was less influential than family or friends. Two medium-quality studies ^73, 85^ reported that greater trust in government advice was associated with greater vaccine acceptance This was supported by two qualitative studies ^64, 81^ in which adolescents reported being vaccine hesitant due to lack of trust in the government and medical institutions. Religion or cultural advice or norms were reasons to accept or decline vaccines in six quantitative studies (Table 2). On the other hand, religious beliefs were not found to be significantly associated with vaccine acceptance in one medium-quality quantitative study ^86^.Two qualitative studies ^32, 87^ reported adolescents repeating the words or practices of cultural or religious leaders e.g., taking herbal remedies or believing the vaccine to be “ungodly”, but specifically in cultures in which religion was more dominant. Teacher influence was a reason to accept or decline a vaccine in three low-quality quantitative studies (Table 2) and one medium-quality qualitative study ^88^, but was not measured elsewhere. Qualitative reports showed that the mechanism of social influence was also important. While being influenced by others, adolescents were resistant to vaccine instructions or mandates from government, healthcare workers or parents/families ^39^ and also resisted instructing others ^38^.

Autonomy and choice were important aspects of social influence. One quantitative study reported that making one’s own decision was a reason to accept the vaccine, and four studies reported parent refusal on their behalf as a reason not to accept the vaccine (Table 2). Two medium- and one low-quality quantitative studies found more collaborative parent-adolescent decision making ^55, 89^ or increased perceived behavioural control ^67^ to be significantly associated with increased acceptance. Qualitative studies detailed a range of decision-making processes including false autonomy (e.g., “they were going to make me do it anyway” ^38^.

Adolescents were more confident in the vaccine than their hesitant parents ^38^ and saw benefits to adolescent peers being in control of vaccine decisions, ^82, 90^ e.g., “I know a few of my friends who really want to get vaccinated, but their parents are like totally anti-vax or like do not believe in science” ^82^.

Although a lack of credible information (n = 11) and belief in conspiracy theories (n = 10) were reasons not to be vaccinated (Table 2), social media was not a wholly negative influence on vaccine acceptance. A low-quality quantitative study reported that greater belief in some conspiracy theories were associated with decreased vaccine intention or uptake, whereby those who believed that Covid-19 did not exist, was developed to make money or leads to sterility were less likely to receive a vaccine ^42^ (Table 2). One high- and two medium-quality studies reported that greater use of social media ^35, 65, 91^ was associated with lower vaccine acceptance. On the other hand, one medium-quality study reported no association between belief in conspiracy theories and acceptance ^86^, two medium-quality quantitative studies reported greater use of Facebook and Instagram (but not other social media platforms) ^89^ and greater use of overall media ^72^ to be significantly associated with increased vaccine acceptance. Furthermore, in two qualitative studies ^32, 81^ participants saw older relatives as those more likely to believe in and share conspiracy theories, and took on the role of “debunkers” of the misinformation to which their older relatives were exposed.

Finally, overall social connection and prosocial behaviour were reasons to accept a vaccine. While self-protection (n = 10) was a reason to accept the vaccine, a desire to protect others (n = 16) was more commonly stated as a reason to be vaccinated (Table 2). Quantitative studies (n = 12) reported a desire to end the crisis and resume normal life were reasons to be vaccinated, and in two studies this was specifically to support the economy, society and contribute to herd immunity (Table 2). Three high- or medium-quality quantitative studies found a greater desire to protect others was significantly associated with increased vaccine acceptance ^39, 73, 92^, although while one study reported no significant association between prosocial behaviour and vaccine behaviour ^84^. Qualitative results also showed vaccine acceptance was motivated by both the desire to return to a normal life ^31, 81, 90^ but also to alleviate their families’ suffering ^81^ and for the benefit of society ^38, 66^. Those who felt more a part of their school community were significantly more likely to accept the vaccine in one high quality study ^35^, although loneliness was not significantly associated with vaccine acceptance in either a high- or medium-quality study ^35, 85^.

##### Motivation

###### i) Reflective

Vaccine safety (n = 17) and side effects (n = 25) concerns were the most common reasons not to accept the vaccine in quantitative studies. Equally, confidence in the safety of the vaccines (n = 9) was a reason to accept the vaccine (Table 2). Those with greater safety or side effect concerns were significantly less likely to be vaccine acceptant in one high-, 11 medium- and two low-quality studies ^34, 49, 56, 69, 72, 73, 76, 80, 83, 86, 89, 92–94^, although as an exception one high quality study found no significant association between vaccine safety concerns and vaccine acceptance ^36^. These concerns related to the speed and perceived lack of rigour in vaccine development, reported in five quantitative studies (Table 2). Qualitative studies also reported vaccine safety ^31^ and side effect concerns ^64, 66, 87, 90^ often as part of a broader mistrust of state, pharmaceutical and medical authorities, especially among racially diverse and or more deprived communities.

Adolescents evaluated the necessity of the vaccines based on their perceptions of susceptibility to Covid-19, vaccine efficacy, and severity of Covid-19. There was strong evidence that adolescents did not consider themselves susceptible to Covid-19, and this was related to lower vaccine intention or uptake. Perceived lack of susceptibility was a reason not to accept the vaccine in quantitative (n = 19, Table 2) and qualitative (n = 1) ^31^ studies. Six quantitative studies found the more an adolescent perceived themselves susceptible to Covid- 19, the greater their vaccine acceptance ^36, 80, 83, 95–97^. However, one high- and one medium-quality study found no significant association between perceived susceptibility to Covid-19 and vaccine intention or uptake ^37, 76^.

Perception the vaccine was ineffective (n = 27) and preference for “natural immunity” (n = 7) were reasons not to accept the vaccine in quantitative studies (Table 2). In six studies, the more participants viewed the vaccine as effective the more likely they were to be vaccine acceptant ^34, 67, 69, 73, 80, 86^, although one medium-quality study found no significant association ^74^. In three medium-quality studies, the more participants viewed Covid-19 as severe, the more likely they were to be vaccine acceptant ^67, 69, 93^. However, one high- and two medium-quality studies found no evidence for an association between perceived severity and vaccine acceptance ^37, 39, 95^. Finally, there was moderate evidence in one high- and one low-quality study that those with a greater intention to vaccinate were more likely to receive a vaccination ^34, 48^.

###### **ii)** Automatic

While worry or fear of Covid-19 (n = 5) were reasons to accept the vaccine (Table 2), fear of needles/injections was conversely a reason *not* to accept the vaccine in both quantitative (n = 9, Table 2) and one qualitative study ^66^. Two medium-quality studies found increased fear of Covid-19 was associated with lower vaccine acceptance ^71, 74^. However, one high-quality and three medium-quality studies found no significant association between worry or fear of Covid-19 and vaccine acceptance ^33, 80, 84, 89^. One medium-quality study found that greater fear of needles was significantly associated with lower vaccine acceptance ^80^.

While having a negative prior vaccine experience was a reason not to accept a vaccine in quantitative studies (n = 3, Table 2), one high- ^36^, two medium- ^86, 98^ and one low-quality study ^61^ reported that those who had previously received a vaccine were significantly more likely to accept the Covid-19 vaccine.

Finally, there was moderate evidence that trust influences vaccine behaviour. Trust in vaccines (n = 3) and in following government advice (n = 4) were reasons to accept vaccines (Table 2). Likewise, distrust in vaccines, governments, and/or pharmaceutical organisations (n = 15) were reasons *not* to accept vaccines. Two medium-quality studies investigated associations between trust and vaccine behaviour: while one ^85^ found that the greater the trust in government, the more likely participants were to accept the vaccine, the other ^86^ found no evidence for an association.

##### Other

There was mixed evidence relating to the impact of mental health conditions on vaccine acceptance, with heterogenous mental health conditions investigated. Two medium-quality studies found no significant association between poorer mental health and vaccine acceptance. Higher rates of depression, anxiety and /or stress were significantly associated with vaccine acceptance in one high-quality ^35^ and three medium-quality quantitative studies ^66, 83, 84^, but a further two medium-quality studies ^77, 89^ found no association. One nigh-quality study found that while increased traumatic stress was associated with vaccine acceptance, general stress or anxiety was not ^33^.

## 4. Discussion

To our knowledge, this is the first systematic review to synthesise quantitative and qualitative evidence on psychological factors influencing adolescent Covid-19 vaccine-related attitudes, intentions, and behaviours and to map the results to COM-B ^18^. While other systematic reviews investigating factors associated with adolescent Covid-19 vaccination exist ^15, 16^, they included 15 and 7 studies respectively, as compared to the 73 included in this study. Further, they did not categorise findings according to the COM-B model. Overall, COM-B was a useful framework for categorising factors related to adolescent vaccine acceptance and hesitancy, although in line with prior literature we found that mental health sat across components ^22^. Reflective motivation and social opportunity were the COM-B components most frequently associated with adolescent vaccination. Consistent with wider literature ^29, 99–101^ vaccine safety and side effect concerns and negative perceptions that the vaccine was neither effective nor necessary were critical barriers to vaccination. Therefore, interventions supporting adolescents’ assessment of the risks and benefits of vaccines should be considered.

Furthermore, this review shows that vaccination decisions have a social dimension, supporting prior vaccine ^15, 102, 103^ and health behaviour research ^104^. Our results suggest autonomy was an important component of social influence, with adolescents involved in vaccine decision-making more likely to accept vaccines. This differentiates the adolescent vaccine experience from that of younger children (with no autonomy) or adults (with full autonomy). Autonomy is also related to sociodemographic factors such as age (younger adolescents more likely to follow parental advice), deprivation (adolescents in areas of deprivation less likely to be exposed to parental recommendations of vaccinates or positive vaccination stories in their community) ^31^ and support networks: while some adolescents have stable family structures and rely on familial guidance, others e.g., those in care, may not.

Evidence as to how autonomy impacts vaccine acceptance, as well as how unsupported adolescents make effective vaccine decisions, should be another area for future research.

In this review, social media was not a major influence on adolescent vaccine behaviour, contrary to prior suggestions ^102, 105^. During the Covid-19 pandemic, social media became important for social connection given the removal of in-person social opportunities under restrictions ^106^. However, this review suggests this did not necessarily have a negative impact on vaccine acceptance. Both positive and negative vaccine messaging was shared on social media with vaccine-positive messaging more likely to come from younger users ^105^. It has been suggested that belief in conspiracy theories peaks during adolescence ^107^, but evidence that belief in conspiracy theories influenced vaccine decision-making was limited in this review. Furthermore, in one study the opposite effect was shown, with some adolescents acting as ‘debunkers’ of vaccine misinformation and conspiracy theories for their parents and families ^81^.

Many adolescent-facing vaccine interventions focus on increasing knowledge ^17, 108^. This review found mixed evidence on the role of knowledge in vaccine intention or uptake. While knowledge has been shown to be a predictor of adolescent preventive behaviours during Covid-19 ^109^, the impact of knowledge-based interventions on either adolescent health behaviours ^110^ or uptake of vaccines ^111, 112^ is limited. Therefore, while education could form one arm of vaccine interventions, adolescents should also be supported to critically assess the pros and cons of vaccines and understand how social norms impact their decisions.

Finally, this review underscores prior research ^8, 113, 114^ that vaccine acceptance or refusal are not binary concepts. Instead they span a spectrum, encompassing attitudes, questions, doubts, concerns and philosophical positions that evolve with life stages and events ^113^. In this review, for example, improving physical opportunities to receive vaccines (e.g., making vaccines more accessible, lowering their cost) and receiving information from trusted healthcare workers helped to persuade adolescents who were vaccine hesitant (defined as undecided) to accept vaccines, but did not change the minds of those with more entrenched vaccine resistant views. Therefore, a range of measures to address individual drivers to vaccine acceptance are necessary.

## 5. Limitations of included studies

Few studies included in this review were high quality. Most were cross-sectional, providing a snapshot of adolescent vaccine perspectives at one timepoint, but failing to prove causality or show how vaccine acceptance shifted over time or in response to events. Furthermore, vaccine hesitancy and acceptance were defined inconsistently across studies: some using validated, reliable vaccine hesitancy scales and others self-authoring scales or measuring vaccine intentions as binary concepts. Most studies used non-probability-based sampling, with sampling frames potentially introducing bias. For example, school-based studies excluded adolescents not attending mainstream education: that is, potentially those disengaged with society and less trusting of established authority and medicine and with lower vaccine uptake ^115^. Furthermore, some studies relied on technology-based data collection e.g., mobile phones or apps, excluding those without access to technology or digital literacy skills (again potentially affecting the least affluent or trusting communities).

Such communities are typically underserved by Covid-19 research ^116^, yet their engagement in vaccination programmes is crucial for public health. Future research methods should be inclusive of those communities. Finally, with few exceptions, participants were not involved in the design of studies. As such, factors investigated could be subject to the biases of (adult) researchers.

## 6. Limitations of this study

There is currently no consistent definition of adolescence in the literature. While we used WHO’s definition of aged 10 to 19 years, this encompasses broad developmental changes from primary education and parental decision-making to adulthood. In this context, it is hard to view adolescents as an homogenous group. Age and other sociodemographic predictors of vaccination were not included, and existing literature suggests associations between age, socioeconomic status (SES) and vaccine uptake ^117–119^. Further research should investigate to what extent age, SES and other sociodemographic variables mediate psychological factors associated with vaccine hesitancy.

## 7. Conclusion

Vaccine hesitancy is a major threat to public health. During the Covid-19 pandemic, vaccine uptake was particularly low amongst adolescents. This systematic review synthesised factors influencing adolescent Covid-19 vaccine attitudes, intention, and uptake. Mapping these factors to COM-B highlights that adolescent-facing vaccine interventions must move beyond increasing vaccine-related knowledge and should help adolescents to critically weigh vaccine pros and cons and consider external influences on their decisions.

## Data Availability

No novel data were collected as part of this study. All data are already publicly available.

## Ethics statement

All data used were in the public domain, therefore ethical approval was not required.

## Data availability

No novel data were collected as part of this study. All data are already publicly available.

## CRediT authorship contribution statement

**Angie Pitt:** Conceptualization, Data curation, Formal analysis, Methodology, Validation, Visualisation, Writing – original draft.

**G. James Rubin:** Supervision, Funding acquisition, Conceptualization, Methodology, Writing – review & editing.

**Richard Amlȏt:** Funding acquisition, Supervision, Writing – review & editing.

**Catherine Heffernan:** Supervision, Methodology, Writing – review & editing.

**Louise E. Smith:** Supervision, Funding acquisition, Conceptualization, Methodology, Validation, Formal analysis, Visualisation, Writing – review & editing.

**List of abbreviations:** World Health Organization (WHO); Preferred Reporting Items for Systematic Reviews and Meta-Analyses (PRISMA); National Heart Lung and Blood Institute (NIH); Critical Appraisal Skills Programme (CASP); Synthesis Without Meta-analysis (SWiM)

