## Supplementary files A, B, C1, C2 for "A systematic review of psychological factors influencing attitudes and intentions toward, and uptake of, Covid-19 vaccines in adolescents"

**Appendix A:** Systematic review, search strategy

**PsycINFO, Embase, MEDLINE and Scopus**

Covid-19 OR “SARS-CoV-2*” OR “coronavirus”

AND

Vaccin* OR immunis* OR immuniz* OR inocul* OR booster*

AND

Adolesc* OR teen* OR “young people” OR child* OR school* OR student* OR “young person*” OR p?ediatric OR student* OR youth* OR juvenil*

AND

Attitud* OR belief* OR knowledge* OR trust* OR Perspective* OR view* OR opinion* OR perception* OR perceiv* OR uptake OR hesit* OR accept* OR refus* OR decision* OR concern* OR barrier* OR reluct* OR anti-vaccin* OR anti-vax* OR experience* OR trait* OR messag* OR behav* OR willing* OR “side effect*” OR “adverse effect*” OR history*

Date limit: 01/01/2020 – 30/03/23

**Google search terms**

Vaccines, adolescents, teenagers, young people, covid, coronavirus, attitudes, hesitancy, uptake, intention.

**Appendix B:** Quality assessments

**Figure B1:** Quantitative studies quality assessment

| Author(s) | Were inclusion and exclusion criteria for being in the study prespecified and applied uniformly to all participants? | | Was a sample size justification or power description provided? | | Were variance and/or effect estimates provided? | | For the analyses in this paper, were the exposure(s) of interest measured prior to the outcome(s) being measure | Was the timeframe sufficient so that one could reasonably expect to see an association between exposure and outcome if it existed? | For exposures that can vary in amount or level, did the study examine different levels of the exposure as related to the outcome (e.g., categories of exposure, or exposure measured as | | Were the exposure measures (independent variables) clearly defined, valid, reliable, and implemented consistently across all study participants? (have to all be yes to pass) Some = 1, all = 2 | | Was the exposure(s) assessed more than once over time? | | Were the outcome measures (dependent variables) clearly defined, valid, reliable, and implemented consistently across all study | | Were the outcome assessors blinded to the exposure status of participants? *= N - outcome assess* | | Was loss to follow-up after baseline 20% or less? | | Were key potential confounding variables measured and adjusted statistically for their impact on the relationship between exposure(s) and outcome(s)? | | |
|---|---|---|---|---|---|---|---|---|---|---|---|---|---|---|---|---|---|---|---|---|---|---|---|
| Adjaottor et al 2022 | Y | 1 | N | 0 | Y | 1 | N | N | Y | 1 | Y | 2 | N | 0 | Y | 1 | N | 0 | NA | 0 | Y | 1 | 10 |
| Afifi et al 2021 | N | 0 | N | 0 | Y | 1 | N | N | N - some but not all | 0 | Some | 1 | N | 0 | N - not validated | 0 | N | 0 | NA | 0 | Y | 1 | 7 |
| Akhtar et al 2022 | Y | 1 | Y | 1 | NA - no analyses carried out | 0 | N | N | N | 0 | N | 0 | N | 0 | N | 0 | N | 0 | NA | 0 | NA - no analyses carried out | 0 | 4 |
| Alemu et al 2023 | N | 0 | Y | 1 | Y | 1 | N | N | CD | 0 | N | 0 | N | 0 | N | 0 | N | 0 | NA | 0 | Y - age included but not reported in results | 1 | 7 |
| Assad et al., 2023 | N | 0 | Y | 1 | Y | 1 | N | n | N | 0 | N | 0 | N | 0 | N | 0 | N | 0 | NA | 0 | Y | 1 | 6 |
| Ates et al 2023 | Y | 1 | N | 0 | Y | | N | N | Y | 1 | Y | 2 | N | 0 | Y | 1 | N | 0 | NA | | Y | 1 | 9 |
| Balaguera et al 2023 | Y | 0 | N | 0 | Y | 1 | N | N | CD | 0 | Some - survey previously validated questions not included so cannot determine whether clearly defned | 1 | N | 0 | N | 0 | N | 0 | NA | 0 | N - multinomial analysis only | 0 | 4 |
| Bergh et al 2023 | Y | 1 | N | 0 | Y | 1 | N | N | CD | 0 | CD | 0 | N | 0 | Y | 1 | N | 0 | NA | 0 | Y | 1 | 8 |
| Bhowmick et al 2022 | ND - eligibility criteria not reported | 0 | Y | 1 | Y | 1 | N | N | N | 0 | N | 0 | N | 0 | N - "validity established by experts" and "tried out" on | 0 | N | 0 | NA | 0 | N | 0 | 5 |
| Bonander et al 2022 | CD | 0 | Y | 1 | Y | 1 | N | Y | NA | 0 | Y | 1 | N | 0 | Y | 1 | N | 0 | NA | 0 | Y | 1 | 8 |
| Bracko & Simon 2022 | ND | 0 | N | 0 | Y | 1 | N | N | CD - checked survey, can't see qu's relating to psych factors | 0 | Some - additional questions for Study B not validated? | 1 | N | 0 | N - attitudes? | 0 | N | 0 | NA | 0 | NA | 0 | 4 |
| Can & Kurtulus, 2021 | Y | 1 | CD - calculation not explained | 0 | Y | 1 | N | N | CD - yes for fear scale, but not given for other questions relating to vax | 0 | Some | 1 | N | 0 | N | 0 | N | 0 | NA | 0 | N - gender included but age not included | 0 | 6 |
| Choi et al 2021 | CD - inclusion criteria not stated | 0 | N | 0 | NA | 0 | N | N | Y | 1 | N | 0 | N | 0 | N | 0 | N | 0 | NA | 0 | NA | 0 | 4 |
| Chung et al 2022 | CD - inclusion criteria mentioned but not | 0 | N | 0 | Y | 1 | N | N | Y | 1 | Some | 1 | N | 0 | N | 0 | N | 0 | NA | 0 | N | 0 | 7 |
| Coronado et al 2022 | Y | 1 | NA | 0 | Y | 1 | N | Y | NA | 0 | Y | 2 | N | 0 | Y | 1 | N | 0 | NA | 0 | Y | 1 | 9 |
| Cupertino et al 2022 | N | 0 | N | 0 | Y | 1 | N | N | N | 0 | N - youth panel designed survey | 0 | N | 0 | N - youth panel designed | 0 | N | 0 | NA | 0 | Y | 1 | 6 |

| | | | | | | | | | | | | | | | | | | | | | | | |
|---|---|---|---|---|---|---|---|---|---|---|---|---|---|---|---|---|---|---|---|---|---|---|---|
| **Das et al 2024** | CD | 0 | N - mentions calculati on but no details | 0 | N | 0 | N | N | CD - measures not reported | 0 | N | 0 | N | 0 | N | 0 | N | 0 | NA | 0 | NA | 0 | 3 |
| **Dhanker et al 2023** | Y | 1 | Y | 1 | Y | 1 | N | N | Y | 1 | N - validated by "experts" | 0 | N - only side effects data collected at diff time | 0 | CD - validated by "experts" | 0 | N | 0 | NA | 0 | Y | 1 | 7 |
| **Dvorsky et al 2022** | N | 0 | N | 0 | Y | 1 | N | N | Y | 1 | Some | 1 | N - hesitancy measured | 0 | N | 0 | N | 0 | NA | 0 | N - age not controlled for | 0 | 6 |
| **Efendi et al 2022** | Y | 1 | Y | 1 | Y | 1 | N | N | Y | 1 | Some | 1 | N | 0 | N - outcome measure not validated | 0 | N | 0 | NA | 0 | Y | 1 | 11 |
| **Efendi et al 2023** | Y | 1 | Y | 1 | Y | 1 | N | N | Y | 1 | Y | 2 | N | 0 | Y | 1 | N | 0 | NA | 0 | Y | 1 | 11 |
| **Euser et al 2022** | ND | | N | 0 | Y | 1 | N | N | CD - "several" questions had scales | 0 | N - youth panel designed survey | 0 | N | 0 | N -youth panel designed | 0 | N | 0 | NA | 0 | Y | 1 | 6 |
| **Fazel et al 2021** | Y | 1 | N | 0 | Y | 1 | N | N | Y | 1 | Some | 1 | N | 0 | Y | 1 | N | 0 | NA | 0 | Y | 1 | 11 |
| **Ganem et al 2023** | ND - prior articles on CSSNC also checked for incl/excl criteria | 0 | N | 0 | Y | 1 | N | N | Y | 1 | Some - adapted from COSMO survey, not not validated? | 2 | Y | 1 | N - not validated? | 0 | N | 0 | N - 22% | 0 | N - age not included for students | 0 | 8 |
| **Garg et al 2024** | CD | 0 | N | 0 | Y | 0 | N | N | N | 0 | CD - measures not reported | 0 | N | 0 | CD - measures not | 0 | N | 0 | NA | 0 | N - gender but not age | 0 | 3 |
| **Gewirtz-Meyden et al 2022** | CD - no inclusion/excl criteria given for child or prior parent study) | 0 | N | 0 | Y | 1 | N | N | Y | 1 | Some - not all (if the adolesecent did not accept the vaccine they were provided with a list of 11 reasons for refusal) | 1 | N | 0 | N - not validated | 0 | N | 0 | NA | 0 | Y | 1 | 8 |
| **Guneysu et al 2023** | Y | 1 | N | 0 | Y | 1 | N | N | CD | 0 | CD | 0 | N | 0 | CD | 0 | N | 0 | NA | 0 | N - age but not gender | 0 | 5 |
| **Hasanatuludhhiyah et al** | Cd | 0 | Y | 1 | Y | 1 | N | N | Y | 1 | Some - no info on how vaccine acceptance measured | 1 | N | 0 | Y | 1 | N | 0 | NA | 0 | Y | 1 | 9 |
| **Hwang et al 2023** | CD | 0 | N | 0 | Y | 1 | N | N | Y | 1 | Y | 2 | N | 0 | Y | 1 | N | 0 | NA | 0 | N | 0 | 8 |

| | | | | | | | | | | | | | | | | | | | | | | | |
|---|---|---|---|---|---|---|---|---|---|---|---|---|---|---|---|---|---|---|---|---|---|---|---|
| **Hopfer et al 2022 – QUANT ONLY (QUAL parents only)** | incl/excl criteria for adolescent ppts | 0 | N | 0 | NA – Survey and focus groups only | 0 | N | N | N | 0 | N | 0 | N | 0 | N | 0 | N | 0 | NA | 0 | N | 0 | 2 |
| **Inaba et al 2022** | N | 0 | Y | 1 | Y | 1 | N | N | Y | 1 | Some | 1 | N | 0 | N | 0 | N | 0 | NA | 0 | | 0 | 7 |
| **Janssen et al 2023** | Y | 1 | NA | 0 | N | 0 | N | N | N | 0 | N | 0 | N | 0 | N | 0 | N | 0 | NA | 0 | NA | 0 | 5 |
| **Kajiwara et al 2022** | CD – no inclusion/excl criteria given | 0 | N | 0 | NA | 0 | N | N | NA | 0 | N | 0 | N | 0 | N | 0 | N | | NA | 0 | N | 0 | 2 |
| **Klein et al 2021** | CD | 0 | N | 0 | N | 0 | N | N | CD | 0 | CD | 0 | N | 0 | CD | 0 | N | 0 | NA | 0 | CD | 0 | 2 |
| **Lee et al 2022** | N | 0 | N | 0 | Y | 1 | N | N | Y | 1 | N | 0 | N | 0 | N | 0 | N | 0 | NA | 0 | Y | 1 | 7 |
| **Li et al 2022 – 1** | N – incl/excl criteria not provided | 0 | N | 0 | N | 0 | N | N | N (checked suppl. Data) | 0 | N | 0 | N | 0 | N | 0 | N | 0 | NA | 0 | N | 0 | 4 |
| **Li et al 2022 – 2** | N – incl/excl criteria not provided | 0 | N | 0 | Y | 1 | N | N | N | 0 | N | 0 | N | 0 | N | 0 | N | 0 | NA | 0 | N – only age, not gender | 0 | 5 |
| **McKinnon et al 2023 – QUAL** | N – no exlc criteria given | 0 | N | 0 | Y | 1 | N | N | Y | 1 | Y | 2 | N | 0 | Y | 1 | N | 0 | NA | 0 | N | 0 | 8 |
| **Meraya et al 2022** | N – no incl/excl criteria given | 0 | N | 0 | N | 0 | N | N | Y | 1 | N – no info on validatation of scales | 0 | N | 0 | N | 0 | N | 0 | NA | 0 | N – multivariate regression for adult data only | 0 | 5 |
| **Middleman et al 2021** | CD – no incl/excl criteria given | 0 | N | 0 | N | 0 | N | N | Y | 1 | N – ppts agreed/disagreed with statements on likert scale, no info on | 0 | Y | 1 | N | 0 | N | 0 | Y | 1 | N – no analyses reported | 0 | 6 |
| **Middleman et al 2022"** | CD | 0 | N | 0 | N | 0 | N | N | CD | 0 | CD | 0 | Y | 1 | CD | 0 | N | 0 | CD | 0 | CD | 0 | 4 |
| **Moore et al., 2024** | Y | 1 | Y | 1 | N – not for factors | 0 | N | N | N | 0 | N | 0 | N | 0 | CD | 0 | N | 0 | NA | 0 | NA | 0 | 5 |
| **Nilsson et al 2021** | CD – no incl/excl criteria given | 0 | N | 0 | Y | 1 | N | N | Y | 1 | Yes – adapted from Brazilian survey & NRS | 2 | N | 0 | N – not validated | 1 | N | 0 | NA | 0 | N – age not included | 0 | 8 |
| **Oka et al 2022** | CD – no incl/excl criteria given | 0 | N | 0 | NA – no analyses | 0 | N | N | N | 0 | N | 0 | N | 0 | N | 0 | N | 0 | NA | 0 | N – no analyses reported | 0 | 3 |
| **Parlak & Ener, 2022** | N | 0 | N | 0 | Y | 1 | N | N | Y | 1 | N | 0 | N | 0 | Y | 1 | N | 0 | NA | 0 | N – not age | 0 | 6 |
| **Persaud et al 2023** | CD – no incl/excl criteria given | 0 | N | 0 | Y | 1 | N | N | NA – free text answers | 0 | N | 0 | N | 0 | N – not validated | 0 | N | 0 | NA | 0 | Y – but psych factors not included in analysis | 0 | 4 |
| **Pimental et al 2022** | Y | 1 | Y | 1 | Y | 0 | N | N | Y | 1 | Y | 2 | N | 0 | N – not validated | 0 | N | 0 | NA | 0 | N – age but not gender | 0 | 8 |
| **Qian et al 2023** | ND | 0 | N | 0 | Y | 1 | N | N | Y | 1 | N | 0 | N | 0 | N | 0 | N | 0 | NA | 0 | N | 0 | 5 |
| **Rehati et al 2022** | Y | 1 | Y | 1 | Y | 1 | N | N | Y | 1 | Y | 2 | N | 0 | N | 0 | N | 0 | NA | 0 | Y– stage of schooling interpreted as | 1 | 12 |

| | | | | | | | | | | | | | | | | | | | | | | | |
|---|---|---|---|---|---|---|---|---|---|---|---|---|---|---|---|---|---|---|---|---|---|---|---|
| **Roethoeft et al 2023** | Y | 1 | N | 0 | Y | 1 | N | N | Y | 1 | N | 0 | N | 0 | N | 0 | N | 0 | NA | 0 | CD | 0 | 7 |
| **Rogers et al 2021** | CD - no incl/excl criteria given | 0 | N | 0 | Y | 1 | N | N | Y | 1 | Some | 1 | N | 0 | N - not validated | 0 | N | 0 | NA | 0 | Y | 1 | 9 |
| **Rosen et al 2022*** | CD | 0 | N | 0 | Y | 1 | N | N | CD | 0 | CD | 0 | N | 0 | CD | 0 | N | 0 | CD | 0 | CD | 0 | 4 |
| **Rosen et al 2024** | Y | 1 | Y | 1 | Y | 0 | N | N | Y | 1 | Some | 1 | N | 0 | N - one question, no validated scale used | 0 | N | 0 | NA | 0 | N | 0 | 7 |
| **Roy et al 2024** | Y | 1 | Y | 1 | Y | 1 | N | N | | | N | 0 | N | 0 | N | 0 | N | 0 | NA | 0 | N | 0 | 6 |
| **Ryan et al 2023** | Y - those vaccinated were excluded | 0 | N | 0 | | | N | N | N - reasons to be/not be vaccinated selected from list | 0 | N -not validated | 0 | N | 0 | N - not validated | 0 | N | 0 | NA | 0 | Y | 1 | 6 |
| **Scherer et al 2021** | CD - incl/excl criteria not listed | 0 | N | 0 | Y | 1 | N | N | CD - no information on how "potential factors" were generated or included in | 0 | N - no info | 0 | N | 0 | N - not validated | 0 | N | 0 | NA | 0 | N | 0 | 4 |
| **Tirawi et al 2022** | CD - no incl/excl criteria given | 0 | N | 0 | N - descriptive data only | 0 | N | N | N | 0 | N | 0 | N | 0 | N | 0 | N | 0 | NA | 0 | NA - no analyses carried out | 0 | 3 |
| **Tu et al 2022** | CD - incl/excl criteria not listed | 0 | Y | 1 | | | | | Y | 1 | Some - based on PMT | 1 | N | 0 | N - not validated | 0 | N | 0 | NA | 0 | CD - gender not reported | 0 | 7 |
| **Unger et al 2023** | Y | 1 | N | 0 | Y | 1 | N | N | Y | 1 | Some - based on PMT | 1 | N | 0 | N - not validated | 1 | N | 0 | NA | 0 | Y | 1 | 10 |
| **Wang et al 2022** | CD - incl/excl criteria not listed | 0 | N - calculation not carried out | 0 | Y | 1 | N | N | Y | 1 | Some | 1 | N | 0 | N | 0 | N | 0 | NA | 0 | Y | 1 | 8 |
| **Wang et al 2022** | | | | | | | | | | | | | | | | | | | | | | | |
| **Wirunpan et al 2021** | CD - incl/excl criteria not listed | 0 | N | 0 | Y | 1 | N | N | Y | 1 | N | 0 | N | 0 | N | 0 | N | 0 | NA | 0 | CD - not reported | 0 | 5 |

| | | | | | | | | | | | | | | | | | | | | | | | |
|---|---|---|---|---|---|---|---|---|---|---|---|---|---|---|---|---|---|---|---|---|---|---|---|
| **Wong et al 2022** | CD - incl/excl criteria not listed | 0 | N | 0 | Y | 1 | N | N | N | 0 | N | 0 | N | 0 | N | 0 | N | 0 | NA | 0 | Y | 1 | 6 |
| **Wong et al 2022** | CD | 0 | N | 0 | N | 0 | N | N | N | 0 | N | 0 | N | 0 | N | 0 | N | 0 | NA | 0 | N | 0 | 4 |
| **Zhang et al 2022** | CD - incl/excl criteria not listed | 0 | N | 0 | Y | 1 | N | N | Y | 1 | Y | 2 | N | 0 | N | 0 | N | 0 | NA | 0 | N | 0 | 7 |
| **Zilhadia et al 2022** | CD - incl/excl criteria not listed | 0 | Y | 1 | Y | 1 | N | N | Y | 1 | Y | 2 | N | 0 | N | 0 | N | 0 | NA | 0 | N | 0 | 8 |

**Figure B1:** The NIH Quality Assessment Tool for observational cohort and cross-sectional studies. Adaptations were: questions answered ‘yes’ were scored one point, and questions answered ‘no’ were scored zero; the question ‘Were the exposure measures (independent variables) clearly defined, valid, reliable, and implemented consistently across all study participants?’ was scored from zero to two, because some studies did not use validated exposure measures (zero points), some used validated measures for some exposure measures (one point) and some used all validated measures (two points); questions six and seven were not included. Reports scoring between zero and five were deemed low quality, those scoring six to nine were deemed medium quality and those scoring 10 or above were considered high quality.

**Figure B2:** Quality assessment: before-after studies

| Author(s) | Outcome(s) | Was the study question or objective in this paper clearly stated? | | Were eligibility/selection criteria for the study population prespecified and clearly described? | | . Were the participants in the study representative of those who would be eligible for the test/service/intervention in the general or clinical population of interest? | | Were all eligible participants that met the prespecified entry criteria enrolled? | | Was the sample size sufficiently large to provide confidence in the findings? | | Was a sample size justification or power description provided? | | Was the test/service/intervention clearly described and delivered consistently across the study population? | | Were the outcome measures prespecified, clearly defined, valid, reliable, and assessed consistently across all study participants? | | Were the people assessing the outcomes blinded to the participants' exposures/interventions? | | Was the loss to follow-up after baseline 20% or less? Were those lost to follow-up accounted for in the analysis? | | Did the statistical methods examine changes in outcome measures from before to after the intervention? Were statistical tests done that | | Were outcome measures of interest taken multiple times before the intervention and multiple times after the intervention (i.e., did they use an interrupted time-series design)? | | . If the intervention was conducted at a group level (e.g., a whole hospital, a community, etc.) did the statistical analysis take into account the use of individual-level data to determine effects at the group level? | | |
|---|---|---|---|---|---|---|---|---|---|---|---|---|---|---|---|---|---|---|---|---|---|---|---|---|---|---|---|---|
| Ganczak et al, 2021 | Intention | Y | 1 | Y | 1 | Y | 1 | Y | 1 | Y | 1 | Y | 1 | Y | 1 | N- not validated scales | 0 | N - ppts were assessors | 0 | NR | 0 | Y | 1 | N | 0 | NA | 0 | 8 |

**Figure B2:** The NIH Before-After Studies with No Control Group for interventions tool. No amendments were made to this tool. Questions answered 'yes' were scored one point, and questions answered 'no' were scored zero. Studies scoring zero to five were considered low quality, scoring six to nine were medium quality and scoring ten or over were high quality.

### Figure B3: Qualitative study quality assessments

| Author(s) | Type of qual data | Was there a clear statement of the aims of the research? | Score | Is a qualitative methodology appropriate? | Score | Was the research design appropriate to address the aims of the research? | Score | Was the recruitment strategy appropriate to the aims of the research? | Score | Was the data collected in a way that addressed the research issue? | Score | Has the relationship between researcher and participants been adequately considered? (reflexivity) | Score | Have the study population been involved in developing discussion guide? | Score | Have ethical issues been taken into consideration? | Score | Was the data analysis sufficiently rigorous? | Score | Is there a clear statement of findings? | Score | How valuable is the research? (0-2) | Total | Evaluation |
|---|---|---|---|---|---|---|---|---|---|---|---|---|---|---|---|---|---|---|---|---|---|---|---|---|
| **Alemu et al., 2023** | **Qual interviews** | Y | **1** | Y | **1** | Y | **1** | Y | **1** | Y | **1** | CT - not reported | **0** | N | **0** | N | **0** | N | **0** | N | **0** | 1 | **5** | Low |
| Budhwani et al., 2021 | Qual interviews | Y | 1 | Y | 1 | Y | 1 | CT - not reported | 0 | Y | 1 | CT - not reported | 0 | | | CT - not reported | 0 | CT - not reported | 0 | Y | 1 | 1 | 6 | Low |
| Delgado et al., 2022 | Qual interviews | Y | 1 | Y | 1 | Y | 1 | Y | 1 | Y | 1 | CT - not reported | 0 | | | Y - privacy | 1 | Y | 1 | Y | 1 | 1 | 9 | Medium |
| Fisher et al., 2021 | Qual interviews | Y | 1 | Y | 1 | Y | 1 | Y | 1 | Y | 1 | CT - not reported | 0 | | | CT - not reported | 0 | Y | 1 | Y | 1 | 2 | 9 | Medium |
| Garcia et al., 2021 | Qual interviews | Y | 1 | Y | 1 | Y | 1 | Y | 1 | Y | 1 | CT - not reported | 0 | | | CT - not reported | 0 | Y | 1 | Y | 1 | 2 | 9 | Medium |
| Groenewald et al 2023 | Qual interviews | Y | 1 | Y | 1 | Y | 1 | Y | 1 | Y | 1 | Y | 1 | N | 0 | Y | 1 | Y | 1 | Y | 1 | 2 | 11 | High |
| Kenworthy et al., 2022 | Focus groups | Y | 1 | Y | 1 | Y | 1 | Y | 1 | N - adolescents only interviewed with caregiver | 0 | N | 0 | Y | 1 | Y | 1 | N - qual statements analysed quantatively | 0 | Y | 1 | 1 | 8 | Medium |
| McKinnon et al., 2023 | Mixed methods/interviews | Y | 1 | Y | 1 | Y | 1 | Y | 1 | Y | 1 | Y - youth trained to be interviewers | 1 | Y | 1 | Y | 1 | Y | 1 | Y | 1 | 2 | 12 | High |
| Mansfield et al., 2023 | Interviews | Y | 1 | Y | 1 | Y | 1 | Y | 1 | Y | 1 | Y | 1 | N | 0 | CT - not reported | 0 | Y | 1 | Y | 1 | 1 | 9 | Medium |
| Nilsson et al., 2021 | Mixed methods / Survey and text from open question | Y | 1 | Y | 1 | N | 0 | Y | 1 | N - only one Q "Feel free to comment about your thoughts on vaccination" | 0 | N | 0 | N | 0 | CT - not reported | 0 | Y | 1 | Y | 1 | 1 | 6 | Low |
| Persaud et al., 2023 | Mixed methods / free text survey (q not reported) | Y | 1 | Y | 1 | N | 0 | CT - not reported | 0 | N | 0 | N | 0 | N | 0 | CT - not reported | 0 | N | 0 | Y | 1 | 1 | 4 | Low |
| Ramaiya et al 2023 | Qualitative focus groups | Y | 1 | Y | 1 | Y | 1 | N - inconsistent across groups | 0 | Y | 1 | N | 0 | N | 0 | Y | 1 | Y | 1 | Y | 1 | 2 | 9 | Medium |
| Schmidt-Sane et al., 2022 | Interviews & focus groups | Y | 1 | Y | 1 | Y | 1 | CT - not reported | 0 | Y | 1 | N | 0 | Y | 1 | CT - not reported | 0 | Y | 1 | Y | 1 | 2 | 9 | Medium |
| Uroko & Nche, 2023 | Interviews | Y | 1 | Y | 1 | Y | 1 | Y | 1 | Y | 1 | N | 0 | N | 0 | CT - not reported | 0 | Y | 1 | Y | 1 | 1 | 8 | Medium |

**Figure B3:** The Critical Appraisal Skills Programme (CASP) tool. An additional question ‘Have the study population been involved in developing discussion guide?’ was included, and answers for questions were scored yes (one point), no (zero points). The question ‘How valuable is the research?’ was scored from zero to two. Studies scoring zero to six were considered low quality, scoring seven to 11 were medium quality and scoring 12 or over were high quality.

### Appendix C

**Table C1:** Overview of full quantitative results on psychological factors influencing vaccination attitudes, intention and uptake.

| Citation *Conference abstract | Attitudes (Descriptive data) | Reasons to be vaccine acceptant (descriptive data) *based on graph estimates | Reasons to be vaccine hesitant (descriptive data) *based on graph estimates | Associations | Not associated | Outcome |
|---|---|---|---|---|---|---|
| **Adjaottor et al., 2022** | | | | Correlations:<br><br>Pearsons r:<br>Significant positive relationships between COVID-19 vaccination acceptance and:<br>Believing Covid-19 information, r = 0.145, p = <0.01<br>Significant negative relationship between COVID-19 vaccination acceptance and traumatic stress (r = −0.090, p = 0.01).<br><br>Hierarchical linear regression:<br>Associated with vaccine acceptance:<br>believing COVID-19 information (β = 0.125, p = 0.001)<br>Danger and contamination fears β = 0.140, p = 0.025),<br>Traumatic stress, β = −0.176, p < 0.001) are its predictive factors, which account for 6.1% of all possible predictive factors (F(17, 762) = 2.910, p < 0.001) | Correlations:<br><br>Pearsons r:<br>No significant relationship between vaccine acceptance and:<br>Fear of COVID-19, r = 0.027, p = nr.<br>Perceived stigma from COVID-19, r = -0.008, p = nr.<br>Self-stigma from COVID-19, r = 0.033, p = nr.<br>Preventive COVID-19 infection behaviours, r = 0.032, p = nr.<br>COVID-19 stress, r = 0.029, p = nr.<br>Socio-economic consequences (subscale of COVID-19 stress)<br>Xenophobia, r = 0.047, p = nr.<br>Danger & Contamination, r = 0.060, p = nr.<br>Compulsive checking, r = 0.036, p = nr.<br><br>Not associated with vaccine acceptance:<br>Fear of Covid-19, β = -0.12, p = 0.777<br>Perceived stigma from Covid-19, β = -0.055, p = 0.165<br>Self-stigma from Covid-19, β = 0.027, p = 0.517 | I / B |

| | | | | |
|---|---|---|---|---|
| | | | Preventive Covid-19 infection behaviours, β = 0.011, p = 0.775 Xenophobia, β = -0.002, p =0.978 Socio-economic consequences, β = -0.043, p = 0.387 Compulsive checking, β = 0.063, p = 0.178 | |
| **Afifi et al., 2021** | Worried that vaccine is not safe (65.9%)<br><br>Don't know enough about vaccine (60.9%)<br><br>Do not think it will work (23.9%)<br>It might cost me money to get it (13.8%)<br><br>Not concerned about getting Covid-19 (15.9%)<br>I do not get any vaccines (5.1%)<br>I don't know how to get it (4.4%)<br><br>Other (13%)<br><br>No response (6.5%)<br>OpenText answers:<br>concerns for vaccine safety because participants perceived that COVID-19 vaccine development was rushed and protocols were skipped, media reports of people getting sick from the vaccine, not enough research on the vaccine, side effects | *n/a - only reported by overall population 16-21* | *n/a -only reported by overall population 16-21* | I |

| | | | | | |
|---|---|---|---|---|---|
| | | cannot be known in the short term, and fear of needles | | | |
| **Akhtar et al., 2023** | Persuaded by parents (36.1%)<br>Friends getting vaccinated (27.7%)<br>Doctors' advice (16.6%) | Fear of side effects of vaccination (65.2%)<br>Allergies (22.2%)<br>No benefit from vaccination (16.6%)<br>No information about Covid-19 vaccination (9.7%)<br>Any chronic medical condition (8.3%)<br>Injection shall hurt (2.7%) | | | I |
| **Alemu et al., 2023** | | | Multivariable Logistic Regression.<br>Associations with vaccine hesitancy:<br>Social media as source of info about Covid-19 vaccination, AOR = 2.42 (1.06-5.57) CI 95%, p = 0.037),<br>HCW as source of info about Covid vax, AOR = (0.05 - 0.038) 95% CI, p = 0.038)<br>Poor knowledge about COVID-19 disease (AOR = 3.18, 95% CI, p = 0.001)<br>Poor knowledge about COVID-19 vaccine (AOR = 5.66, 95% CI, 2.91–11.0, p = 0.00)<br>Unfavourable attitude towards COVID-19 vaccine (AOR = 5.2, 95% CI, 2.76–9.79, p = 0.00) | Multivariable Logistic Regression.<br>Not associated with vaccine hesitancy:<br>Friends/family as source of information: (AOR = 4.57(0.27–1.2, p = 0.14) | I / B |

| | | | | | | |
|---|---|---|---|---|---|---|
| **Assad et al., 2023** | | | Insufficient information (31%)<br><br>Misinformation & rumours (26%)<br><br>Vaccine not safe (18%)<br><br>Distrust of manufacturers (17%)<br><br>Vaccine not effective (8%) | Multivariate analysis:<br><br>Positively associated with vaccine acceptance:<br><br>Parents Covid-19 vaccination OR = 2.66, 95%CI, 1.40-5.05)<br><br>Attitude (will you recommend vaccine to others?) OR = 8.62, 95%CI, 4.60-16.17<br><br>Negatively associated with vaccine acceptance:<br><br>Fear of Covid-19 OR = 0.14, 95%CI, 0.06-0.29 | Multivariate analysis:<br><br>Belief that vaccine will prevent infection or reduce severity, OR = nr (significant in univariate analysis only)<br><br>Belief that the vaccine is important to eliminate Covid-19 in Iraq, OR = nr (significant in univariate analysis only) | |
| **Ates et al., 2023** | | | Fear of side effects (36.3%)<br>Uncertainty about effect of vaccine/insufficient studies (21.7%)<br><br>No valid reason to not get vaccinated (40.1%) | Logistic regression:<br>Predictors of vaccination:<br>Parental vaccine hesitancy, OR = 0.91, 95%CI, 0.87-0.95<br>Anxiety disorder (p = <0.001) | Logistic regression:<br>Not predictors of vaccination:<br>Externalizing score, p = 0.994<br>Internalizing score, p = 0.410<br>Prosocial behaviour, p = 0.278<br>Fear of coronavirus before and after the discovery of the vaccine (results nr ) | I |
| **Balaguera et al., 2023** | Perceived risk from Covid-19:<br>Low (70.18%)<br><br>Moderate (28.7%) | | | Beliefs associated with vaccine refusal:<br>Covid doesn't exist, (OR 7.09, 95% CI 1.62–31.02, p=0.009)<br>Covid-19 created to make money (OR = 6.79 95%CI 2.35–19.59, | Beliefs not associated with vaccine refusal:<br>Created in a lab (OR = 1.95, 95%CI 0.74-5.12, p = 0.25)<br>Created to reduce population size (OR = 2.14, 95% CI0.78-5.84, p = | I / B |

| | | | | | | |
|---|---|---|---|---|---|---|
| | High (1.75%)<br><br>Covid-19 produced in laboratory to cause harm (50.69%)<br>Created to reduce size of the world population (3.79%)<br>Created to make money/for politics (35.8%)<br>Created to sterilize population (7.24%)<br>Vaccine has some component that undermines welfare of individual (12.76%)<br>Could cause alterations in coagulation, heart disease, infertility, neurological disease (21.72%) | | | p=0.0005)<br>COVID-19 vaccine causes sterility (OR 5.38, 95%CI1.52–18.93, p=0.0016)<br>Vaccine can cause death in the medium or long term (OR 14.92, 95%CI5.36–41.53, p=0.00001.<br><br>Beliefs associated with vaccine indecision:<br>Has technologies designed to damage health (OR 3.33 95% CI 1.02-10.79, p = 0.0007)<br>Vaccine can lead to death (OR 3.89, 95% CI 1.207-12.56, p = 0.00002) | 0.96)<br>Causes health damage (OR = 2.65, 95%CI 0.98-7.14, p = 0.46)<br><br>Beliefs not associated with vaccine indecision:<br>Created in a lab (OR = 0.75, 95%CI 0.23-2.43, p = 0.75)<br>Created to reduce population size (OR = 0.95, 95% CI 0.28-3.16, p = 0.907)<br>Created due to money and politics (OR = 2 95%CI 0.59-6.7, p = 0.12)<br>Designed for sterilisation (OR = 1.33, 95%CI 0.32-5.45, p = 0.13)<br>Causes health damage (OR = 0.91, 95%CI 0.29-2.88, p = 0.46) | |
| **Bergh et al., 2023** | | Worry I might get Covid-19 (36.0 %)<br><br>Covid-19 would be a serious illness for people in my community (68.5%)<br><br>Vaccine will protect me from becoming very sick (56.7%)<br><br>Vaccine is safe (50.9%)<br><br>Most of my friends think getting the vaccine is a good thing (51.5%) | Fear of vaccine needle (58.0%)<br><br>Vaccine site too far away (26.4%)<br><br>Do not know where to get the vaccine (14.6%)<br><br>Vaccine centre opening times not convenient (10.3%)<br><br>Vaccine centre waiting times too long (22.3%) | Multivariate analysis:<br><br>Positively associated:<br><br>The vaccine will protect me from getting very sick (OR = 1.11, 95%CI, 1.04-1.19)<br><br>The vaccine will be safe for me (OR = 1.31, 95%CI, 1.18-1.44)<br><br>Negatively associated: | Someone in family has died from Covid-19<br><br>(OR = 1.08, 95%CI, 0.93-1.24)<br><br>Worry I might get Covid-19 (OR = 1.00, 95%CI, 0.96-1.04)<br><br>It would be a serious illness for people in my community (OR = 1.01, 95%CI, 0.95-1.07)<br><br>Most of my friends think getting the Covid-19 vaccine is a good thing (OR = 1.00, 95%CI, 0.97-1.03) | I / B |

| | | | | | | |
|---|---|---|---|---|---|---|
| | | Most of my family think getting the vaccine is a good thing (58.3%) | It will be expensive to get to the vaccine site (4.9%)<br><br>Cannot go to the vaccine site on my own (18.3%)<br><br>There are not enough vaccines at the site I want to go to (6.7%) | Fear of vaccine needle (OR = 0.85, 95%CI, 0.82-0.88) | Most of my family think getting the Covid-19 vaccine is a good thing (OR = 1.02, 95%CI, 0.98-1.07)<br><br>Do not know where to go for the vaccine (OR = 0.83, 95%CI, 0.68-1.01)<br><br>It will be expensive to get to the vaccine site (OR = 1.16, 95%CI, 0.99-1.36)<br><br>Cannot go to the vaccine site alone (OR = 0.84, 95%CI, 0.69-1.02) | |
| **Bhowmick et al., 2022** | I will encourage friends/family to take vaccine (88%)<br>I should take vaccine as soon as possible (83%)<br>vaccination of school adolescents is necessary to open the school again (81.9%)<br>Covid vaccine is essential only for the elderly and adults (26.5%)<br>after vaccination I can lead a normal life (33.8%)<br>I will not get Covid infection after vaccine (74.6%)<br>Covid vaccine is safe to adolescents (78.8%) | Recommended by government (22.7%)<br>Seeing many people get infected with Covid (21.5%)<br>Scared about getting Covid-19 (19%)<br>Did not want to neglect studies (14.4%)<br>Recommended by family members (12.9%)<br>Recommended by school authority (9.4%)<br>Vaccine accessibility/ Vaccine being free | | Associated with good vaccine attitude:<br><br>T-Tests:<br><br>Vaccine accessibility in nearby places, p < 0.00<br><br>Vaccine provided free of cost, p = 0.06 | | A / B |

| | | | | | | |
|---|---|---|---|---|---|---|
| **Bonander et al., 2022** | | | | Associated with vaccine uptake:<br><br>Multivariate analysis:<br><br>Receiving pre-booked appointment<br><br>Municipal analysis:(OR = 12.9, 95%CI, 9.0-16.8)<br><br>Difference in difference estimation (OR = 15.6, 95%CI, 11.6-20.0) | | B |
| **Bracko & Simon, 2022** | | Self-protection and protection of others (12%)<br>Contribution to herd immunity (7.6%)<br>General trust in vaccine(s) (2.4%) | Vaccine safety uncertainties (50.2%)<br>No interest in getting vaccinated (2.8%)<br>No need to do so because of young age, strong immune system or having already had the disease (2.4%)<br>People from high-risk groups and those relevant for a functioning system should become vaccinated first (1.3%)<br>Am a vaccination opponent (1%)<br>Want to wait (4.2%)<br>Undecided (4.3%) | Associated with willingness to be vaccinated:<br>Knowledge about viruses, d = 0.832, 95% CI 0.665-1.0, p = <.001 | | I |
| **Can & Kurtulus, 2021** | Belief in vaccine efficacy (55.2%) | Fear of transmitting virus to family and loved ones (46.2%)<br>Threat of pandemic (8.6%) | Side effects (30%)<br>Not believing effectiveness of vaccine (5.9%) | T-Tests:<br>Associated with belief in vaccine efficacy:<br>Higher fear = perceived lower efficacy (t = 1.348, p = 0.027)<br>Associated with not wanting to be | n/a | A / I |

| | | | | | | |
|---|---|---|---|---|---|---|
| | | Fear of disease and death (7.9%) | Feeling like a guinea pig (3.4%) | vaccinated:<br>Higher fear score (t = 0.471, p = 0.036) | | |
| **Choi et al., 2021** | How likely would you think the Covid-19 vaccines are preventive:<br>Extremely likely (10.3%) Somewhat likely (40.2%)<br>How likely would you think the Covid-19 vaccines are safe?<br>Extremely likely (6%)<br>Somewhat likely (18.8%)<br>Covid-19 is a serious disease?<br><br>Extremely likely (59.8%) Somewhat likely (39.5%)<br>There are members in my family who can become severe if they get Covid-19:<br><br>Extremely likely (21.4%) Somewhat likely (29.9%)<br>My family or I could get Covid-19:<br><br>Extremely likely (32.5%) Somewhat likely (60.7%)<br>I'm worried that I or | | | n/a - multivariate analysis on parental data only | n/a(45) | A |

| | | | | |
|---|---|---|---|---|
| | someone in my family could get Covid-19:<br><br>Extremely likely (41%)<br>Somewhat likely (44.4%)<br><br>I could easily get Covid-19:<br><br>Extremely likely (9.4%)<br>Somewhat likely (42.7%)<br>Actively searched for Covid-19 information (17.9%)<br>Anxiety of vulnerability to COVID-19 in children (52.1%) | | | |
| **Chung et al., 2022** | | Multivariable binary logistic regression:<br>Associated with greater willingness to be vaccinated:<br>Being trustful (aOR = 12.40, 95%CI = 7.72–19.93, p < 0.001)<br>Being neutral (aOR = 3.37, 95%CI 2.55–4.45, p < 0.001)<br><br>Multivariable binary logistic regression:<br>Assoc with vaccine uptake:<br>Trust in govt on pandemic management (aOR = 1.63 95%CI 1.06–2.52, p = 0.026)<br>Willing (OR = 11.42 95%CI 7.47–17.44, p < 0.001) and neutral [(OR = 3.25, 95%CI 2.38–4.45, p < 0.001) to receive the vaccine | Not associated with intending to vaccinate:<br>Loneliness (aOR 0.90 95%CI 0.50-1.62, p = nr)<br>Being "slightly worried" about pandemic (aOR = 0.96, 95%CI 0.44-2.09, p = nr)<br>Being "extremely worried" about pandemic (aOR = 1.79, 95%CI 0.54-5.92, p = nr)<br>Mild v severe mental health problem (aOR = 0.68, 95%CI 0.36-1.27, p = nr)<br>Normal mental health v severe mental health problem (aOR = 0.69, 95%CI 0.35-1.37, p = nr)<br>Low v high resilience (aOR = 2.59, 95%CI 0.67-10.05, p = nr) | I / B |

| | | | | | |
|---|---|---|---|---|---|
| | | | Multivariable binary logistic regression:<br>predictors of intention to receive vaccine (among unvaccinated)<br>being trustful (aOR = 4.49, 95%CI 2.06–9.75, p < 0.001) and neutral (aOR = 1.97 95%CI 1.23–3.16, p = 0.005) to the government regarding pandemic management BUT not after adjustment for willingness to be vaccinated<br>Life satisfaction (aOR = 1.12, 95%CI 1.00–1.24, p = 0.045.<br>Being “very worried” about COVID-19 pandemic (aOR = 3.26, 95%CI 1.16–9.15, p = 0.024) | | |
| **Coronado et al., 2023** | | | Associated with uptake:<br><br>Initiation of HPV vaccine (OR = 4.02, 95%CI, 3.81-4.24) | | B |
| **Cupertino et al., 2022** | To protect the general population (46.9%)<br>To return to a normal life (31.0%) | Fear of side effects (40.4%)<br>Parent decision to not be immunized (23.7%)<br>Vaccine not efficacious (14.9%)<br>Influenced by parents and family’s decisions (65.8%) | n/a - sociodemographic variables only | n/a - sociodemographic variables only | B |

| | | | |
|---|---|---|---|
| **Das et al.,<br>2024** | Vaccine will protect me (65.45%)<br><br>Vaccine is generally safe (44.54%)<br><br>Vaccines will help control the pandemic (55.44%)<br><br>Motivated to get vaccine by:<br><br>Parents (15.58%)<br><br>Husband (3.9%)<br><br>Other family members (2.6%)<br><br>Teachers (72.73%)<br><br>Self (5.19%) | Vaccine will not protect me (3.64%)<br><br>Vaccines are unsafe (12.73%)<br>– Due to side effects (100%)<br><br>Unvaccinated/unwilling to be vaccinated in future:<br><br>Fear of side effects (50%)<br><br>Considered vaccine a hoax (50%) | I / B |
| **Dhanker et<br>al., 2023** | Fear of getting COVID-19 infection (50.5%)<br>family recommendation (34.8%)<br>Government recommendation (33.2%)<br>School authority's recommendation (23%)<br>Many people getting Covid infection (22.7%)<br>For uninterrupted future studies (21.4%)<br>To avail benefits of vaccination (11.1%)<br>Perception of Covid-19 vaccine (agree / strongly agree):<br>Will not have COVID-19 infection after vaccination (27.2%)<br>My vaccination will be | Vaccine might produce serious side effects (35.7%)<br>COVID-19 vaccine is not safe (2.8%) | I / B |

| | | | | | | |
|---|---|---|---|---|---|---|
| | | beneficial to others (80.2%)<br>The vaccine may produce side effects (35.7%)<br>No need to follow COVID appropriate behaviour after vaccination (18.6%)<br><br>Vaccines are safe (30.9%)<br><br>I have a good perception of the vaccine (70.9)<br>After vaccination one can lead a normal life (53.4%)<br>The vaccine will reduce the severity of COVID -19 infection (73.2%)<br>The vaccine is essential for reopening schools (88.9%)<br>The vaccine is available in all government health facilities (79.5%) | | | | |
| **Dvorsky et al 2022** | Confidence in the safety of vaccines:<br>Higher in adolescents without ADHD (M = 1.65) compared to adolescents with ADHD (M = 0.95; F(1) = 7.73, p = .006, d = 0.40 | | | Associated with vaccine willingness:<br>Without ADHD more likely to report intent to vaccine than those with ADHD χ2(3) = 10.03, p = .018, d = 0.36<br>Confidence in the safety of vaccines higher in adolescents without ADHD (M = 1.65) compared to adolescents with ADHD (M = 0.95; F(1) = 7.73, p = .006, d = 0.40<br>Experiencing a negative impact of COVID-19 on relationships (β = 0.19, p = .004), greater concerns about COVID-19 (β = 0.17, p = .017), and greater media use (β = 0.16, p = .020) associated with higher levels of vaccination | Adolescents with ADHD:<br><br>Concerns about pandemic not associated with increased vaccine safety confidence (β = 0.12, p = .255) | I |

| | | | | |
|---|---|---|---|---|
| | | willingness for all adolescents.<br>Significant interaction between ADHD status and not following social distancing guidelines in predicting vaccination willingness (β = 0.44, p = .022).<br><br>With ADHD only:<br>Engaging in large gatherings associated with less vaccine willingness (β = 0.37, p < .001), and not adolescents without ADHD (β = 0.07, p = .472)<br><br>Without ADHD only:<br><br>Increased concerns about pandemic associated with increased vaccine safety confidence (β = 0.32, p < .001) | | |
| **Efendi et al., 2022** | Non-adherence to vaccines 1/2:<br>Parents forbade them (49.6%) / (57.5%)<br>Fear of side effects (22.9%) / (19.5%)<br>Efficacy doubts (15.1%) / (10.8%)<br>Did not believe in existence of Covid-19 (0.8%) / (10.4%)<br>Worried about long-term effects of vaccine (8.4%) / (0.1%)<br>Religious/traditional leaders' doctrine (0.4%) / (0.6%)<br>Others (2.8%) / (1.1%) | Bivariate analysis:<br>Associated with vaccine adherence:<br>Vaccine willingness (x2 = 2261.324, p <.001)<br>Attitude toward Covid-19 vaccine (x2 = 3562.956, p = <.001)<br>Knowledge level of Covid-19 vaccine (x2 = 2965.669, p <.001)<br>Confidence in Covid-19 vaccine (x2 = 3562.956, p = <.001)<br><br>Binary logistic regression: adolescents who were not willing to get vaccinated had an adherence rate [AOR = 0.159; 95% CI = 0.130-0.195] 0.159 times lower than vaccinated adolescents.<br>High level of knowledge, AOR = | *n/a - sociodemographic only* | I / B |

| | | | |
|---|---|---|---|
| | | 1.962 (1.568-2.455), 95% CI<br>Positive attitude to Covid-19 vaccine, AOR = 7.072 (5.652-8.849), 95%CI<br>High level of confidence in Covid-19 vaccine, AOR = 1.872 (1.495-2.343) 95% CI | |
| **Efendi et al., 2023** | | Bivariate analysis:<br><br>Positively associated with vaccine adherence:<br><br>National / Java / Maluku & Papua<br><br>High knowledge of vaccine (x2 = 1798.53 / 20.47 / 1711.94)<br><br>Belief in vaccine (x2 = 2874.01 / 91.79 / 226.71)<br><br>Attitude toward vaccine (x2 = 3336.92 / 1711.94 / 1554.95 / 1911.91)<br><br>Mediation analysis (psych factors only):<br><br>Attitude to vaccine significant positive mediator in association between belief in vaccine and adherence to vaccine ($p < 0.01$) | B |

| | | | | | | |
|---|---|---|---|---|---|---|
| **Euser et al., 2022** | Vaccine beliefs (%age completely agree*): Willing/hesitant/unwilling: The vaccine protects youth (88%)/(62%)/(28%) The vaccine protects others (90%)/(70%)/(32%) Afraid of side effects (15%)/(58%)/(65%) Vaccine is safe (88%)/(55%)/(18%) Vaccinating youth helps end the crisis (82%)/(55%)/(18%) Important to follow advice from the government (80%)/(55%)/(20%) My friends would get the vaccine: 88% / 68% / 38% My parents expect me t get vaccinated - 62% / 42% / 16% My friends expect me to get vaccinated - 62% / 48% / 22% | Willing/Non-willing: I want to protect myself (81%)/(41%) I want to protect others (79%/40%) I want to help ending the corona crisis (67%/42%) I want to go on vacation abroad without testing for corona (52%/44%)<br><br>I want to go to an event without testing for corona (42%/33%) I no longer want to keep distance from others (40% /25%) My parents want me to get vaccinated (23%/11%) I want to do what is best according to the government (13%/7%)<br><br>Other (2%)/ (8%) | Not willing / hesitant: I am afraid of unknown long-term effects of the vaccine 73% / 56% I do not know enough about the vaccine yet 38% /45% I am afraid of side effects 38% / 41% I do not think I will get sick of the coronavirus 38% / 24% I think the vaccines are not safe 33% / 10% All vulnerable people have already been vaccinated 30%/ 15% My parents do not want me to get vaccinated 17% / 3% I think the vaccines are not effective 15% / 6% I am afraid of needles 14% / 17% I do not think I will get infected with the coronavirus 13% / 8% Other 10% / 7%<br><br>Vaccine beliefs: Willing/hesitant/unwilling | Binary logistic regression: Willing v not willing Assoc with willingness to be vaccinated: Belief that vaccines protects young people, (OR = 1.45, 95%CI 1.08-1.94, p = <.05 Belief vaccine is safe (OR = 1.80, 95%CI 1.36-2.38, p <.001) Belief vaccinating youth helps end crisis (OR = 1.74, 95%CI 1.34-2.26, p < .001) Less afraid of side effects (OR = 0.54, 95%CI 0.44--0.67p = <.001 Social: My friends would get vaccine (OR = 1.33, 95%CI 1.05-1.68, p = <.05) My parents expect me to vaccinate (OR = 2.05, 95%CI 1.65-2.56, p <.001)<br><br>Willing v hesitant: The vaccine protects others, OR = 1.45 (1.02-2.06) 95% CI Vaccine is safe (OR = 1.45, 95%CI 1.06-1.98, p <.001) Less afraid of vaccine side effects (OR = 0.43, 95%CI 0.35-0.53, p <.001) Important to follow advice from govt (OR = 1.49, 95%CI 1.06-2.09, p = <.05) Social: My parents expect me to vaccinate (OR = 1.46, 95%CI 1.16-1.84, p = <.01) | Binary logistic regression: Willing v not willing: Belief the vaccine protects others (OR = 1.30, 95%CI 0.97-1.75, p = nr) Important to follow advice from the government (OR = 1.32, 95%CI 0.99-1.76, p = nr) My friends expect me to vaccinate (OR = 1.04, 95%CI 0.83-1.32, p = nr)<br><br>Willing v hesitant Belief the vaccine protects youth (OR = 0.74, 95%CI 0.51-1.09, p = nr) Vaccinating youth helps end the crisis (OR = 0.95, 95%CI 0.70-1.30, p = nr) My friends would get the vaccine (OR = 0.82 , 95%CI 0.62-1.07, p = nr) My friends expect me to vaccinate (OR = 1.11, 95%CI 0.88-1.40, p = nr) | I |
| **Fazel et al., 2021** | | | | Multinomial regression. Vaccine hesitant/undecided: Spent more than four hours a day | Not associated with vaccine hesitant/undecided: Paranoid feelings (BCAP) (OR=1.02, | I |

| | | | | |
|---|---|---|---|---|
| | | on social media, OR = 1.49 (1.36-1.63), 95% CI, p <.001<br>Identified less with being part of school community, (OR=1.10, 95%CI: 1.07-1.14, p < 0.001)<br>Depression/anxiety (RCADS score) (OR= 0.85, 95%CI: 0.80-0.90, p < 0.001)<br>Mental health - as RCADS total score increased, the odds of students describing themselves as undecided (or opting-out of) a vaccination decreased in comparison with describing oneself as opting-in.<br><br>Opting out of vaccination:<br>Identified less with being part of school community (OR=1.16, 95%CI: 1.11-1.20, p < 0.001)<br>Spent more than four hours a day on social media (OR = 1.51, 95%CI 1.33-1.72, p <.001)<br>Been bullied in the past year (OR = 1.28, 95%CI 1.03-1.6, p = 0.027)<br>Depression/anxiety (RCADS score) (OR=0.81, 95%CI: 0.74-0.88, p < 0.001) | 95%CI: 0.96-1.08, p = 0.558)<br>Frequency of bullying (OR = 1.07, CI95% 0.91-1.26, p = .387)<br>Feeling lonely (OR = 1.08, 95%CI: 0.96-1.21, p = 0.223<br><br>Not associated with opting-out of vaccine:<br>Paranoid feelings (BCAP) (OR = 1.08, 95%CI: 1.00-1.18, p = 0.058)<br>Feeling lonely (OR = 0.96, 95%CI, 0.81-1.14, p = 0.626) | |
| **Ganem et al., 2023** | **October 2021:**<br>Concern with the time to develop the vaccine (63.6%)<br>Concern about side effects (50.9%)<br>Necessity for more information before deciding to vaccinate (45.5%).<br>Already had Covid (16%*)<br>Belief that Covid vaccine does not protect against the disease (35%*) | Univariate analysis.<br>Negatively associated with vaccine acceptance:<br>Perception it is easy to avoid a SARS-CoV-2 infection (OR = 0.29, CI 95% 0.09-0.88, p = 0.029)<br>Use of herbal supplements/homeopathies to avoid SARS-CoV-2 infection - Yes/No (OR = 0.22, CI 95%, 0.08-0.63, p = 0.004) | Univariate analysis.<br><br>Not associated with vaccine acceptance:<br><br>Perceived risk to be infected with Covid-19 - Unlikely/ Very Likely (OR = 3.93, CI 95% 0.51 - 29.96, p = 0.188)<br>Perceived severity if infected with Covid-19 (won't be very sick / will be very sick) (OR = 1.96, CI 95% 0.43-8.92, p = 0.384) | I / B |

| | | | | | | |
|---|---|---|---|---|---|---|
| | | | Distrust in pharmaceutical co's (26%*)<br>Belief there is no risk from Covid-19 (19%*)<br>Preference for natural therapies (21%*)<br>Distrust in vaccines (15%*)<br>Belief that Covid-19 does not exist (15%*)<br>**January 2022:**<br>Concern about side effects (68.4%)<br>Time to develop the vaccine (63.2%)<br>Previous COVID-19 disease (42.1%)<br>Belief that vaccine does not protect against disease (37%*)<br>Distrust in pharmaceutical co's (31%*)<br>Need more information (26%*)<br>Belief there is no risk from Covid-19 (21%*)<br>Preference for alternative/natural therapies (21%*)<br>Distrust in vaccines (11%*)<br>Belief that Covid-19 does not exist (0%) | Positively associated with vaccine acceptance:<br>Higher *self-perceived* knowledge low/high (OR = 3.58, CI 95% 1.27-10.11, p = 0.017)<br>Belief that adherence to vaccination strategies is important no/yes (OR = 15.23, CI 95% 5.13-45.19), p = <0.001)<br>Belief that routine vaccination behaviour is important no/yes (OR = 5.49, CI 95% 2.08 - 14.49, p = 0.001) | Perception about your current mental health (OR = 1.4, CI 95% 0.51 - 3.86, p = 0.514)<br>Factual knowledge about Covid-19 Low / High (OR = 2.49, CI 95% 0.93-6.67, p = 0.069)<br>Preventive behaviour to avoid Covid-19 in last 7 days no / yes (OR = 2.88, CI 95% 0.78 - 10.53, p = 0.111) | |
| **Ganczak et al., 2021** | "Covid-19 is a serious disease" pre-intervention / post-intervention:<br>Males:<br>Agree 69.9% / 81.3%<br>Disagree 19.9% / 12.5% | "I would vaccinate myself against SARS-CoV-2 if a vaccine were available" pre-intervention / post-intervention:<br>Males:<br>Agree 60.2% / 62.5%<br>Disagree 18.2% / 16.5%<br>Not sure - 21.6% / 21.0% | | | | A/I |

Not sure 10.2% / 6.3%
Positive shift 18% / Negative shift 4%
p < .0001

Females:
Agree 74.1% / 83.8%
Disagree 14% / 4.4%
Not sure 10.5% / 6.7%
Positive shift 19% / Negative shift 5%
p < .0001

Life sciences ppts:
Agree 76.6% / 90.2%
Disagree 13.6% / 4.5%
Not sure 9.8% / 5.3%
Positive shift 18% / Negative shift 2%
p < .0001

"Covid-19 Doesn't Concern Me" pre-intervention / post-intervention:
Males
Agree 17.0% / 11.4%
Disagree 71.0% / 83.0%
Not sure 11.9% / 5.7%
Positive shift 20% / Negative shift 8%

Positive shift 11% / Negative shift 11%
p = 0.38

Females:
Agree 49.1% / 53.2%
Disagree 22.5% / 18.7%
Not sure 28.4% / 28.1%
Positive shift 11% / Negative shift 3%
p = 0.004

Life sciences ppts:
Agree 55.3% / 60.6%
Disagree 17.6% / 14.1%
Not sure 27.1% / 25.3%
Positive shift 12% / Negative shift 3%
p = 0.0003

| | | | | | | |
|---|---|---|---|---|---|---|
| | p = 0.0001<br><br>Females<br>Agree 9.9% / 9.4%<br>Disagree 73.4% / 83.3%<br>Not sure 16.7% / 7.3%<br>Positive shift 20% / Negative shift 16%<br>p < 0. 0001<br><br>Life Sciences:<br>Agree 9.3% / 8%<br>Disagree 76.6% / 86.2%<br>Not sure 14.1% / 5.9%<br>Positive shift 31% / Negative shift 9%<br>p < 0.0001 | | | | | |
| **Garg et al., 2024** | | To protect self (78%)<br><br>To protect family/friends (74%)<br><br>To protect community (25%)<br><br>Other (7%) | Do not trust vaccine (57%)<br><br>Question efficacy (21%)<br><br>Worried about side effects (18%)<br><br>Parent/guardian decision (39%)<br><br>Religious (12%)<br><br>Other (11%) | Prior flu vaccine (OR = 1.39, nrCI, 1.01-1.92, p = 0.04) | n/a | B |

| | | | | | |
|---|---|---|---|---|---|
| **Gewirtz-Meyden et al., 2022** | | Do not trust drug companies to make sure vaccine will be safe (56.1%)<br>Virus not dangerous (53.7%)<br>Do not believe in the safety of the vaccine in the short term (51.2%)<br>Prefer not to put drugs or chemicals in their bodies (51.2%)<br>Do not trust government that the vaccine will be safe (41.5%)<br>Do not believe the vaccine is effective (34.1%)<br>Do not believe I should get vaccinated if others get vaccinated (34.1%)<br>I recovered from Covid-19 and I am immune (22%)<br>Most of my friends were not vaccinated (17.1%)<br>2.4% Religious reasons<br>17.1% Other reasons | Vaccinated significantly more likely to say vaccination was joint decision (with parents) (71.9% vs. 54.0%), whereas vaccinated less likely (10.4%) than unvaccinated (28.0%) to say decision was solely their parents ($\mathbf{x}^2 = 7.8$, p = 0.02)<br><br>Logistic regression, bivariate analysis, associated with vaccine acceptance:<br>Both parents vaccinated (OR = 7.1, CI 95% 2.7-18.4, p < .001)<br><br>More time spent on social media networks (OR = 1.3, CI 95% 1.0 - 1.5, p = .02)<br><br>Use of some social media platforms:<br>Facebook (OR = 3.1, CI 95% 1.4-6.7, p = .004)<br>Instagram (OR = 2.8, CI 95% 1.3-6.1, p = .02)<br><br>Distress over effects of the vaccine (OR = 0.7, CI 95% 0.6-0.8, p < .001) | Logistic regression, bivariate analysis.<br><br>Friends/family outside home tested positive (OR = 0.5, CI 95% 0.2-1.0, p = .07)<br>Fear another outbreak (OR = 1.0, CI 95% 0.9-1.1, p = .89)<br>Subjective wellbeing (OR = 1.1, CI 95% 0.9-1.2, p = .09)<br>Depressive/anxiety symptoms (OR = 1.0, CI 95% 0.9-1.1, p = .92)<br>Social support (OR = 1.0, CI 95%0.98-1.03, p = .81)<br>Health (OR = 0.8, CI 95% 0.5-1.3, p = .43)<br>Use of some social media platforms:<br><br>TikTok (OR = 0.9, CI 95% 0.5-1.9, p = 0.89)<br>Snapchat (OR = 0.9, CI 95% 0.5 - 2.0, p = 0.98)<br>YouTube (OR = 0.7, CI 95% 0.3-1.5, p = 0.37)<br>Twitter (OR = 2.1, CI 95% 0.7-6.7, p = 0.21) | B |
| Guneysu et al., 2023 | If I get sick, I will have a lighter disease course (46.5%)<br><br>I fulfil my social responsibility (25.8%) | Due to vaccine side effects (5.8%)<br><br>Distrust of vaccine content (2.6%) | n/a – sociodemographic only | n/a – sociodemographic only | |

| | | | | |
|---|---|---|---|---|
| | I will not get infected again (12.2%)<br><br>Reduces my fear of getting sick (10.1%)<br><br>Parents' request (8.7%)<br><br>To be free to go abroad (4.2%)<br><br>Other (0.1%) | Don't think the vaccine will work (2.5%)<br><br>Needle – afraid of injections (2.3%)<br><br>May be allergic to vaccine (2.2%)<br><br>Would rather have the disease and be immune (1.9%)<br><br>Vaccine is new & not reliable (1.7%)<br><br>Already had disease and am immune (1.4%)<br><br>If I get ill, it will be light (0.9%)<br><br>I am healthy, my risk of infection is low (0.5%)<br><br>I think it is a bioweapon (0.5%)<br><br>Religious disapproval (0.1%)<br><br>Other (0.4%) | | |
| Hasanatulu dhhiyah et al., 2023 | | | Binary logistic regression:<br><br>Covid-19 module (OR = 0.60, 95%CI, 0.22-1.64, p = 0.326) | I/B |
| **Hopfer et al., 2022 (Quantitati** | Confidence in vaccine safety - 99<br>Social benefit -89<br>Confidence in vaccine efficacy - 82 | Concerns over potential long term side effects - 128<br>Concerns over potential short term side effects or reactions - 99 | | I |

| | | | | | | |
|---|---|---|---|---|---|---|
| **ve only included)** | | It is important to protect others 73<br>A HCW tells me it is good to vaccinate - 63<br>Important others think it is good to vaccinate - 42<br>Confidence in how the vaccine was developed - 38<br>No cost - 32<br>Not worried about short term side effects - 27<br>Not worried about long term side effects - 23<br>Easy access - 22<br>I think Covid-19 can make me really sick - 22<br>I think I may be at high risk of getting Covid-19 - 22<br>Not worried about the ingredients in the vaccine - 14<br>Certain about vaccine benefits - 13<br>Having enough information or access to information - 9<br>Personal allergies - 8<br>Personal medical history - 8<br>Social media or internet - 3<br>*NB Ranking scores not %ages* | Concerns over how well the vaccine works - 59<br>Fear of needles - 51<br>Concerns over vaccine safety - 51<br>Not enough vaccine information or a lack of access to vaccine information - 44<br>Concerns over the ingredients that make up the vaccine - 44<br>COVID-19 isn't a very severe disease for kids - 38<br>Trust in how the vaccine was developed - 31<br>Personal allergies -22<br>Uncertainty about vaccine benefits - 21<br>Low perceptions of risk of getting COVID-19 - 20<br>Cost - 15<br>COVID-19 isn't a very severe disease for adults - 15<br>Fear of receiving too many vaccines - 14<br>Personal medical history - 11<br>Lack of support by important others - 11<br>Social media or Internet stories or comments - 8<br>Preference for natural immunity - 4<br>I've already had COVID-19 - 1<br>*NB Ranking scores not %ages* | | | |
| **Hwang et al., 2023** | | | | Correlations:<br><br>Correlated with intention to vaccinate: | Chinese Health Questionnaire (psychiatric morbidity) (r = 0.062, p = nr) | I |

| | | | | |
|---|---|---|---|---|
| | | Impact of event scale (intrusive thoughts/avoidance behaviours) (r = 0.214, p = 0.012) | | |
| **Inaba et al., 2022** | | | Binomial regression:<br>Don't want to be vaccinated not significantly associated with self-assessment of health status in any group:<br>Japan boys: r = 0.694, OR = 2.001, CI95% (-0.270-1.622), p = nr<br>Russia boys: r = -0.299, OR = 0.741, CI95% (-1.068-0.476), p = nr<br>Japan girls: r = 0.661, OR = 1.936, CI95% (-0.226-1.539),p = nr<br>Russia girls: r = 0.912, OR = 0.901, CI95% (-0.726-0.529), p = nr | I |
| **Janssen et al., 2023** | Aged 12-15 years:<br><br>Do not trust vaccine (28%)<br><br>Admitted to hospital (4%)<br><br>Forgot (4%)<br><br>Too soon after surgery (4%)<br><br>Upcoming emigration (4%)<br><br>Could not be reached (32%)<br><br>No specific reason (16%) | | | B |

| | | |
|---|---|---|
| | Aged 16-17 years:<br><br>Fear of needles (4%)<br><br>Do not trust vaccine (13%)<br><br>Taking post-op medication (4%)<br><br>Could not be reached (17%) | |
| **Kajiwara et al., 2022** | Would not get vaccinated / would not accept 3rd dose:<br>I would be concerned about side effects from the vaccine (29.5%) / (33.3%)<br>I am afraid/anxious (25%) / (0%)<br>I do not like needles/injections (20.5%) / 0%<br>I would be concerned about getting infected with coronavirus/other disease/worried about vaccine safety (15.9%) / (11.1%)<br>I am allergic to vaccines/have other physical reasons (11.4%)<br>I am not concerned about Coronavirus/I do not need it 6.8% / (7.4%)<br>I will not have time to get vaccinated 2.8% / 0%<br>I do not think vaccines work very well 2.3% / 0%<br>It depends on what people around me do / there is not enough information to make a decision - / 33.3% | I / B |

| | | | | | |
|---|---|---|---|---|---|
| **Klein et al 2021*** | **(see Middleman et al., 2022a)** | | | | |
| **Lee et al., 2022** | Perceived level of Covid-19 knowledge:<br><br>Know well/very well (32.9%)<br>Perceived safety of Covid-19 vaccines?<br><br>Safe/very safe (51%)<br><br>Not safe/very not safe – (24.2%)<br><br>Unsure (24.8%)<br>Perceived efficacy:<br><br>Very effective/effective (57.3%)<br><br>Ineffective/not at all (10.4%)<br><br>Unsure (32.4%)<br>How important?<br>Important/very important (47.6%)<br>Neutral (30.8%)<br><br>Not important at all (9.2%) | Prevent infection for oneself (62%*)<br>Prevent transmission family and friends (58%*)<br>Return to normal lifestyle (30%*)<br>Prevent transmission in community (12%*)<br>Relieve public health measures (15%) | Concerns for safety issues (90%*)<br>Do not want interruptions in school and studies (30%*)<br>Do not want shot (18%*)<br>Low risk of infection (20%*)<br>Public health measures sufficient for prevention (20%*) | Multivariate regression:<br>Associated with intention to vaccinate:<br>Perception of safety (OR = 4.09, CI95% 3.96-4.22), p = <.001)<br>Perception of effectiveness (OR = 2.24, CI95% 2.17-2.32, p = <.001)<br>Perception of risk-benefit (OR = 1.75, CI95% 1.72-1.78, p = <.001)<br>Perceived risk of Covid-19 infection (OR = 1.14, CI95% 1.12-1.17, P = <.001)<br>Perceived severity of Covid-19 infection<br>Self-health perception (OR = 1.12, ci95% 1.10-1.13), P = <.001)<br><br>Related to vaccine hesitancy:<br>Self perceived knowledge OR = 0.96, CI95% 0.95-0.98, p = <.001) | I |

| | | | | | | |
|---|---|---|---|---|---|---|
| | Unsure (12.4%)<br>Risk-benefit?<br><br>Risks higher/much higher (16.5%)<br><br>Equal (38.1%)<br><br>Benefits higher/much higher (28.9%)<br><br>Unsure (16.5%) | | | | | |
| **Li et al., 2022a** | | Totally own decision, no specific reason (70.6%)<br>Consulted the scientific literature (10.8%)<br>Encouraged by family members (11.6%)<br>Encouraged by teachers (10.5%)<br>Encouraged by friends (5.2%)<br>Encouraged by doctors (2.9%)<br>Encouraged by propaganda News (5.1%)<br>Requested by School manager (14.6%)<br>Unwilling to, but followed school arrangement (3.0%)<br>Others (6.5%)<br>Confidence in vaccine effectiveness (in vaccinated) 95% (in not vaccinated) 93.6%<br><br>Life is affected by Covid-19 - seriously affected/affected but not seriously (in vaccinated) 64.1% (in non-vaccinated 58.1%) | Not willing to without reason 4 (13.3%)<br>Not willing to, but with the excuse such as toothache or cold 1 (3.3%)<br>Worried about the efficacy of the vaccine 2 (6.7)<br>Worry about the short-term protection (3–6 months) 2 (6.7)<br>Worry about the vaccine safety 1 (3.3)<br>Worry about political involvement 2 (6.7)<br>Influenced by parents or relatives 1 (3.3)<br>Influenced by friends or teachers 0 (0.0)<br>Influenced by Internet 0 (0.0)<br>Worry about the safety without own chronic diseases 2 (6.7)<br>Worry about the safety because of own chronic diseases 5 (16.7)<br>Willing to take, but declined by vaccination staff 6 (20.0) | Univariate analysis.<br>Acceptance of Covid-19 vaccine associated with:<br>Willingness for vaccination (p = .001) | Univariate analysis:<br>Not associated with vaccine acceptance:<br><br>Worry about vaccine safety (p = 0.131)<br>Confidence in vaccine effectiveness (p = 0.668)<br>Is life affected by Covid-19 (p = 0.151) | B |

| | | |
|---|---|---|
| | Others 3 (9.3)<br>Worry about vaccine safety (in vaccinated) 15.5%, in non-vaccinated (25.8%) | |
| **Li et al., 2022b** | | Logistic regression, univariate analysis:<br>Reasons for Covid-19 vaccine acceptance (not vaccinated/vaccinated)<br>Worry about vaccine safety:<br>Worry 25.8% / 15.5%, No worry 74.2% / 84.5%, p = .131<br>Confidence for vaccine effectiveness:<br>Confident 93.6% / 95%, Not confident 6.4%, 5%, p = .668<br>Willingness for vaccination:<br>Yes 87.1% / 97.1%, No 12.9% / 2.9%, p = .001<br><br>Logistic regression, multivariate analysis:<br>Associated with willingness to accept third dose:<br>Worry about vaccine safety v no worry (B = 1.04, SE = 0.24, OR = 2.82, 95%CI 1.75-4.54, p < .001<br>Confidence in vaccine effectiveness v no confidence (B = -1.43, se = 0.32, OR = 0.24, 95%CI 0.13-0.45, p < .001)<br>Parent's opinion of children's vaccination v support:<br>Neutral (B = -1.35, SE = 0.38, OR = 0.26, 95%CI 0.12-0.55), p = .001)<br>Opposed (B = -2.10, SE = 1.00, OR = 0.12, 95% 0.02-0.87, p = 0.036)<br>Unknown (B = -1.15, SE = 0.50, OR = 0.32, 95%CI 0.12-0.84, p = 0.021) |

| | | | | | |
|---|---|---|---|---|---|
| **McKinnon et al., 2023** | Vaccinated agree with following statements:<br>Vaccines against COVID-19 are safe (46.9%)<br>Vaccines are an effective way to reduce the risk of contracting COVID-19 (60.6%)<br>I distrust COVID-19 vaccines because they were developed too quickly (28.6%)<br>By being vaccinated against COVID-19, I am protecting myself from the disease (70.4%)<br>Those who have had COVID-19 do not need to be vaccinated (24.8%)<br>Pharmaceutical companies cover up the dangers of vaccines (20.6%)<br>The government is trying to cover up the link between vaccines and autism (12.2%)<br>Most of my friends and family members are vaccinated against COVID-19 (82.7%)<br>Vaccination against COVID-19 goes against my religious beliefs or personal principles (11.2%)<br>Only those at risk of becoming seriously ill from COVID-19 need to be vaccinated (24.2%)<br>By being vaccinated against COVID-19, I am helping to protect the health of others in my community (77.6%)<br>Physical distancing, frequent handwashing and wearing a mask are enough to protect me from COVID-19 (36.6%)<br>The severity of the pandemic has been overstated (34.6%)<br>COVID-19 vaccination should be | Unvaccinated - agree with following statements:<br>Vaccines against COVID-19 are safe (10%)<br>Vaccines are an effective way to reduce the risk of contracting COVID-19 (27.8%)<br>I distrust COVID-19 vaccines because they were developed too quickly (44.3%)<br>By being vaccinated against COVID-19, I am protecting myself from the disease (32.1%)<br>Those who have had COVID-19 do not need to be vaccinated (30.4%)<br>Pharmaceutical companies cover up the dangers of vaccines (35.4%)<br>The government is trying to cover up the link between vaccines and autism (12.7%)<br>Most of my friends and family members are vaccinated against COVID-19 (60.8%)<br>Vaccination against COVID-19 goes against my religious beliefs or personal principles (10.3%)<br>Only those at risk of becoming seriously ill from COVID-19 need to be vaccinated (29.1%)<br>By being vaccinated against COVID-19, I am helping to protect the health of others in my community (37.2%)<br>Physical distancing, frequent handwashing and wearing a mask are enough to protect me from COVID-19 (48.1%)<br>The severity of the pandemic has been overstated COVID-19 | Logistic regression:<br><br>Associated with vaccine acceptance:<br>Confidence in the vaccine: they were more likely to report that they agree (vs. disagree or do not know) that the vaccine is safe (44% vs. 13%, $p < .001$) and effective (58% vs. 33%, $p < .001$).<br>Collective responsibility, considering vaccination as a means of protecting the health of others in the community (76% vs. 42%, $p < .001$).<br>Friends/family were vaccinated 83% v 62%, p = <.001<br>Greater support for vaccine passports (47% v 38%, $p = .01$) | complacency toward vaccination e.g., believing only those at risk of serious illness need to be vaccinated 24% v 29%<br>constraints of daily life (13% v 17%) influenced vaccination decisions did not differ significantly by vaccination status | B |

| | | | | | | |
|---|---|---|---|---|---|---|
| | | mandatory for health care workers (64%)<br>The constraints of daily life might prevent me from getting vaccinated against COVID-19 (12.7%)<br>The vaccine passport is a good strategy to encourage people to get vaccinated (48.6%)<br>It is a good idea to make unvaccinated people pay an additional tax (health contribution) to fund the healthcare system (24.8%)<br>People in my neighbourhood can be trusted (50.9%)<br>I have a strong sense of belonging to Quebec (53.7%) | (41.8%)<br>Vaccination should be mandatory for health care workers (31.6%)<br>The constraints of daily life might prevent me from getting vaccinated against COVID-19 (16.7%)<br>The vaccine passport is a good strategy to encourage people to get vaccinated (32.9%)<br>It is a good idea to make unvaccinated people pay an additional tax (health contribution) to fund the healthcare system (8.9%)<br>People in my neighbourhood can be trusted (43%)<br>I have a strong sense of belonging to Quebec (36.7%) | | | |
| **Meraya et al., 2022** | Interested in Covid vaccine? Yes (73.5%)<br>Are Covid vaccines 100% effective?<br><br>Yes (11.4%)<br><br>No (66.7%)<br><br>Dk (22%)<br>Are Covid-19 vaccines safe?<br><br>Yes (59.1%)<br><br>No (8.3%) | | | n/a - multivariate regression for adult data only | n/a - multivariate regression for adult data only | A |

DK (32.6%)
Do Covid vaccines reduce risk of disease?

Yes (67.4%)

No (16.7%)

DK (15.9%)
Do Covid vaccines lessen severity of symptoms?

Yes (71.2%)

No (9.1%)

DK (19.7%)
Was getting vaccinated a top priority for you? Yes (66.7%)

No (29.5%)

DK (3.8%)

"I am worried about the potential side effects of COVID-19 vaccinations in children": (53.3% agree/ strongly agree)

"I am worried about the potential side effects of COVID-19 vaccinations in pregnant women": 55.5% agree/strongly agree

| | | | | | |
|---|---|---|---|---|---|
| | “I am worried about the long-term side effects of COVID-19 vaccinations”: 53% agree/strongly agree | | | | |
| **Middleman et al., 2022a** | Agree / strongly agree: Wave 1/2/3:<br>Covid-19 is a serious disease (83%/82%/85%)<br>it's possible I will get infected with covid-19 - 60% / 59% / 57%<br>it's important for all teens to get the recommended vaccines - 72% / 73% / 74%<br>If my friends get vaccinated, I will get vaccinated too (58%/54%/53%) | Agree / strongly agree: Wave 1/2/3:<br>I want to protect myself (53% / 49% / 51%)<br>I want to protect everyone in my family (47% / 52% / 44%)<br>Vaccination is the best way for me to avoid a potentially serious disease (37%/38% /38%)<br>I want to help protect my community (35%/36%/34%)<br>Life will not get back to normal until most people are vaccinated including teens (37%/35% /34%)<br>I would be safe around other people (38%/39%/37%)<br>A family member is at high risk for Covid-19 (31%/35%/30%) | %age choosing agree / strongly agree:<br>I have some concerns about the safety of vaccines - 56% / 64% / 62%<br>I have some concerns about the effectiveness of some vaccines - 55% / 64% / 63%<br>What I have read on social media has concerned me about the safety of some vaccines - 50% 56% / 57%<br><br>Wave 1 / 2 / 3:<br>Concerned about possible side effects - 40% / 47% / 58%<br>Concerned I could get Covid from the vaccine - 23% / 20% / 15%<br>I do not think the Covid vaccine will work well - 17% / 20% / 15%<br>I do not like getting shots / needles - 20% / 20% / 21%<br>I think the Covid-19 outbreak is not as serious as some say - 12% / 14% / 14%<br>I think teens do not get seriously ill from Covid-19 - 11% / 16% / 19% | | A / B / I |

| | | | | |
|---|---|---|---|---|
| **Middleman et al., 2022b*** | **(see Middleman et al., 2022a)** | | | |
| **Moore et al., 2024** | | *To protect self (80.8%)<br><br>*To protect others (81.3%)<br><br>*I want to return to normal life (11.5%)<br><br>To protect my family/household (83.5%)<br><br>My parents want me to be vaccinated (57.5%)<br><br>No reason not to be (57.3%)<br><br>Did not want to go against my parents (42.3%)<br><br>Required by my school/job (29.3%)<br><br>I believe that the vaccine will work better if more people get them (64.3%)<br><br>Vaccination required to take part in sports/extra-curricular activities at school (31.3%)<br><br>I trust vaccine science (60.8%)<br><br>Other (13%) | *We do not know about long-term effects of vaccine (57.7%)<br><br>*Concern about side effects (56.6%)<br><br>*I do not like needles/shots (45.2%)<br><br>My parents did not want me to be vaccinated (28.9%<br><br>I do not trust the government (41.9%)<br><br>I do not need the vaccine (36.1%)<br><br>I did not want to go against my parents (30.0%)<br><br>Not sure the vaccine is safe (38.1%)<br><br>Other people need it more (32.8%)<br><br>Confused about whether I was eligible (22.7%)<br><br>Concerned about cost of vaccines (24.0%) | I / B |

| | | | | |
|---|---|---|---|---|
| | *Ranked importance 1, 2 3 respectively | Do not trust healthcare system (27.9%)<br><br>Concerned about immigration status (14.3%)<br><br>Other (7.9%)<br><br><br>Biggest barriers<br><br>Parent preference (43.3%)<br><br>I had little or no free time to get vaccinated (38.73%)<br><br>Few or not vaccination sites available (13.3%)<br><br><br>*Ranked importance 1, 2 3 respectively | | |
| **Nilsson et al., 2021**<br><br>**(Quantitative)** | | | Multivariate analysis.<br><br>Associated with willingness to get vaccine:<br>Higher anxiety (ORa = 2.09, CI 95% 1.26-3.45) | I |
| **Oka et al., 2022** | Vaccine knowledge:<br>Not 100% effective (98.3%)<br>Minor side effects (97.6%)<br>Free (96.3%)<br>Effective after two doses (94.6%)<br>Does not modify genes (92%) | Long term risk to health (38.7%)<br>Vaccine efficacy (37.4%)<br>Fear turning into a carrier (21.3%) | n/a | I |

| | | | | | |
|---|---|---|---|---|---|
| | | Awareness of side effects/concern about side effects:<br>Anaphylactic shock - (18.9%/73.2%)<br>Fast heartbeat (19.6%/58%)<br>Low blood pressure - (14.3%/58.3%)<br>Fever (89.1%/44.1%)<br>Nausea (55.9%/31.5%)<br>Headache (78.7%/31.5%)<br>Muscle pain (73.5%/21.1%)<br>Redness / swelling - (55.9%/17.2%)<br><br>Chills (45.0%/17.4%)<br>Tiredness (80.0%/16.3%) | Feel it is not necessary (17.0%)<br><br>Fear of needles (16.1%)<br>Negative past experience (15.7%)<br>Fear it may affect swab test results (12.1%)<br>Unsure of application procedures (9.3%) | | |
| **Parlak & Ener, 2022** | Yes / No / Unsure<br><br>Vaccine refusal is a problem that poses threat to the entire community (57.9%/36.7%/5.4%<br>People can be forced to get vaccinated (52.8%/43.9/3.3%)<br>Anti-vaccination arguments influence me (28.7%/67.1%/4.2%)<br>Vaccination is necessary for public health (67.8%/22.9%/9.3%)<br><br>I try to persuade vaccine-hesitant people around me to | | Concerns about adverse effects of vaccines (25.7%)<br>Negative comments of others on vaccines (18.8%)<br>I think the COVID-19 vaccine is ineffective (18.2%)<br>Believing the immune system will fight off the disease (11.6%)<br>Negative opinions of experts regarding vaccines (9.6%)<br>Waiting for a Turkish COVID-19 vaccine to be available (4.6%)<br><br>I think there are no adequate studies on COVID-19 vaccines (4.3%) Vaccines are manufactured abroad (3.0%)<br>Religious concerns regarding vaccine content (3.3%)<br>I think that there is no such | ANOVA / Correlation:<br><br>Associated with vaccine hesitancy:<br><br>Primary source of information /vaccine hesitancy scale in pandemics:<br>Physicians / HCW - 13.3% - 28.6 +/- 4.2<br>Media/internet - 70.7% - 29.9 +/- 4.7<br>Other (neighbours, friends, relatives, religious leaders) - 16.0% - 32.7 +/- 4.8<br>p = 0.001<br>Media outlets influencing decision about vaccine:<br>TV, radio, newspapers, magazines - 60.3% - 29.5 +/- 4.7<br>Social media - 39.7% - 31.2 +/- 4.7 | I |

| | | | | | | |
|---|---|---|---|---|---|---|
| | get vaccinated<br>(62.3%/29.0%/8.7%) | | disease as COVID-19 (3.0%)<br>I am waiting for others to get vaccinated first (2.6%)<br>Negative opinion of a physician/healthcare provider about the COVID-19 vaccine (2.3%)<br>Fear of injection (2.0%)<br>Considering vaccination as some plot of foreign countries (e.g., secretly implanting microchips via vaccination) (0.7%) | p = 0.003<br>Believing anti-vaccine content circulating on social media:<br>Yes - 12.6% - 34.9 +/- 4.8<br>No - 63.4% - 29.0 +/- 4.5<br>Unsure - 3.7% - 33.2 +/- 5.5<br>p = 0.001<br>Influenced by anti-vaccine rhetoric:<br>Yes - 27.1% - 31.9 +/- 4.8<br>No - 63.4% - 29.0 +/- 4.5<br>Unsure - 4% - 33.2 +/- 2.9<br>p = 0.001 | | |
| **Persaud et al., 2023** | n/a | | Lack of personal utility/benefit (53%)<br>Mistrust / misinformation / rushed (40%)<br>Unanswered medical questions (7%) | n/a - sociodemographic only | n/a - sociodemographic only | I / B |
| **Pimental et al., 2022** | n/a | | | Bivariate analysis:<br>Associated with perception of Covid-19 threat to health:<br><br>Health literacy (b = 0.041, CI 95% 0.006-0.076, p = 0.010)<br><br>Associated with intention **not** to vaccinate:<br>Health literacy (PR = 0.95, CI 95% 0.91-0.98, p = 0.003) | Multivariate analysis:<br><br>Associated with intention **not** to vaccinate:<br>Health literacy (PR = 0.97, CI 95% 0.93-1.01, p = 0.091) | I |
| **Qian et al., 2023** | | Very likely to choose for themselves (66.7%)<br>Get most info about healthcare from my parents/guardians (65.3%)<br>Peers are main sources of health information (12.5%) | | Correlation:<br><br>Positive correlation between:<br>Opinion of Covid-19 vaccine and likelihood of choosing vaccine for self (without parental consent) (r = 0.91)<br>Parental covid-19 vaccine positive | | I |

| | | | | | | |
|---|---|---|---|---|---|---|
| | | Social media is main source of health information (13.9%) | | opinions and adolescent vaccine positive opinions (r = 0.82)<br>Being satisfied with how parents communicate covid-19 vaccine decisions and agreeing with parents (r = 0.75)<br>Feeling safe in asking parents about controversial health topics and agreeing on health topics (r = -.74) | | |
| **Rehati et al., 2022** | Vaccination concerns:<br>Resistant / hesitant / willing:<br>I am afraid of Sars-Cov-2 transmission:<br>Agree (7% / 27.7% / 65.3%)<br>Not sure (4.6% / 9.8% / 25.6%)<br>Disagree (20.8% / 27.2% / 52.0%)<br>I have a potential risk of being infected with Sars-Cov-2:<br>Agree (4.7% / 20.6% / 74.7%)<br>Not sure (4.6% / 47.9% / 25.6%)<br>Disagree (15.7% / 30.8% / 53.5%) | | Major concerns that affect my Covid-19 vaccination decision:<br>Resistant / Hesitant / Willing:<br>Safety (8% / 31.3% / 60.7%)<br>Effectiveness (7.5% / 30.7% / 61.8%)<br>Price (6.1% / 31.3% / 62.7%)<br>Convenience (6.0% / 29.0% / 65.0%)<br>Doctors' recommendation (6.7% / 29.1% / 64.2%) | Multinomial logistic regression:<br>Vaccine resistance v willing:<br>I am afraid of Sars-cov-2 transmission - Disagree (OR = 1.35, 95%Ci 2.86-4.46, p = 0.013) i.e. students less afraid of Covid-19 more likely to resist<br>Did not know much about Covid-19, more resistant (OR = 2.09; 95% CI: 1.70–2.57)<br>Do not perceive self to be susceptible to risk of infection - Disagree (OR = 3.57, 95%Ci 2.86-4.46, p <.001) and unsure (OR = 1.43, 95%CI 1.08-1.89, p = 0.012) both more vaccine resistant<br>Covid-19 will have no impact on my life / unsure about impact (OR = 3.24; 95% CI: 2.56–4.11)<br>Major concern: efficacy (OR = 0.66, 95%CI 0.52-0.83, p < .001)<br>Had experience of prior influenza vaccination:<br>Not sure (OR = 1.30, 95%CI 1.01-1.67, p = 0.038)<br>Disagree (OR = 1.57, 95%CI 1.25-1.98, p < .001)<br><br>Vaccine hesitancy v willing:<br>Afraid of Sars-Cov-2 transmission: | Multinomial logistic regression.<br><br>Vaccine resistant v willing:<br>I am afraid of Sars-Cov-2 transmission - not sure (OR = 0.87, 95%Ci 0.58-1.29, p = 0.487<br>Major concerns that affect my vaccination:<br>Safety (OR = 0.80, 95%CI 0.58-1.10, p = 0.167)<br>Price (OR = 0.86, 95%CI 0.69-1.07, p = 0.169)<br>Convenience (OR = 0.81, 95%CI 0.64-1.01, p = 0.064)<br>Doctors recommendation (OR = 0.93, 95%CI 0.75-1.15, p = 0.496)<br><br>Vaccine hesitant v willing:<br>I am afraid of Sars-Cov-2 transmission - Disagree (OR = 0.93, 95%CI 0.78-1.12, p = 0.458)<br>Major concerns that affect my vaccination: Safety (OR = 1.20, 95%CI 0.95-1.50, p = 0.130) | I |

| | | | |
|---|---|---|---|
| | | Not sure (OR = 2.09, 95%CI 1.70-2.57, p < .001) *NB Interpreted as "did not know much about Sars-Cov-2"*<br>I have a potential risk of being infected with Sars-Cov-2: Not sure (OR = 1.93, 95%CI 1.67-2.22, p < .001), Disagree (OR = 1.72, 95%CI 1.50-1.97, p < .001)<br>Major concerns that affect my vaccination:<br>Effectiveness (OR = 0.84, 95%CI 0.72-0.98, p = 0.027) i.e. More effective vaccine perceived to be - less hesitant<br>Price (OR = 1.20, 95%CI 1.05-1.36, p = 0.007)<br>Convenience (OR = 0.84; 95% CI: 0.73–0.96) - convenience reduced vaccine hesitancy<br>Doctors' recommendation (OR = 0.86; 95% CI: 0.76–0.98) Drs rec reduced vaccine hesitancy<br>Had experience of prior influenza vaccination:<br>Not sure (OR = 1.44, 95%CI 1.24-1.67, p < .001)<br>Disagree (OR = 1.33, 95%CI 1.14-1.55, p < .001) | |
| **Rogers et al., 2021** | Frequency counts for vaccine concern items on 5-point scale:<br>Vaccine not tested enough (M = 3.12, SD = 1.38)<br>Worried about side effects (M= 3.32, SD = 1.34)<br>Might get Covid from the vaccine (M = 2.38, SD = 1.42)<br>Will wait for others to get the vaccine (M = 2.95, SD = 1.47)<br>The vaccine might not be effective (M = 2.44, SD = 1.37) | T-Tests:<br>Yes/already v no:<br>Covid anxiety (M = 2.32, SD = 0.91 v M = 2.32, SD = 0.80, p < .001, d = .58)<br>Concern- safety/efficacy (M = 2.35, SD = 0.99 v M = 3.54, SD = 1.00, p < .001, d = -1.19)<br>Concern - necessity (M = 1.76, SD = 0.94 v M = 3.17, SD = 1.14, p < .001, d = -1.41)<br>Friend norms (M = 3.61, SD = 1.11 v M = 1.92, SD = 0.13, p < .001, d | I / B |

Necessity concerns:
Covid-19 is mild (M = 2.33, SD = 1.36)
My age group is not at risk (M = 2.14, SD = 1.31)
Natural immunity is better (M = 2.31, SD = 1.33)

= 1.67)
Parent norms (M = 4.17, SD = 0.66 v M = 1.80, SD = 0.85, P < .001, d = 3.26)

Correlations:
Vaccine intention correlates to:
Parent norms (.69, p < .001)
Friend norms (.59, p < .001)
Concern: safety (-.47, p < .001)
Concern: necessity (-.49, p < .001)
Covid anxiety (.22, p < .001)

Hierarchical multiple linear regressions:
Step 3: Covid-19-related anxiety positively associated with vaccine intention - more anxiety, more willingness (B = .20, 95%CI 0.14-0.26, p = .00) and explained 4.6% of the variance in intention to accept vaccine (after sociodemographic variables)

Step 4: Significant negative association between vaccine concerns and intention. More concerns about safety, efficacy, & necessity - less likely to be vaccinated:
Safety/efficacy (B = -.24, 95%CI .14 - .24, p = .000)
Necessity (B = -.21, 95%CI -.26 - .17, p = .000)
This accounted for a further 27% of variance (after sociodemographic and anxiety)
Step 5:Significant positive relationship between friend and parent norms and vaccine intention: Perception of support of and uptake of the vaccine by parents and peers - more willing to be vaccinated.
Friend (B = 0.13, 95%CI .09-.16, p = .000)

| | | | |
|---|---|---|---|
| | Parent (B = .36, 95%CI .32 - .40, p = .000)<br>This accounted for a further 23.4% of variance (after the above)<br>Mediation analysis:<br>parent norms associated with vaccination intentions directly (b = .58, 95%CI .53 - .63), and indirectly through safety and efficacy concerns (b = .04, 95%CI .02 -.05) and doubts about vaccine necessity (b = .04; 95%CI .02 -.06).<br>Peer norms also associated with vaccine intentions directly (b = .16, 95%CI .15 -.20), and indirectly through doubts about vaccine necessity (b = .03, 95%CI .02 -.05) | | |
| **Rosen et al 2022*** | Multivariable logistic regression:<br>Associated with intent to receive Covid-19 vaccination:<br>Positive attitude toward vaccines in general (aOR = 3.55, 95%Ci 2.17-5.80, p < .001)<br>Perceiving Covid-19 vaccine to be safe (aOR = 1.54, 95%CI 1.32-1.81, p < .001) | Knowing enough about Covid-19 vaccine to decide (data nr) | I |
| **Rosen et al., 2024** | Multivariable logistic regression:<br>Positively associated with intent to receive Covid-19 vaccination: | Normative beliefs (nr)<br><br>Sources of information (nr) | |

| | | | | | |
|---|---|---|---|---|---|
| | | | Lower vaccine hesitancy (aOR = 1.50, 95%CI, 1.29-1.76)<br><br>Perception that vaccine is safe (aOR = 2.80, 95%CI, 1.67-4.71)<br><br>Negatively associated:<br><br>Perception that vaccine has serious side effects (aOR = 0.53, 95%CI, 0.32-0.88) | | |
| **Rothoeft et al., 2023** | Concern about contracting Covid-19 (82.3%)<br><br>Access to leisure facilities (76.9%)<br><br>Protecting family members (70.9%) | | | | B |
| **Roy et al., 2023** | Belief that vaccine is safe (75.17%)<br><br>Belief in vaccine efficacy<br><br>(75.63%)<br><br>Am well communicated about vaccine (71.8%)<br><br>Trust (81.37%)<br><br>Culture (usual habit) (79.4%) | Fear of side effects (60.97%)<br><br>Injection anxiety (70.28%)<br><br>Political influence (30.25%)<br><br>Conspiracy belief (12.4%)<br><br>Rumour (50.46%)<br><br>Religious belief (18.4%)<br><br>Negative information (79.99%) | Binary logistic regression:<br><br>Positively associated with vaccine acceptance:<br><br>Belief in vaccine safety (B = 1.125, SE = 0.251, p = <.05)<br><br>Belief in vaccine efficacy (B = 0.262, SE = 0.205, p = <.05)<br><br>Communication (B = 0.085, SE = 0.240, p = <.05) | Religious belief (B = 0.802, SE = 0.243, p = nr)<br><br>Conspiracy belief (B = 0.065, SE = 0.343, p = nr)<br><br>Political influence (B = 0.839, SE = 0.349, p = nr)<br><br>Trust (B = 0.412, SE = 0.516, p = nr)<br><br>Negative information (B = 0.854, SE = 0.345, p = nr) | I |

| | | | | | | |
|---|---|---|---|---|---|---|
| | | Vaccine should be mandated (67.64%) | | Culture (usual) (B = 0.186, SE = 0.265, p < .01)<br><br>Negatively associated with vaccine acceptance:<br><br>Fear of side effects B = -0.503, SE = 0.251, p <.01<br><br>Injection anxiety (B = -0.787, SE = 0.552, p < .01) | Rumour (B = 0.557, SE = 0.283, p = nr)<br><br>Belief vaccine should be mandated (B = 0.587, SE = 0.484, p = nr) | |
| **Ryan et al., 2023** | | Vaccine-acceptant:<br>Protecting the health of family and friends (87.2%)<br>Personal Covid-19 prevention (77.3%)<br>Protecting the health of communities (75.6%)<br>To resume social activities (67.8%)<br>it would allow me to travel (51.2%)<br>I am concerned about the severity of Covid-19 (49.2%)<br>Recommended to me by a family member or friend (42.1%)<br>Help me get back to school (43%)<br><br>Saw people in my community getting vaccinated (34.7%) | Vaccine-hesitant:<br>Concerrn about side effects (45%)<br>Waiting to see if it is safe (43.9%)<br>My parents/caregivers will decide for me (36.7%)<br>Lack of trust in vaccines (28.0%)<br>Other people need the vaccine more (27.5%)<br>Don't like needles (20.7%)<br>Don't know if the Covid-19 vaccine will work (20.7%)<br>Don't believe I need a Covid-19 vaccine (20.0%)<br>Concerned about having an allergic reaction (18.3%)<br>Don't think Covid-19 is a big threat (17.1%)<br>Obstacles prevent me getting a vaccine (8.5%) | Chi-squared:<br><br>Associated with vaccine intention:<br>Concern about Covid-19 infection (hesitant / acceptant):<br>Not concerned (34.1%) / (10%)<br>Slightly concerned (24.2%) / (20.5%)<br>Somewhat concerned (26.2% / 32.3%)<br>Very concerned (15.4%) / (32.3%)<br>x2 = 65.06, p < .001<br><br>Concern about vaccine side effects<br>Not concerned (17.6%) / (24.0%)<br>Slightly concerned (25.9%) / (36.4%)<br>Somewhat concerned (28.5%) / 24.8%) | | I |

| | | | | | |
|---|---|---|---|---|---|
| | Allow me to get back to school (34.3%)<br>Know someone who became ill/died from Covid-19 (27.7%)<br>Recommended by a healthcare provider (15.6%)<br>My school requires it (10.3%)<br>My workplace required it (2.5%) | Other people in my community are choosing not to get vaccinated (6.6%)<br>Concerned about the cost of a Covid-19 vaccine (5.6%) | Very concerned (28.0%) / (14.9%), x2 = 23.06, p < .001. | | |
| **Scherer et al., 2021** | Factors which would increase intention (among unvaccinated):<br>More information about safety (21.7%)<br>More information about efficacy (17.6%)<br>If vaccination was a school requirement (23.9%)<br>Preventing the spread of COVID-19 to family and friends (17.1%)<br>Allowing resumption of or increase in social activities (15.5%), or traveling (14.5%)<br>Recommendation by a health care professional as a factor that<br>would increase vaccination intentions (8.9%)<br>Others* (estimated based on graph, %ages not provided):<br>0-20%: Workplace requirement, family or friend recommendation, able to go back to school, large increase in Covid-19 cases, reduce community spread, if Covid-19 severity in adolescents increased | | n/a - demographic only | n/a - demographic only | I |

| | | | | | | |
|---|---|---|---|---|---|---|
| **Tirawi et al., 2022** | Would recommend vaccine to others:<br>Yes - 99.58%<br>No - 0.42% | Motivation to have vaccine:<br>Parents (69.8%)<br>Teachers (24.08%)<br>Friends (4.86%)<br>Others (1.27%)<br><br>Knowledge about vaccines:<br>Yes (99.47%)<br>No (0.53%) | | | | A / I |
| **Tu et al., 2022** | | Reasons for being vaccinated:<br>To protect family and friends (44.3%)<br>To protect self (43.1%)<br>To socialise / go out freely (25.6%)<br><br>Factors which would encourage hesitant to be vaccinated:<br>Evidence showing efficacy (17.1%)<br>More information about safety (6.1%)<br>School mandates (10.7%)<br><br>NB One third of unvaccinated/unwilling expressed no motivators which would persuade them to be vaccinated | | ANOVA/T-Tests:<br><br>Associated with vaccine uptake:<br>Internal motivation (beliefs about safety/efficacy, to avoid getting sick, to protect loved ones) ($x^2$ = 4.117, p = 0.042)<br><br>Logistic regression:<br>Risk perception higher in **v**accinated (M = 2.93, SD = 0.62) than unvaccinated (M = 2.51, SD = 0.78; t = −7.064, p <.001)<br>Being vaccinated associated with higher risk perception (r2 = 0.18, P < .001)<br>Risk perception of internally motivated (M = 3.11, SD = 0.59) significantly higher than externally motivated (M = 2.52, SD = 0.61; t = -3.23, p = <.01).<br>Internal motivation associated with risk perception (r2 = 0.07, p = .001).<br>As risk perception more strongly correlated with vaccination status (r2 = 0.1816, p < 0.001), and that vaccinated individuals were more internally motivated<br>than unvaccinated individuals (X2 = 4.117, p-value = 0.042, as | Logistic regression.<br><br>Perceived knowledge and risk perception (r2 = -.005, p = 0.81) | I / B |

| | | | | |
|---|---|---|---|---|
| | | reported above), internal motivation and higher risk perception to act in tandem to influence vaccination in adolescents.<br><br>Perceived knowledge mediates risk perception:<br>Vaccinated with low knowledge had higher risk perception than unvaccinated with low perceived knowledge (nr, graph only). Those with high perceived knowledge - no difference in risk perception based on vaccination status (r2 = -.005, p = 0.806)<br><br>Logistic regression:<br>Associated with vaccine hesitancy:<br>Risk perception (B = -0.982, SE = 0.128, Wald = 58.641 (df)1, p < .001)<br>Motivation (B = 2.559, SE = 0.424, Wald = 36.343 (df)1, p < .001) | | |
| **Unger et al., 2023** | n/a | Binary logistic regression:<br>Associated with being fully vaccinated:<br>Response efficacy *“The COVID-19 vaccine will protect me from getting COVID-19”, “The COVID-19 vaccine will protect me from being hospitalized if I get COVID-19"* (OR = 2.26, CI 95% 1.60-3.19, p = nr)<br>Self efficacy *“It is easy to get the COVID-19 vaccine” and “I am able to get the COVID-19 vaccine without much effort”* (OR = 1.68, CI 95% 1.17-2.40, p = nr) | Not associated with being fully vaccinated:<br>Perceived severity *"If you get COVID-19, what are the chances you’d be hospitalized?” “If you get COVID-19, what are the chances it would result in long-term side effects?”* (OR = 1.14, 95%CI 0.81-1.60, p = nr)<br>Perceived vulnerability *"What are your chances of getting COVID-19?”* (OR = 0.99, CI 95% 0.73 - 1.35, p = nr) | B |

| | | | | | |
|---|---|---|---|---|---|
| **Wang et al., 2022a** | To keep self and family safe (94.3%)<br>Parents'/family's will (71.7%)<br>Doctor suggested (63.1%) | Perceived low necessity (46.7%)<br>Concerns about effectiveness (11.2%)<br>Concerns about safety (45%)<br>Fear of getting unlicensed/experimental/poor qiality vaccine (2.1%)<br>Religious reasons (1.5%)<br>Influence of conspiracy theories (2.4%)<br>Personal liberty concerns (1.5%) | Binomial regression.<br><br>Associated with vaccine hesitancy:<br>Vaccine safety:<br>Compared to those who perceived COVID-19 vaccines to be very safe among adolescents, adolescents perceiving COVID-19 vaccines to be somewhat safe, not very safe, and not safe at all had 1.9 times (aPR = 1.87, 95%CI 1.57 - 2.22), 3.5 times (aPR = 3.52, 95%CI 3.02 - 4.10), and 3.5 times (aPR = 3.52, 95%CI 3.00 - 4.13) the prevalence of COVID-19 vaccine hesitancy, respectively.<br><br>Vaccine efficacy:<br>Compared to adolescents who believed that COVID-19 vaccines were very effective, adolescents who perceived COVID-19 vaccines to be somewhat effective, not very effective, and not effective at all had 1.9 times (aPR = 1.94, 95%CI 1.65-2.28), 3.3 times (aPR = 3.29, 95%CI 2.83-3.81), and 3.5 times (aPR = 3.46, 95%CI 2.97-4.03) the prevalence of vaccine hesitancy, respectively.<br><br>Impact on daily activities:<br>Experiencing impacts of COVID-19 pandemic on daily activities associated with 11% lower prevalence of COVID-19 vaccine hesitancy (aPR = 0.89, 95%CI 0.81-0.98) | Binomial regression.<br><br>Psychological distress:<br>Mild aPR = 1.05, CI 95% 0.95 - 1.16<br>Moderate aPR = 0.99, CI 95% 0.81 - 1.21<br>Severe aPR = 0.93, CI 95% 0.60 - 1.43<br><br>Perceived risk of exposure:<br>Low aPR = 0.93, CI 95% 0.66 - 1.30<br>High aPR = 0.86, CI 95% 0.57 - 1.30<br>Very high aPR = 0.81, CI 95% 0.43 - 1.53<br><br>High knowledge of Covid-19:<br>aPR = 0.83, CI 95% 0.65 - 1.07 | I |

| | | | | | | |
|---|---|---|---|---|---|---|
| **Wang et al., 2022b** | See Wang et al., 2022a | | | | | |
| **Wirunpan et al., 2021** | | | Concern over the unknown side effects (24.3%)<br>Lack of confidence in government (16.9%)<br>Personal reasons (11.8%)<br>Belief that they are not exposed to the diseases (0.7%) | Generalized linear model:<br>Associated with willingness to be vaccinated:<br>Positive attitude to vaccine (B = 0.38, p = <.01)<br>Correlation:<br>Associated with willingness to vaccinate:<br>Attitude toward vaccination (r = 0.618, p < 0.01) | Knowing someone with COVID-19 (M = 2.30, SD = 1.35) v not knowing someone with Covid-19 (M = 2.29, p = 1.23).<br>Knowledge about COVID-19 (r = 0.064, p = nr) | I |
| **Wong et al., 2022a** | n/a | Worried to be infected (59%)<br>To protect family (53%)<br>To return to the normality before COVID-19 (42%) | Concerned about vaccine safety (79%)<br>Concerned about vaccine efficacy (52%)<br>Facemasks and social distancing are sufficient (26%) | Logistic regression.<br><br>Associated vaccine intention:<br><br>One or more parent vaccinated with against Covid-19 (aOR = 5.022, CI 95% 4.211-5.989)<br><br>Received influenza vaccine in last year (aOR = 1.642, CI 95% 1.355-1.988)<br><br>Know someone diagnosed with Covid-19 (aOR = 2.098, CI 95% 1.202-3.663) | n/a - demographic only | I |

| | | | | | | |
|---|---|---|---|---|---|---|
| **Wong et al., 2022b** | See Wong et al., 2022a | | | | | |
| **Zhang et al., 2022** | | Reduce the risk of infection (66.3%)<br>Vaccine has been proved to be safe (56.6%)<br><br>Low risk of side effects from the vaccine (51.2%)<br>Reduce the risk of severe illness after infection (44.6%)<br>Vaccinations recommended by doctors/professionals (38.6 %)<br><br>Convenient place and time for vaccination (22.9%)<br>Promote the reopening of society (36.1%)<br>Stop the recurrence of the epidemic (28.3%)<br>Ensure workplace or school safety (25.9%)<br>Promote economic recovery (27.1%)<br>Vaccination recommended by the government administration (25.9%)<br><br>Others (unpaid vaccination, etc) (10.8%) | Concerns over side effects (70.5%)<br>No risk of infection (34.9%)<br>No severe illness after infection (15.7%<br>Relying only on protection from innate immunity (11.4%)<br>Exaggerated impact of the epidemic (11.4%)<br>Special physical conditions not suitable for vaccination (5.4%)<br>Unpleasant vaccination experience (19.3%)<br>Distrust in vaccine efficacy (16.3%)<br>Lack of access to vaccination-related information (13.9%)<br>Others(no time, etc) (9.0%)<br>Vaccination conspiracy theories (0.1%) | Multivariate logistic regression:<br>Associated with vaccine hesitancy:<br>Abnormal illness behaviour (e.g., hypochondriacal beliefs, preoccupation with one's own health status etc - maladaptive) (OR = 1.17, CI 95% 1.07 - 1.28, p = .001)<br><br>Protective factor against vaccine hesitancy:<br>Utilisation of support (OR = 0.86, CI 95% 1.30 - 5.15, p = .007) = r2 = 0.036 | Multivariate logistic regression:<br><br>Acceptance v hesitancy<br>Stress 3.06 +/- 2.23 v 3.63 +/- 2.50, p = 0.009<br>Global wellbeing 6.90 +/- 1.85 v 6.56 +/- 2.02, p = 0.063<br>Psychological distress - anxiety, depression, sleep disturbances, 12.81 =?- 8.59 v 15.57 +/- 9.60, p = 0.001<br>Objective support - visible, direct support 7.95 =/- 2.59 v 7.28 +/- 2.58, p = 0.001<br>Subjective support (perceived support from family/friends/colleagues) 21.05+/- 4.23 v 19.98+/- 4.32, p = 0.003<br><br>Attendance at Covid-19 vaccination lecture (knowledge) 42.1 v 40.4, p = 0.676 | I |
| **Zilhadia et al., 2022** | n/a | | | Bivariate analysis.<br><br>Associated with vaccine uptake (Mean rank: not received v received):<br>Perceived susceptibility (199.81 v | Bivariate analysis.<br><br>Not associated with vaccine uptake:<br>Cues to action (i.e. recommendation from religious leader, HCW, friend, teacher, family) (204.94 v 228.79, Z = | I / B |

231.54, Z = -2.86, p = 0.004)
Perceived severity (191.14 v 236.15, Z = -3.76, p = < 0.001)
Perceived benefits (197.57 v 232.72, Z = -2.94, p = 0.003)
Perceived barriers: high score = low perceived barrier (163.05 v 251.13, Z = 7.09, p = <0.001)
Self efficacy (188.48 v 237.57, Z = 4.11, p = <0.001)
Attitude (192.68 v 235.33, Z = 3.56, p = <0.001)
Social norm (198.45 v 232.26, Z = -2.72, p = 0.006)

Associated with *intention* to have vaccine:
Mean rank - do not intend v intend:
Perceived susceptibility (44.15 v 89.84, Z = -6.31, p = <0.001)
Perceived severity (50.81 v 87.24, Z = -4.84, P = <0.001)
Perceived benefits (49.51 v 87.75, Z = -5.18, p = <0.001)
Perceived barriers (64.26 v 81.98, Z = -2.27, p = 0.023)
i to action (65.41 v 81.53, Z = -2.06, p = 0.039)
Self-efficacy (43.51 v 90.09, Z = -6.18, p = <0.001)
Attitude (48.17 v 88.27, Z = -5.27, p = <0.001)
Social norm (57.93 v 84.45, Z = -3.41, p = 0.001)
Perceived behavioural control (92.12 v 71.09, Z = -2.85, p = 0.004)

-1.93, p = 0.054)
Perceived behavioural control: *ie free/convenient* (206.95 v 227.72, Z = -1.76, p = 0.078)

Not associated with vaccine intention to receive: none

**Appendix C:** Full data extraction results

**Table C2:** Overview of qualitative data on psychological factors influencing vaccination attitudes, intention and uptake.

| Citation | Themes | | |
|---|---|---|---|
| **Alemu et al., 2023** | 1. At low risk of Covid-19 | 2. Don’t trust the vaccine | 3. Vaccine not permitted by my religion |
| **British Academy, 2022** | 1. Youth vaccine hesitancy based on place conditions and social environment. Young people do not lack Covid-19 vaccine information but overwhelmed by amount of information - inundated by information from plurality of sources (news/parents/social media). | 2. Younger youth are less likely to be vaccine hesitant: Less access to smartphones, so less exposed to misinformation. More likely to listen to parents. | 3. Vaccination to ‘return to normal’, to attend social events and be more socially connected. BUT those who didn't follow Covid-19 guidelines already living more 'normal' life so less incentive to accept vaccine. In UK - in less deprived area, older youth spoke of "following the science" and spoke negatively of anti-vax peers who they saw as listening to conspiracy theories over scientific guidance. |
| | 4. Experiences of deprivation may drive vaccine hesitancy<br>Young people in areas of deprivation more likely to be vaccine hesitant<br>More vaccine hesitant than certain they are anti-vaccination<br>Less likely to have parent(s) telling them to be vaccinated / peers who are vaccinated / less exposed to positive vaccination stories. | 5. Young people’s in-group shaped vaccine attitudes, base behaviours on what their 'in group' was doing - most vaccinated adolescents said their friends/family were also vaccinated and vice versa - but not also explicitly mentioned as reason for vaccine decisions. Young people in less deprived areas more future oriented. | 6. Structural inequalities shape youth experiences of disadvantage. Young people in areas of disadvantage already labelled "bad" or "trouble" in public discourse and this contributes to their sense of social exclusion and marginalisation - sometimes mirrored in vaccine non-compliant as another way that they are "bad". |

| | | | |
|---|---|---|---|
| | 7. Trust (medical (mis)trust drives youth vaccine hesitancy). Information. Community Trust - community anti-trust of authorities' narrative - idea of collective wisdom. | 8. Intuition / gut feeling | 9. Concerns over safety of the vaccines, particularly in the long-term. Disinterest or lack of relevance to their lives.<br><br>Perception that they were strong and healthy. Perception that they could develop natural immunity without vaccination. Perception that the government was forcing them to get vaccinated. |
| | 10. What are young people and their families most concerned about? Security and safety more important than COVID-19. | 11. Young people want to be safe, but 'safety' has different interpretations. Can be a reason to be vaccinated - ie to protect health, or a concern about the vaccine if deemed risky or unsafe. | |
| **Budhwani et al., 2021** | 1. Influence of community leaders and older family members:<br>Influenced own likelihood of accepting vaccine | 2. Fear of side effects and misinformation:<br>Misinformation and fear of side effects intertwined | 3. Institutional mistrust:<br>Scepticism towards government and healthcare systems |
| **Delgado et al., 2023** | 1. Family values and practices around adolescent vaccination:<br>Sub themes:<br>i) Parent-driven approach to vaccination decisions with limited adolescent engagement<br>ii) Encouraging adolescent medical autonomy around health decisions including vaccines<br>Letting adolescent decide as they are near adulthood<br><br>iii) Psychosocial environment and peer group influences on vaccination values | 2. Differences in parent and adolescent support for vaccine self-consent laws:<br>Subthemes:<br>i) Adolescent support for vaccine self-consent<br>Adolescents support self-consent as they see other teens could benefit, who are currently at a disadvantage for not getting vaccine (eg may have to move schools) because older consent-givers are anti-vax. | 3. Parent and adolescent uncertainty on readiness for vaccine self-consent laws:<br>Teens uncertain about self-consent as view other teens as "irresponsible" / less informed |

| | | | |
|---|---|---|---|
| **Fisher et al., 2021** | 1. Acceptability of vaccination against COVID-19<br>All willing to be vaccinated. Motivated by:<br>Altruism (to protect others at greater risk of Covid-19)<br>Reduce impact of disease amongst population<br>To help return 'back to normal'<br>May not agree to be vaccinated if:<br>Potential for harm (side effectives, weak effectiveness)<br>vaccine should be prioritised for those at greatest risk<br><br>pro-vaccine, felt that negative views about vaccines in general unlikely to change their mind. | 2. Differences in parent and adolescent support for vaccine self-consent laws:<br>Subthemes:<br>i) Adolescent support for vaccine self-consent<br>Adolescents support self consent as they see other teens could benefit, who are currently at a disadvantage for not getting vaccine (eg may have to move schools) because older consent-givers are anti-vax.<br>*ii) Parent support for vaccine self-consent* | 3. Parent and adolescent uncertainty on readiness for vaccine self-consent laws:<br>Teens uncertain about self-consent as view other teens as "irresponsible" / less informed |
| **Garcia et al., 2021** | 1. Tempered optimism for vaccines to protect family well-being:<br>Valued (by teens) as a way to get young people back into school<br>*As a way to end the pandemic, as a tool to alleviate families' suffering, concerned about lack of access to healthcare in case of side effects.* | 2. Hesitancy rooted in mistrust<br>vaccine hesitancy rooted in mistrust of medical and political institutions: young people spoke about conspiracy theories parents receiving misinformation via social media eg microchips in vaccines: see parents as unable to tell real news from fake.<br>Intergenerational communication central to determining trustworthiness of info and decison-making - teens as a source of info for parents | |
| **Groenewald et al., 2023** | 1. Children's perspectives on Covid-19 vaccines: CYP generally referred to the benefits of vaccines, and saw them as a source of hope, but also had limited vaccine knowledge. Fears were around mistrust or doubts around vaccine efficacy. | 2. Intergenerational influence. Children expressed similar attitudes to their parents about Covid-19 vaccines | |
| **Kenworthy et al., 2022** | 1. Vaccine hesitancy & %age of students endorsing theme: i) Potential side effects (44.4%) "s of concern included death, allergic reactions, infertility, and less frequently, vaccines turning individuals into zombies or turning the injection site magnetic"<br>ii) Vaccine development (11.1%) included " unknown long-term effects of the vaccine and the speed in which vaccines were | 2. Cultural/family traditions & %age of students endoring theme:<br>*i) Family practices (0%)*<br>ii) Cultural practices (11.1%) "some cultural traditions, like drinking a medicinal tea, were | 3. Compatibility of policies and places with recommended health behavior & %age of students endorsing theme: *i) (44.4%) - influence of context or physical environment on ability to engage in health behaviours eg social distancing in crowded school* |

| | | | |
|---|---|---|---|
| | developed"<br>iii) Content of vaccine (22.2%) "unknown content such as allergens or animal DNA", age of recipient (11.1%) | described as preventative or treatment approaches"<br>*iii) Beliefs ()%)* | |
| | 4. Perceived risk & %age of students endorsing theme (66.6%)<br>i) apathy (1.1%)<br>ii) concern impacting thoughts (22.2%) eg worry about illness/death from Covid-19<br>iii) concern impacting behaviours 55.5% eg concern for health motivated them to have a vaccine / others do not engage in heath behaviours because they feel safe or protected from Covid-19 | 5. Reliability of information & %age of students endorsing theme (22.2%);<br>*i) Questionnable information (0%)*<br>ii) Mixed messages freom agencies and conflicting recommendations 22.2% eg local/state v Centers for Disease Control & Prevention (CDC)<br>iii) politicised information (11.1%) in some cases led to ignoring news altogether<br>*iv) Mistrust (0%)*<br>Students less concerned about reliability of information than staff/caregivers. | |
| **Mansfield et al., 2023** | 1. Family anticipation and hesitation about the approval of COVID-19 vaccines for adolescents:<br>Enthusiasm to be part of something important for society;<br>Adolescents confident about vaccine<br>themselves did not share vaccine safety concerns of parents; the only hesitation expressed by multiple adolescents was a dislike of needles and nervousness that the injection might be painful. | 2. Parents' choice or adolescents' choice: The decision maker for adolescent COVID-19 vaccination<br>Mix of parent-driven, adolescent-driven and shared decision-making with parents. No conflict reported between parent/child.<br>Parent-drive: (male adolescent) "I'd like agreed with him. They were gonna make me do it anyway"<br>Adolescent driven (chn of nurses only, not physicians' chn. | 3. Leveraging one's vaccination status to encourage others to get vaccinated<br>Yp prepared peers to get vaccine "just like a flu shot" and prepared them for side effects eg pain<br>Yp expressed benefits of vaccine and importance of relying on accurate information<br>Experiences of talking to friends about vaccine (but wouldn't force it on others) |
| **McKinnon et al., 2023** | 1. Being misinformed contributes to COVID-19 vaccine hesitancy among nonvaccinated adolescents:<br>Rapid development and approval of vaccine (as compared to other vaccines) linked to lack of rigour/transparency;<br>Perceived lack of effectiveness of 19 vaccine supported by the fact that vaccinated people can still get infected with the virus and | 2. Nonvaccinated adolescents value autonomy in making decisions about Covid-19 vaccination:<br>Adolescents valued making free and independent decisions about COVID-19 vaccination, & expressed strong aversion to efforts perceived as pressuring them to take the vaccine, whether from friends and | 3. Vaccine mandates consistently described as unfair but effective<br>Strongly opposed to vaccine mandates<br>Vaccine passports seen as "manipulative" and "unfair" but also main motivation for receiving vaccination<br>Most reported that parents had accepted vaccine to comply with travel/work mandates |

| | | | |
|---|---|---|---|
| | become seriously ill;<br>Personal anecdotes contributing to apprehension (eg a vaccinated family member died of Covid-19);<br>Did not belief conspiracy theories and were not 'anti-vax' - had received other vaccines;<br>Confusion about how vaccines work;<br>Desire for answers re safety and efficacy | family or government mandates;<br>Influenced by friends/family not to have vaccine, but didn't want to be influenced to have the vaccination;<br>Decision framed as individual choice based on context;<br>None mentioned community implications of vaccination;<br>Mentioned impact of Covid-19 on life but did not link to getting vaccine | Ppts considered getting vaccine for work/travel/passport. Commented on "absurdity" of getting vaccinated to eat in a restaurant or play sport rather than to protect health |
| **Nilsson et al., 2021** | 1. Adolescents expressed a need to know more<br>Yp had weighed risks and benefits of vaccines - didn't want to be megative towards vaccines but wanted to know more eg rapidity of development and concern re side effects made them uncertain and want to do more research<br>Fear: of injections/shs and of as yet unknown "bad" side effects. | 2. The adolescents did not consider themselves to be in need of vaccination:<br>Yp identified as healthy and not at risk, not "their" disease. They did not fear Covid-19 as death rates were low. Better to be infected than vaccinated."<br>Altrusim - yp expressed willingness to be vaccinated for the sake of others - felt a general responsibility to be vaccinated for protection of others and economy and contribute to herd immunity. | |
| **Persaud et al., 2023** | 1. Lack of personal utility (benefit) from vaccine or not ready, eg "I'm still debating whether or not I should get it) | 2. Mistrust, misinformation, rushed eg "I just don't trust it enough". | 3. Unanswered medical questions eg "I don't know the risks it could have on me, I would like to know the pros and cons about the vaccination" |
| **Ramaiya et al., 2023** | 1. Family. Attitudes (whether positive or negative) of family members influenced vaccine attitudes, but adolescents were critical of distrusting older relatives. | 2. Community. Doctors seen as trustworthy. Church could be either a positive or negative influence. | 3. Institutional. Strong institutional trust in China, some trust in science but also distrust of vaccine safety due to fast development. Distrust of government, and media/social media seen as a source of misinformation. |
| | 4. Policy. Mixed views on vaccine mandates: in favour in China/Indonesia but sceptical of mandates in USA. | | |

| | | | |
|---|---|---|---|
| **Uroko et al., 2023** | 1. vaccine consequences related to the apocalypse: a new world order would begin if enough people are “forced” to accept the vaccine. | 2. Vaccine viewed as dehumanising people of human qualities, by altering their DNA so they cannot make it to heaven. | 3. 666 (Mark of the Beast). Vaccine causes untimely death and makes one an agent of the devil (by inserting a chip). |
| | 4. Vaccine seen as a way to shorten one’s lifespan thereby reducing the population of Nigeria. | | |
